# Spatial and temporal epidemiology of SARS-CoV-2 virus lineages in Teesside, UK, in 2020: effects of socio-economic deprivation, weather, and lockdown on lineage dynamics

**DOI:** 10.1101/2022.02.05.22269279

**Authors:** E.D. Moss, S.P. Rushton, P. Baker, M. Bashton, M.R. Crown, R.N. dos Santos, A. Nelson, S.J. O’Brien, Z. Richards, R.A. Sanderson, W.C. Yew, G.R. Young, C.M. McCann, D.L. Smith

## Abstract

**Background:** SARS-CoV-2 emerged in the UK in January 2020. The UK government introduced control measures including national ‘lockdowns’ and local ‘tiers’ in England to control virus transmission. As the outbreak continued, new variants were detected through two national monitoring programmes that conducted genomic sequencing. This study aimed to determine the effects of weather, demographic features, and national and local COVID-19 restrictions on positive PCR tests at a sub-regional scale.

**Methods:** We examined the spatial and temporal patterns of COVID-19 in the Teesside sub-region of the UK, from January to December 2020, capturing the first two waves of the epidemic. We used a combination of disease mapping and mixed-effect modelling to analyse the total positive tests, and those of the eight most common virus lineages, in response to potential infection risk factors: socio-economic deprivation, population size, temperature, rainfall, government interventions, and a government restaurant subsidy (“Eat Out to Help Out”).

**Results:** Total positive tests of SARS-CoV-2 were decreased by temperature and the first national lockdown (the only one to include school closures), while deprivation, population, the second national lockdown, and the local tiered interventions were associated with increased cases. The restaurant subsidy and rainfall had no apparent effect. The relationships between positive tests and covariates varied greatly between lineages, likely due to the strong heterogeneity in their spatial and temporal distributions. Cases during the second wave appeared to be higher in areas that recorded fewer first-wave cases, however, an additional model showed the number of first-wave cases was not predictive of second-wave cases.

**Discussion:** National and local government interventions appeared to be ineffective at the sub-regional level if they did not include school closures. Examination of viral lineages at the sub-regional scale was less useful in terms of investigating covariate associations but may be more useful for tracking spread within communities. Our study highlights the importance of understanding the effects of government interventions in local and regional contexts, and the importance of applying local restrictions appropriately within such settings.

## Introduction

It is widely understood that many factors can affect COVID-19 transmission via social and/or epidemiological mechanisms, including weather, lineage of SARS-CoV-2 (the virus causing COVID-19), mask wearing, and government control measures (non-pharmaceutical interventions) that limit social contact (Bo et al., 2021; Volz et al., 2021; Ganslmeier et al., 2021; Ge et al., 2022). The risk of contracting COVID-19 can also be affected by demographic factors such as socio-economic deprivation and race (Whittle & Diaz-Artiles, 2020; Brainard et al., 2022; Holt et al., 2022). However, these relationships are complex and there is considerable variation in the observed impact of these factors between different countries, at national vs. regional scales, and during different waves of the pandemic (Gao et al., 2021; Bo et al., 2021; Hunter et al., 2021; Ganslmeier et al., 2021; Brainard et al., 2022; Ge et al., 2022). It is imperative that these relationships are better understood to improve future pandemic/epidemic responses, especially in countries like the United Kingdom, which experienced high case numbers and excess deaths relative to its neighbours (Islam et al., 2021; Hunter et al., 2021). While there is evidence that strict national restrictions reduced cases and mortality on a national-scale in the UK (Hunter et al., 2021; Sharma et al., 2021), there has been very little evaluation at local or regional scales or in socio-economically deprived populations.

Government interventions (national lockdowns) introduced during the first wave of the pandemic in the UK were successful in reducing cases, which was also seen in other countries, with school closures and measures that reduce social gatherings proving the most effective (Brauner et al., 2021; Hunter et al., 2021; Mendez-Brito et al., 2021; Ge et al., 2022). However, the picture is less clear for later waves of the pandemic, as the effectiveness of control measures changed and as countries diverged in terms of both the implementation of interventions and patterns of cases (Pozo-Martin et al., 2021; Mendez-Brito et al., 2021; Ge et al., 2022). The UK government introduced a heterogenous “tier” system of restrictions in England during the second wave, which were applied on local scales and were intended to be more responsive and appropriate to the local disease context (UK Government, 2020c). While analysis of the tier system across the entirety of England has revealed the more stringent tiers to be more effective than the less stringent ones (Davies et al., 2021; Laydon et al., 2021), there has been very little consideration of these tiers, or comparison with the national-level restrictions, within their specific geographic and community contexts.

The relationship between socio-economic deprivation and COVID-19 mortality has been well-documented, with a majority of studies demonstrating increased mortality in areas of high deprivation, including in the UK (Brainard et al., 2022; McGowan & Bambra, 2022). This is due to a combination of underlying factors that can be summarised as unequal exposure due to employment and living conditions and unequal vulnerability due to pre-existing health conditions (McGowan & Bambra, 2022). However, the relationship between deprivation and COVID-19 infection is more complicated and less clear; the UK context is typical of the broader picture with some studies finding an increase in infections linked to higher deprivation while others find the opposite or no relationship (Niedzwiedz et al., 2020; Brainard et al., 2022; McGowan & Bambra, 2022).

The relationship between COVID-19 infection and weather is also complex. Higher temperatures have been associated with a reduction in cases in many countries including the UK (Ganslmeier et al., 2021; Alaniz et al., 2023; Nottmeyer et al., 2023), though a minority of studies have reported a positive effect (Tan & Schultz, 2022). Increased rainfall effects are more uncertain due to less research focus, with a meta-analysis finding evidence of increased infections (Tan & Schultz, 2022), while other research detected no relationship (Ganslmeier et al., 2021). The impacts of weather on COVID-19 infections are complicated by the interaction between several mechanisms. Weather can affect transmission directly, e.g. increased temperatures reduce the viability of the virus particles (Ganslmeier et al., 2021), and also indirectly via social behaviours, e.g. colder temperatures or higher rainfall could discourage outdoor socialising and reduce transmission, or encourage indoor socialising and increase transmission. Several studies have also shown that the presence and strength of these indirect effects is mediated by other factors, such as how closely the timing of weather patterns and events coincides with mealtimes and the presence/absence of government control measures (Ganslmeier et al., 2021; Fetzer, 2022). This is particularly relevant in England, where the government introduced a subsidy that encouraged people to eat in restaurants during August 2020 (Fetzer, 2022).

Previous research has demonstrated that transmissibility of COVID-19 varies greatly between different lineages of the SARS-CoV-2 virus, which can explain how some lineages become dominant at certain points in time (Volz et al., 2021; Panovska-Griffiths et al., 2022). For example, lineage B.1.1.7 (the Alpha variant) has been estimated to be between 20-100% more transmissible than the wild type (estimates vary by study), which likely explains how this lineage became dominant across the UK in autumn/winter 2020 (Volz et al., 2021; Hinch et al., 2022; Panovska-Griffiths et al., 2022). However, there are also examples of lineages that became dominant despite no, or only a small, increase in transmissibility, like that of lineage B.1.177 during autumn 2020 in the UK (Hodcroft et al., 2021; Vöhringer et al., 2021; Hinch et al., 2022). Establishment of a lineage is subject to epidemiological factors as well as transmissibility, such as the number of introductions (Grubaugh et al., 2020), which could be facilitated by a relaxation of travel restrictions (Hodcroft et al., 2021). It seems likely that other non-pharmaceutical interventions and other environmental factors could also affect the number of cases of different COVID-19 lineages, regardless of whether they demonstrate increased transmissibility. While some research has investigated the impact of interventions and vaccinations on the spread of specific lineages (Hinch et al., 2022; Panovska-Griffiths et al., 2022), there has been very little comparison across many lineages, and no investigations that also incorporate other factors like weather and socio-economic deprivation.

Our aim was to investigate how a range of variables that can influence COVID-19 cases affected positive tests in the Teesside sub-region of North East England during 2020. We wanted to understand how the national and local government interventions introduced over the course of this year affected the population of this sub-region, which has high levels of deprivation and is reasonably geographically isolated. Examining cases in his context also allows us make policy recommendations aiming to improve outcomes in future epidemics, for this and other vulnerable populations in the UK. We used Generalised Linear Mixed Effect Models (GLMMs) to investigate how local and national government interventions, weather, levels of socio-economic deprivation, and the government restaurant subsidy impacted: 1) the total number of COVID-19 cases; and 2) the number of cases of each of the most common lineages in Teesside. We also used Bayesian spatial models to determine patterns of disease risk across Teesside during 2020.

## Methods

### Location and timeframe

The wider Teesside area is a sub-region of the North East region of England, centred around the mouth of the river Tees. While Teesside has reasonably good local and national transport links, it is relatively isolated geographically as it borders the North Sea to the north east, The North York Moors national park to the south east, and extensive farmland to the west. Teesside has a distinct cultural identity due to its industrial history, which has also left its population with a greater burden of diseases and socio-economic deprivation that stigmatises and further culturally isolates the area (Bush et al., 2001). Teesside contains a mixture of urban, suburban, and rural environments and is formed from a collection of separate communities including Middlesbrough, Redcar, Thornaby-on-Tees, Billingham, Hartlepool, and Stockton-on-Tees, each with their own identities, facilities, schools, etc. All of these characteristics make Teesside an interesting case study that deserves research focus.

We chose to examine positive tests within Teesside during only 2020 for several reasons. Firstly, the local tiered restrictions were abandoned when the country entered a new national lockdown in early January 2021, and were not reinstated (UK Government, 2021), which means there were no further locally applied restrictions that could be examined. Secondly, the UK’s vaccination programmed begun in December 2020, which means that analysing data that included cases into 2021 would have required additional data to properly adjust for this confounder. Inclusion of vaccination uptake information would have been needed, rather than numbers/proportion of eligible people in the population, as uptake is lower amongst deprived people (Mounier-Jack et al., 2023), but this would have been difficult to ascertain at the necessary spatial and temporal scales. And finally, including data collected over a longer time period would open up the analysis to other potential sources of confounding that cannot easily be controlled for, such as ‘pandemic fatigue’.

### Data collation

The UK government introduced two ‘pillars’ of COVID-19 testing, which utilised polymerase chain reaction (PCR) to identify positive cases. Pillar 1 testing was of staff and patients in hospitals and care homes, mainly focussing on symptomatic individuals, but also included asymptomatic staff (e.g. contacts of confirmed cases). Pillar 2 was community testing of symptomatic individuals, which also began to include asymptomatic testing of suspected cases and high-risk situations (e.g. confirming lateral flow results, elective care settings, care homes, and contacts of confirmed cases) from autumn 2020 (Dept. Health & Social Care, 2020; UK Health Security Agency, 2023). Genomic sequencing was conducted on a random sample of the PCR samples that tested positive for SARS-CoV-2, which permitted characterisation of genetic lineages in individual cases, and the information was stored in the Covid-19 Genomics UK (COG-UK) dataset (COVID-19 Genomics UK (COG-UK) Consortium, 2020; Smallman-Raynor et al., 2022). We collated records of PCR tests positive for SARS-CoV-2 in the Teesside area during 2020, from the COG-UK genomic dataset, which provided the dates, postcodes, and viral lineages of all sequenced positive tests from January 2020 to January 2021. We summarised the number of cases for all lineages, and each lineage separately, by week of year and individual postcode districts (TS1 to TS29, see Figure 1). In addition, we also summarised the total number of cases of each lineage recorded by each pillar during the period when sampling was run contemporaneously for both pillars (from week of year 19 to 53), creating a dataset of total cases of each lineage summed across time for each pillar (each lineage had one value per pillar). We ran a Spearman’s correlation on this dataset, to check for a sampling bias for different lineages between the two pillars, as only pillar 1 tested the most severely ill patients, and illness severity can vary between lineages (Sievers et al., 2022; Goethem et al., 2022).

**Figure 1.**
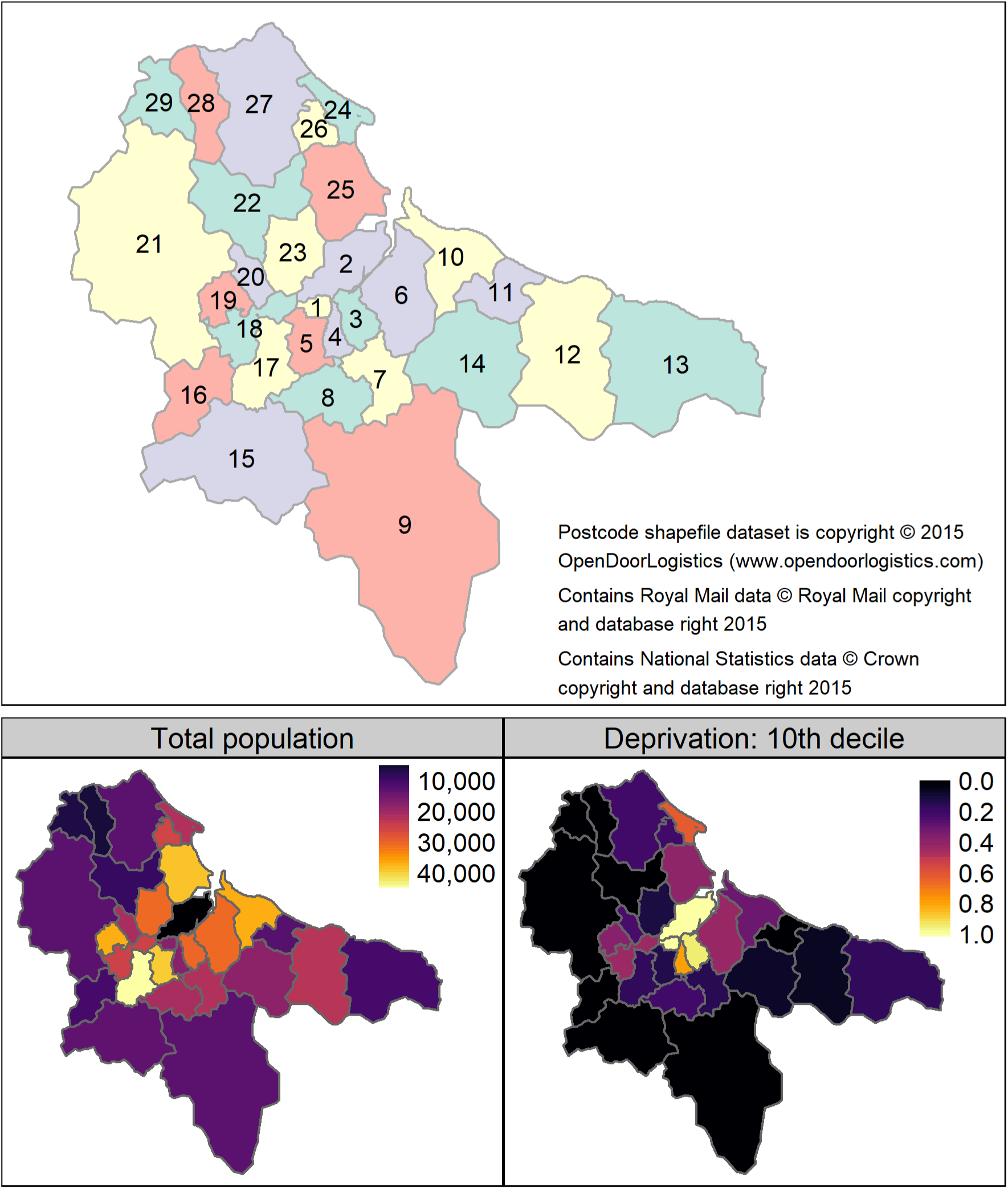
Labelled map of the 29 Teesside postcode districts (“TS” prefixes have been omitted for clarity and colours are purely to aid discrimination between polygons), and maps of the total resident population and proportion of the population within the 10th decile of the Index of Multiple Deprivation (when assessed at the scale of England as a whole) for each Teesside postcode district (Source: Office for National Statistics licensed under the Open Government Licence v.3.0). The north east boundary meets the North Sea, to the south east is a national park, and to the west is farmland.

We used recent demographic data on socio-economic deprivation (Index of Multiple Deprivation IMD (UK Government, 2019)) to calculate the proportion of the population in each postcode district that was in the most deprived category (10^th^ decile), relative to England as a whole (Figure 1 and Table S1 in Supplementary Material). Mean weekly temperature and rainfall values were summarised from raw daily data (calculated from daily means and daily totals respectively) obtained from weather stations in Teesside International Airport (temperature) and Hartlepool (rainfall) (we assumed weather to be homogenous across the area at this temporal scale). We collated the start and end dates of the national government interventions (lockdowns) and the local tier system of interventions (where higher tiers had greater restrictions) as applied to Teesside in 2020. The tier system was applied at the local government level, but during 2020 the same tier restrictions were applied to all local authority areas in Teesside. We also collated the start/end dates of the government restaurant subsidy (“Eat Out to Help Out Scheme”) that was introduced to support hospitality businesses, which provided customers with a 50% discount at participating restaurants. The interventions and subsidy dates were converted to week of year format, where values counted the number of weeks since the measure was introduced (weeks before introduction and after stoppage were given a value of zero). The national lockdowns in England were defined by: stay-at-home orders, closure of all non-essential businesses, prohibition of all social gatherings and events (UK Government, 2020a; d), and the first lockdown also included school closures (UK Government, 2020b) (Table 1). The local tier 2 and 3 restrictions in England (Local Covid Alert Levels ‘High’ and ‘Very High’) allowed for some non-essential businesses to remain open and permitted outdoor socialising in small groups (UK Government, 2020c) (see Table 1 for details). The end date of the first lockdown is complicated, as different restrictions were lifted at different times (Table 2). We used the earliest end date applicable to Teesside to define the final week of the first lockdown (week 23).

**Table 1.** Summary of the control measures included in each national and local restriction category in England. Business closures during tier 3 varied across England, those shown below were applied to Teesside during 2020.

| Included measures | 1st Lockdown | 2nd Lockdown | Tier 1 | Tier 2 | Tier 3 | Tier 4 |
| --- | --- | --- | --- | --- | --- | --- |
| Stay at home orders | X | X |  |  |  | X |
| School closures | X |  |  |  |  |  |
| Face masks indoors in public spaces |  | X | X | X | X | X |
| *Business/venue closures:* |  |  |  |  |  |  |
| Pubs, bars, and restaurants | X | X |  |  | X | X |
| Retail | X | X |  |  |  | X |
| Personal care | X | X |  |  |  | X |
| Leisure and entertainment | X | X |  |  | X | X |
| Gyms | X | X |  |  |  | X |
| Places of worship | X | X |  |  |  |  |
| *Prohibition of social gatherings:* |  |  |  |  |  |  |
| Indoors up to 6 people | X | X |  | X | X | X |
| Indoors more than 6 people | X | X | X | X | X | X |
| Outdoors up to 6 people | X | X |  |  |  | X |

**Table 2.**
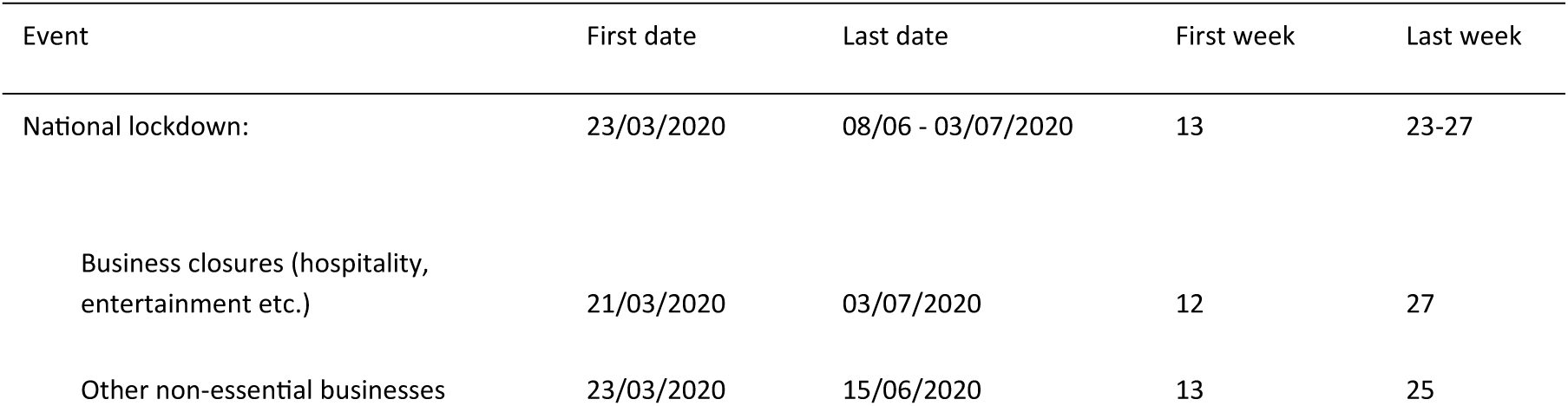
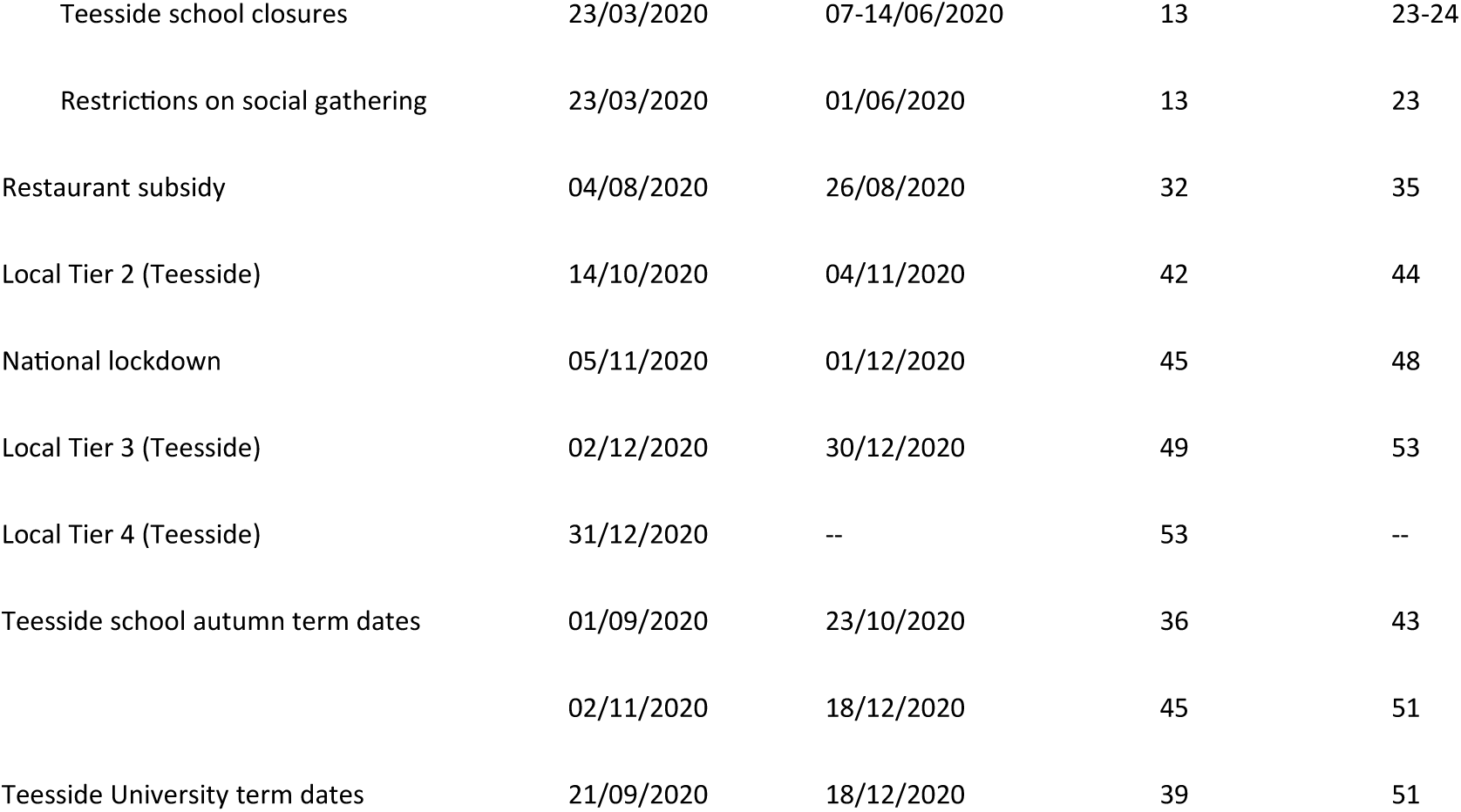
First and last dates of government control measures applied in Teesside, restaurant subsidy, and school and university terms in Teesside during 2020.

### Disease mapping

To help us define and visualise the spatial heterogeneity of COVID-19 in Teesside, we used Bayesian spatial models (Riebler et al., 2016) to assess the area-specific relative risk of a positive test in each postcode district across the study period. We separately modelled all disease cases (regardless of lineage), and cases of each of the most common lineages in Teesside that had sufficient data. We used the most recent version of the Besag, York, Mollie “BYM” model (Besag et al., 1991), the “BYM2” (Riebler et al., 2016), which is a conditional autoregressive model that adjusts for spatial dependency between adjacent spatial units, as numbers of cases in one district are likely to be partially dependent on those arising in neighbouring districts, with which there may be contact leading to enhanced transmission. We included the expected number of cases for each postcode district (calculated using district population sizes, total population across all districts, and total cases in all districts) in our models as an offset, which converts the output to the risk of a positive test in each district relative to the overall risk across all districts (Blangiardo & Cameletti, 2015). These models were fitted using Integrated Nested Laplace Approximation via the “INLA” package (version 22.05.07) (Rue et al., 2009) in R version 4.2.2 (R Core Team, 2022). As the response variables in these models are aggregated counts or rates, we fit the models with negative binomial error distributions (using log link functions) and penalised complexity priors, which reduce the chance of overfitting (Riebler et al., 2016). We used the default penalised complexity priors for the BYM2 model (Riebler et al., 2016) throughout, as a sensitivity analysis using different hyperparameter values showed no improvement to model fit. Model validation, via PIT (probability integral transforms) and plotting observed values against a sample generated from the posterior distribution, demonstrated a poor fit to the data for three of our lineages (B.1.1.309, B.1.1.315, and B.1.1.37), which have been dropped from the results section of the paper (see section 3.1 in the supplementary material for model validation and hyperparameter sensitivity analyses). The shapefiles for the Teesside postcode districts that were used in this analysis (and figures) were downloaded from: https://www.opendoorlogistics.com/data/.

### Mixed-effect modelling

We used generalised linear mixed-effect models (GLMMs) to investigate the role of socio-economic deprivation (proportion of the population in the 10^th^ Decile of IMD), weather (mean weekly temperature and rainfall), UK government interventions (national lockdowns and local tiers), and government restaurant subsidy on the frequency of positive PCR tests recorded each week of 2020 in each postcode district. These models were non-Bayesian and were fitted using maximum likelihood estimation. We separately modelled all disease cases (regardless of lineage), cases of each of the most common lineages in Teesside that had sufficient data, and the total cases of the most common second wave lineages. We included an autoregressive term of order 1 (AR1) for week of year for each postcode district to account for temporal auto-correlation (non-independence) of cases over time within each separate district. We included a random intercept for postcode district to account for repeated measures of postcodes across time and for unmeasured geographical variation that might have influenced the recording of cases. We controlled for population size by including district total population (rescaled to measure population in thousands rather than single people) as a fixed effect. The second-wave lineages model also included the total cases of the most common first wave lineages for each postcode district as an additional fixed effect, which allowed us to investigate the potential for increased immunity between successive waves of the epidemic due to previous infections. We fitted an additional model to the all-case data, including the same fixed effects and the random intercept for postcode, but this model did not include an AR1 term and instead used a random gradient for week for each postcode district (allowing the effect of week on cases to vary by postcode), which allowed us to map and compare the rate of change in cases reported each week across the different postcodes.

To account for variation in symptom onset and testing delay after infection, we applied a two-week time lag to all temporal variables (temperature, rainfall, and the government interventions and subsidy). Because we aggregated case numbers and summarised temporal variables by week, a one-week delay could have artificially assumed a greater separation between cases and temporal events than actually happened. Additionally, the sensitivity analyses in Hunter et al. (2021) demonstrated that non-pharmaceutical interventions in the UK did not show an impact on numbers of cases until after 14 days.

As our response variable (positive tests) is a count, we fit our models with Poisson and negative binomial distributions (using log link functions) before validating them with the “DHARMa” R package (Hartig, 2022), which uses a simulation approach to create interpretable scaled residuals. We used DHARMa’s test and plotting functions to assess deviations from the expected distribution, dispersion, heteroskedasticity, temporal autocorrelation, and zero-inflation, alongside plots of observed values against those fitted from the models, which demonstrated that Poisson was a better fit for the data in all models, except for the all-cases model fit with a random gradient (for full details and output see section 3.2 of the supplementary material). Because it would be reasonable to assume that the number of cases over time would follow a non-linear trajectory, we also fit several alternative model specifications that included week of year as a fixed effect with either a smooth or restricted cubic spline instead of an AR1 term, however, these models were a poorer fit for all datasets than the AR1 models (see section 3.2 of the supplementary material). All regression models were fit using the package “glmmTMB” (Brooks et al., 2017) in R version 4.2.2.

We chose to retain all variables in the models regardless of AIC or significance as they are all biologically/epidemiologically meaningful. Predictor variables were only removed if they were found to be collinear via variance inflation factor (VIF) (calculated using the “performance” R package (Lüdecke et al., 2021)), whereupon we followed the procedure defined by Zuur et al. (2010) of removing variables sequentially until none of the recalculated VIF values are above 3. We also confirmed variable removal via redundancy analysis and variable clustering (conducted using the “Hmisc” R package (Harrell & Dupont, 2021)) (see section 3.2 of supplementary materials for further details, redundancy analysis outputs, and variable clustering plots).

Full details of all the model specifications (disease mapping and GLMMs) can be seen in the analysis scripts (https://doi.org/10.25405/data.ncl.23815077) and section 2 of the supplementary material.

## Results

### Data and trends

The Teesside postal area had a population of 599,600 people across the 29 postcode districts, with considerable variation in the proportion of the total population in the 10^th^ IMD decile (Figure 1, Table S1). The populations of urban postcode districts of central Middlesbrough (TS1, TS4 and TS3) all had high proportions of their populations in the 10^th^ decile for IMD, whereas many of the suburban and rural postcode districts had little to none of their population in this decile. There were 2,328 positive COVID-19 tests for all lineages of which 1,073 were pillar 1 and 1,255 were pillar 2. Temperature was highest in summer while rainfall was generally lowest in spring and summer, but there were also periods of heavy rain in early and late summer (Figure 2a).

**Figure 2.**
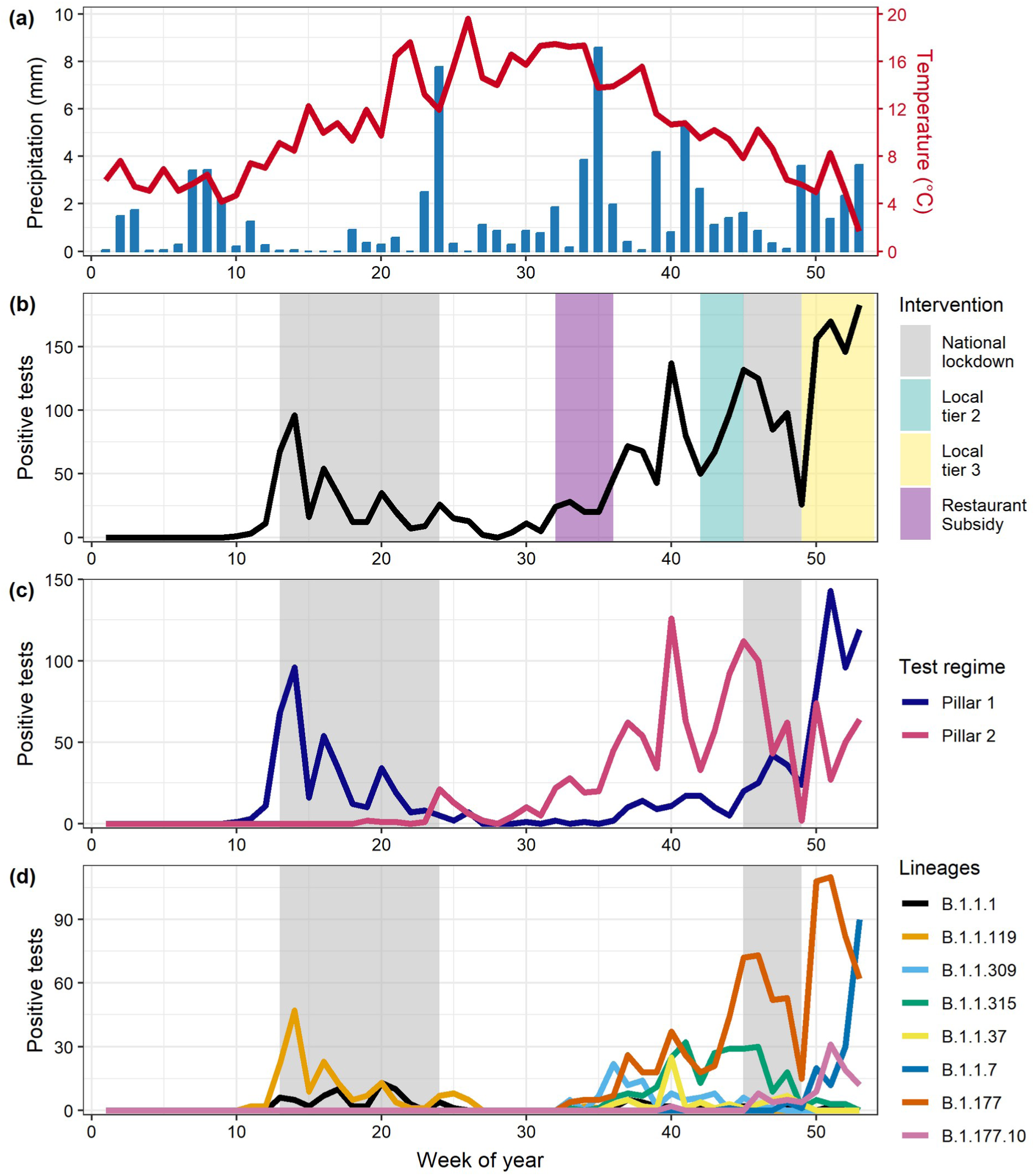
Trends in weather and positive tests of COVID-19 over the course of 2020, in Teesside. (a) Mean temperature and rainfall per week. (b) Total positive tests per week (across all lineages), coloured areas represent the periods of different interventions: national lockdown, weak local restrictions (Local tier 2), stricter local restrictions (Local tier 3), and the ‘Eat out to help out’ government subsidy to promote eating within restaurants. (c) Positive tests detected via the two testing regimes per week. (d) Positives tests of the eight most commonly detected lineages in Teesside per week.

The overall temporal trend in positive tests was bimodal with a first wave in spring (cases peaked in week 14, early April 2020) followed by a decline through the summer after introduction of the first lockdown, before the onset of the second wave of the epidemic in the early autumn (week 36) (Figure 2b). There was a large spike in cases around week 40 that were mainly detected via pillar 2 community testing (Figure 2c), which coincided with the start of school and university autumn terms in Teesside (Table 2). There was a strong positive correlation between the total number of cases of each lineage that were recorded by the two different pillars (rho=0.65, S=14017, df=60, P<0.001), suggesting that both pillars were reflecting similar patterns of infection across Teesside.

Out of 86 distinct lineages recorded in Teesside during 2020, only 8 were recorded more than 60 times: B.1.1.1, B.1.1.119, B.1.1.309, B.1.1.315, B.1.1.37, B.1.1.7, B.1.177 and B.1.177.10 (Figure 2d). These 8 lineages were recorded elsewhere in the UK before being recorded in Teesside. There were two dominant lineages in the first wave of the epidemic, B.1.1.1 and B.1.1.119, records for which declined to minimal levels during the late spring and summer. Patterns for other lineages were more complex in the second wave. B.1.177 and B.1.1.315 both increased in frequency during weeks 32 to 45 before declining during the second lockdown. However, after the end of this lockdown, B.1.177 increased substantially whilst B.1.1.315 did not. The more transmissible SARS-CoV-2 variant, B.1.1.7 (Alpha variant; VoC 202012/1), was first detected in the UK in December 2020 (Public Health England, 2021) and cases increased rapidly in Teesside after the end of the second national lockdown.

### Disease mapping

The relative risk of a positive test for COVID-19 in each Teesside postcode varied greatly amongst the lineages and between the two waves of the epidemic (Figure 3). The spatial pattern of risk of a positive test for any lineage was highest in some of the central areas, particularly in central Middlesbrough (40-60% higher in TS4), while the lowest risks were in the more rural eastern and western districts (20-60% lower). The spatial pattern does not follow a clear urban-rural divide however, as some urban and suburban areas had relative risks up to 20% lower than the overall risk across all districts (e.g., Thornaby (TS17), Redcar (TS10), and Eston (TS6)). During the first wave of the pandemic (lineages B.1.1.1 and B.1.1.119), risk was highest in the central areas of Teesside, particularly Middlesbrough (TS4, TS1, TS3, and TS8) and Guisborough (TS14), while northern and western districts had a very low risk. One of the second wave lineages (B.1.177) showed a more homogenous risk pattern, except for a raised risk in central Middlesbrough (TS4), though there is a high degree of uncertainty around this value (see Figures S1 and S2 in supplementary material for exceedance probabilities). The remaining two second wave lineages (B.1.1.7 and B.1.177.10) are those that occurred mainly towards the end of 2020; the spatial risk pattern for these lineages is less uniform and more focussed in northern districts with the highest risks in Hartlepool (TS24, TS25, and TS26), Trimdon (TS29), and Thornaby (TS17), with relative risk increase likely exceeding 50% for Hartlepool (Figures S1, S2).

**Figure 3.**
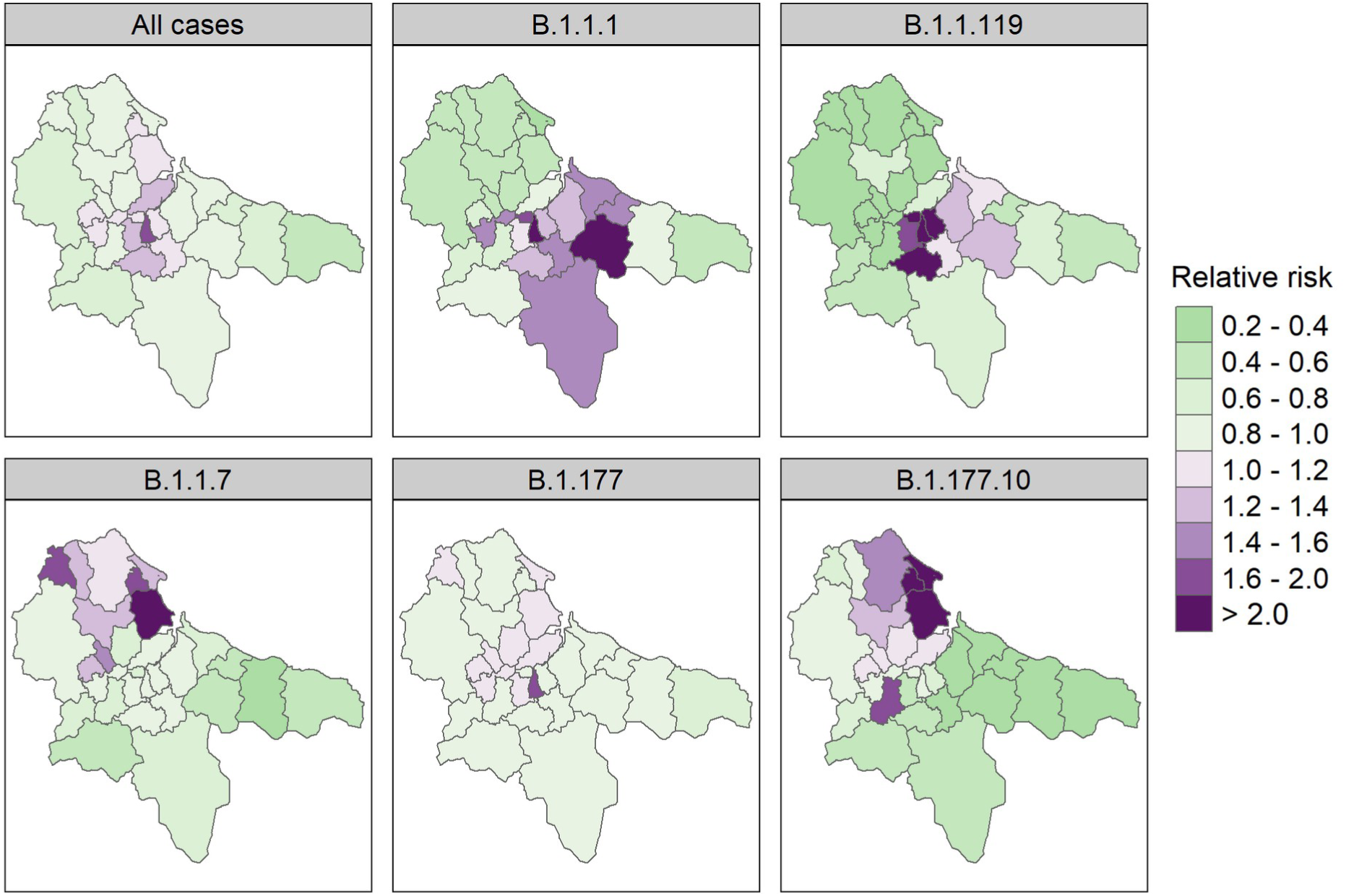
Area-specific relative risk of a positive test for any SARS-CoV-2 lineage (“All cases”), and 5 of the most common lineages, in each Teesside postcode district during 2020 (from the spatial CAR models). Maps for 3 of the lineages are not shown due to poor model fit (B.1.1.309, B.1.1.315, and B.1.1.37). Values are a ratio: green colours indicate a risk of infection that is lower than the overall risk for the entire study area (< 1), while purple colours indicate a higher risk (>1), and colour intensity indicates the strength of this effect. See Figures S1 and S2 in Supplementary Material for corresponding exceedance probabilities.

### Mixed-effect modelling

Most of the GLMMs had one fixed effect covariate that was identified as redundant and had a VIF value over 3 (the all-cases, B.1.1.1, and B.1.1.119 models did not), so the models reported here are the simplified ones where these fixed effects have been dropped (Figure 4). See Supplementary Materials for model output from both the full and final (simplified) versions (the coefficients from the full models are accurate in terms of the magnitude and direction of any effects, but the errors are unreliable). This collinearity was between the temporal variables and was most often strongest for the weather variables, though the collinearity was also very strong for the interventions or subsidy where lineages had a very narrow temporal window (e.g. B.1.1.7). The model for lineage B.1.1.309 was refit without the postcode random intercept as it had a variance that was almost 0 and the model showed temporal autocorrelation, which was controlled upon refitting.

**Figure 4.**
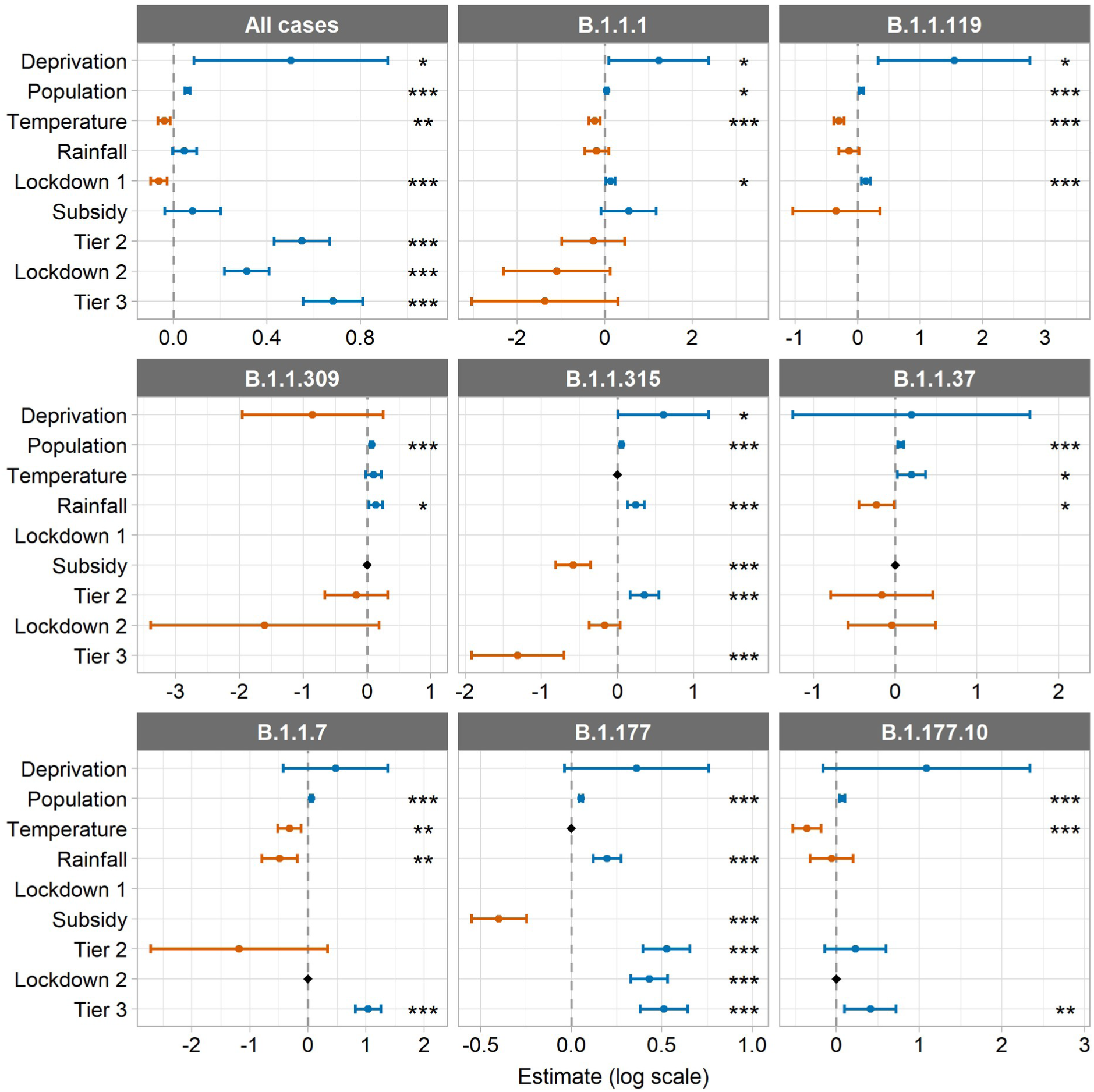
Fixed effects estimates and 95% confidence intervals from the final versions of the GLMMs with an AR1 term for week for each postcode examining all cases and the 8 most common lineages in Teesside (see Tables S3 and S6 for values). All models include a random intercept for postcode, except where the model fitted better without it (B.1.1.309). Black diamonds indicate variables that were dropped from the models due to VIF values > 3 (see Table S7 for summary values from the full models), remaining absent values represent non-overlap between some of the lineages and temporal variables (see Figure 2). Display order of fixed effects is as follows: spatial variables, continuous temporal variables, transient temporal variables (in chronological order of imposition). Estimates are on the original model scale (log). Asterisks indicate significance level: ‘***’ indicates p ≤ 0.001, ‘**’ indicates p ≤ 0.01, and ‘*’ indicates p ≤ 0.05. Note that each panel has a different y axis scale.

There were significant negative relationships between total positive tests of COVID-19 and mean weekly temperature and the first lockdown (Figure 4, Table S3). There were significant positive relationships between cases and total population, socio-economic deprivation (proportion of the population in the most deprived IMD decile), local tier 2, the second national lockdown, and tier 3. Mean weekly rainfall and the restaurant subsidy had no significant effect. The rate of increase in the total number of cases over time relative to the mean, was slower in many of the areas that experienced higher case numbers during the first wave, and faster in many areas that experienced more records during the second wave (Figures 5, 3 and S4, Table S5, see Figure S3 for observed records per week per postcode).

**Figure 5.**
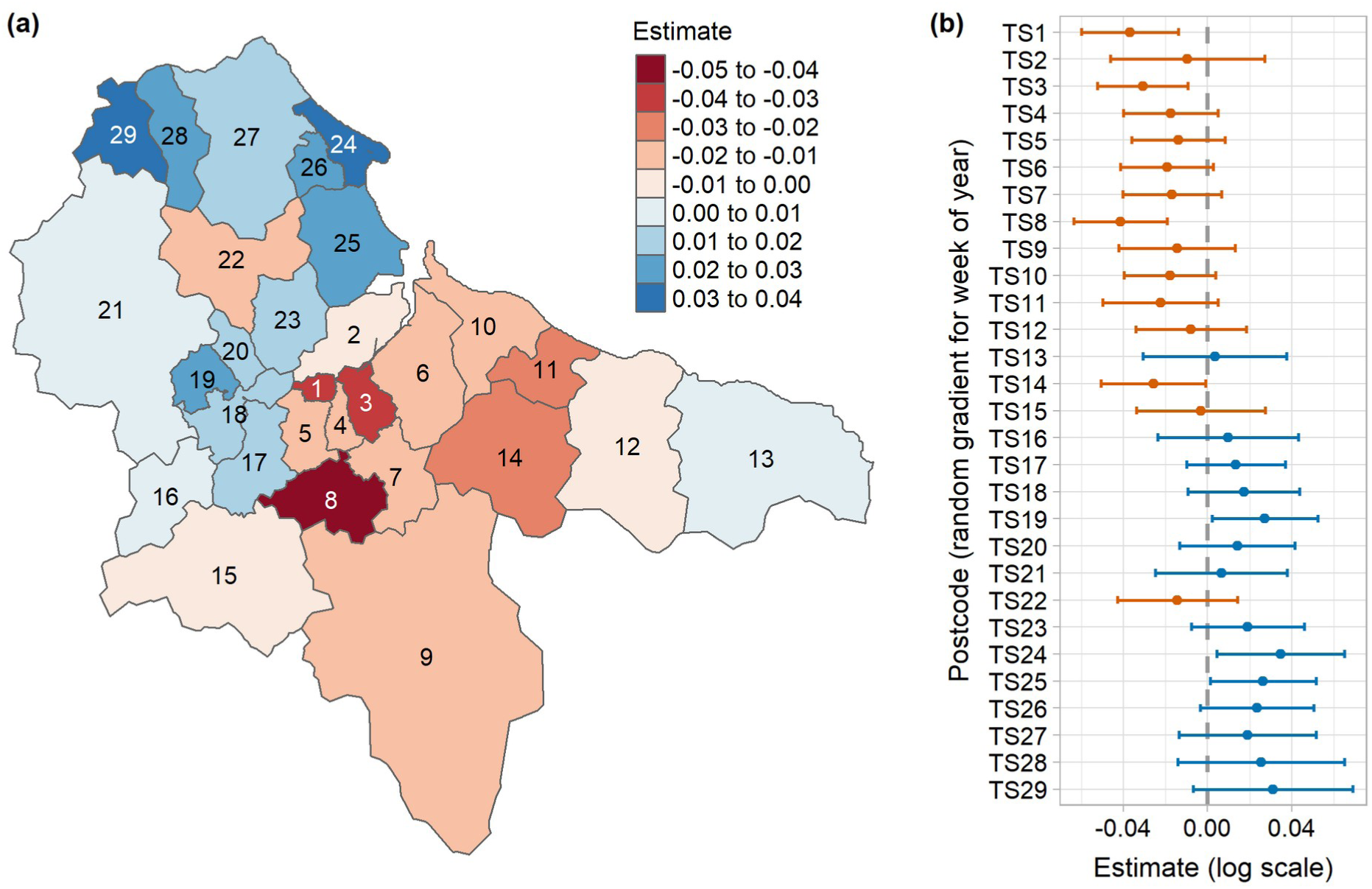
Random gradient estimates from the all-cases GLMM that included a random gradient for week for each postcode district, showing (a) the estimates on a map, and (b) the estimates with their 95% confidence intervals. Values represent the rate of change in the number of cases over time relative to the mean, where blue values are higher than the mean and a faster rate of increase (a steeper and more curved gradient), and red values are lower and a slower rate of increase (a shallower and less curved gradient) (see Figure S4 for predicted rate of change curves for each postcode district from the same model). Estimates are on the original model scale (log) (see Table S5 for full random effect summary values).

The relationships between cases and covariates for the two first wave lineages (B.1.1.1 and B.1.1.119) were similar, with a significant negative relationship for temperature, and significant positive relationships for deprivation, population, and the first lockdown (Figure 4, Table S6). Rainfall, the restaurant subsidy, tier 2, tier 3, and the second lockdown had no effect.

The patterns for the second wave lineages are less uniform, the only consistent relationship was a significant positive association with population in all models (Figure 4, Table S6). The only significant association with deprivation was a positive one with cases of B.1.1.315. Temperature had a significant negative relationship with cases of B.1.1.7 and B.1.177.10, and a positive relationship with cases of B.1.1.37. There were significant negative relationships between rainfall and cases of B.1.1.37 and B.1.1.7, and positive relationships with cases of B.1.1.309, B.1.1.315, and B.1.177. Cases were significantly decreased in relation to the restaurant subsidy for B.1.1.315 and B.1.177. There were significant positive relationships between cases and tier 2 for B.1.1.315 and B.1.177, and cases and lockdown 2 for B.1.177. Tier 3 was significantly negatively associated with cases of B.1.1.315, and positively associated with cases of B.1.1.7, B.1.177, and B.1.177.10.

Finally, the second-wave lineages model showed no relationship between the total number of cases of the two first wave lineages (B.1.1.1 and B.1.1.119) and the total number of cases of the six second wave lineages (B.1.1.309, B.1.1.315, B.1.1.37, B.1.1.7, B.1.177, and B.1.177.10) (see Table S8 for model output).

## Discussion

Our analyses indicate there was considerable spatial and temporal variation in occurrence of SARS-CoV-2 in Teesside during 2020 and that the patterns were related to demographic features, weather, and government interventions. These results must be interpreted with caution due to several limitations. Firstly, positive test ascertainment differed between the two epidemic waves during 2020. Community testing (pillar 2) was introduced later on in the first wave and did not become widespread until later in the year, which means that testing during the first wave will be biased towards severely ill individuals and healthcare workers. While the positive correlation between the number of pillar 1 vs pillar 2 tests for each lineage suggests that this bias did not affect the detection of different lineages, it is possible that it could have contributed to differences in the covariate associations that we found between the two waves. It is also possible that the timing of and access to tests could have differed between the two pillars, however, we believe the availability and promotion of local testing facilities and free at-home postal test kits, and the fact that both pillars included asymptomatic and symptomatic individuals, should minimise any related bias. Secondly, the resolution of the data aggregation may have been too coarse to detect some relationships. Aggregation was essential to ensure anonymity of records. However, the spatial resolution of postcode districts and the temporal resolution of one week, may have been too coarse to capture the spatial and temporal variation in the underlying epidemiological processes of transmission or other social and environmental drivers. Thirdly, collinearity between temporal variables was present in most of the separate lineage GLMM models. While we were able to assess and correct for this using a systematic approach, our results must be viewed in the context of the variables that were dropped. It is possible that some of the effects we see in our simplified models are actually being driven by those variables that were not included. And finally, many covariates associated with disease transmission, including proximity to infectious individuals and social contacts, could not be measured. We also did not have access to mobility data of a sufficient spatial resolution to incorporate into our models (due to cost). The covariates used in our models were therefore surrogates for the underlying mechanisms associated with disease transmission and spread. However, this is less of an issue in the context of our study, as we are more interested in highlighting the overall impact on cases in Teesside in relation to the local and national restriction policies (and other covariates), rather than the specific mechanisms that may be driving these relationships.

The spatial variation in total positive SARS-CoV-2 tests across the Teesside area was influenced by demographic factors, as we found the number of positive tests was increased in postcode districts with a higher population and those with higher levels of socio-economic deprivation. These are logical outcomes as transmission is more likely to occur in more densely-populated areas and amongst people who have higher exposure due to employment and living conditions (Niedzwiedz et al., 2020; Whittle & Diaz-Artiles, 2020; McGowan & Bambra, 2022). Our model output maps, when viewed in combination with the demographic information and regional knowledge, did not demonstrate a clear pattern of risk of, or rate of increase in, positive tests in relation to how urban or rural the postcode districts are. Research conducted on pillar 1 testing during the first wave in the East of England region found higher risks of infection in more urban areas (Brainard et al., 2022). Because we did not formally investigate land use in any of our models, we do not know what the true effect was, though we can speculate as to why we did not detect any sort of clear signal. It is possible that the size of our spatial units were too large and heterogenous in terms of their social environments to be able to detect a relationship like that found by Brainard et al. (2022), however, it is also possible that our longer timescale and inclusion of community testing collected a more representative sample of positive tests, or that this relationship does not hold in more urban and deprived areas like Teesside.

Our findings indicate that some weather variables affected the total positive tests across Teesside in 2020. We found positive tests were reduced by higher temperatures, in accordance with previous research (Ganslmeier et al., 2021; Alaniz et al., 2023; Nottmeyer et al., 2023). This is probably due to both indirect and direct mechanisms, as warmer weather encourages outdoor (rather than indoor) socialising and increased indoor ventilation, while also reducing transmissibility of the virus particles (Ganslmeier et al., 2021). However, we did not find an effect of rainfall on positive tests, and while some research has also found no significant associations (Ganslmeier et al., 2021), other studies have found a relationship (Tan & Schultz, 2022). Evidence from studies examining hourly rainfall and temperature data indicates that the effects of weather variables on positive tests are stronger during periods that facilitate transmission, such as mealtimes and when there are low/no restrictions on movement and socialising (Ganslmeier et al., 2021; Fetzer, 2022). Therefore, a finer temporal resolution than was used in this study is required to clarify the links between weather, transmission, and cases. It is also likely that the temporal resolution we used was too coarse to detect an impact of the government restaurant subsidy on cases, as the effects would have been limited to days when the subsidy applied (Monday-Wednesday) and mealtimes. Other research has found that cases were increased by the subsidy, but this effect was weaker when high rainfall coincided with mealtimes (Fetzer, 2022).

The government interventions introduced to manage COVID-19 cases during 2020 had mixed impacts on the total number of positive tests in Teesside. The first lockdown was successful in reducing cases, while all of the later interventions appeared to have the opposite effect. The only difference in restrictions between the two national lockdowns was that the first included school closures, whereas the second did not. A large body of evidence has demonstrated that school closures were one of the most important interventions in controlling COVID-19 (Brauner et al., 2021; Davies et al., 2021; Hunter et al., 2021; Ge et al., 2022; Torres et al., 2022). The length of the second lockdown was also shorter than the first one, though in Teesside it was immediately followed by tier 3 restrictions, which only differed in terms of the reopening of gyms, retail, and personal care non-essential businesses. The benefits of closing such business is unclear, with some studies showing modest or no benefits (Brauner et al., 2021; Hunter et al., 2021), while others show considerable benefits (Sharma et al., 2021), which makes it difficult to infer whether the length of the second lockdown related to its apparent failure. However, it does seem likely that school closures were an important factor of the success of the first lockdown relative to the failure of the second. The apparent positive effect of the second lockdown and the tier 2 and 3 restrictions could be due to the timing of imposition coinciding with one or more events that are epidemiologically important, such as the introduction of more transmissible lineages of the virus into the region, like B.1.177 and B.1.1.7 (Volz et al., 2021; Hinch et al., 2022; Panovska-Griffiths et al., 2022); or temporary changes in social behaviour or movement that facilitated transmission, like the reopening of retail businesses during the Christmas season. Other research has also found no effect or positive effects of non-pharmaceutical interventions on case counts, and it has been suggested that this could represent an association with increases in testing capacity or with changes in testing policy (Giudici et al., 2023; Lison et al., 2023). However, this is effect will be minimised in our dataset (total sequenced PCR tests rather than total positive PCR and/or antigen tests) by the sequencing capacity of the COG-UK consortium, which did not increase at the same rate as testing capacity in 2020. It is also possible that the difference in effectiveness could reflect differences in exposure between lockdowns due to changes in behaviour or routine, e.g. increased use of public transport during the second wave. People experiencing greater socio-economic deprivation in the UK have been shown to experience increased exposure to high infection risk activities permitted during the lockdowns, and that this varied slightly over time between different lockdowns/restrictions (Beale et al., 2022). Further research is needed to understand the factors affecting lockdown success in different communities, particularly ones with high levels of deprivation, such as Teesside.

Our findings suggest that the local tier system of interventions was less effective at reducing cases than a long and strict national lockdown, which has also been found in other studies (Davies et al., 2021; Torres et al., 2022). We also found that the tier restrictions were equally ineffective as the second national lockdown, all of which were applied during the second wave. This suggests that if there are any benefits to applying local-scale interventions in response to local-scale cases (rather than cases on the national scale, which in this context were determined by more populous and distant regions), they are masked by the effects of other factors, such as stringency and duration of restrictions, introduction events, and transmissibility of present lineages. Another possibility is that the tiered restrictions were not followed as rigorously as the national restrictions, either intentionally due to “pandemic fatigue” or accidentally due to poor communication (Smith et al., 2022; Delussu et al., 2022), especially as the tier levels in the UK could change at short notice and were not applied consistently across locations. While the tier levels were consistent within Teesside during their periods of imposition, communication of the restrictions was still unclear and complex as the tier levels were applied slightly differently in different parts of England and the information was usually published in long lists.

The analyses of positive tests of the eight most common SARS-CoV-2 lineages in Teesside demonstrated different spatial and temporal relationships between the lineages, which also differed to those of total positive tests, with differences in both magnitude and direction. Some of this disparity could be explained by the relatively small sample sizes of the lineage models; for example, the effect of socio-economic deprivation was significant for three lineages (B.1.1.1, B.1.1.119, B.1.1.315), but the confidence intervals for this variable were very large in all models. It seems likely that most of the differences between the total cases model and the separate lineages models are caused by the restricted spatial or temporal presence of the individual lineages when compared to all cases. For example, positive tests of lineage B.1.1.37 were positively related to temperature, but case numbers were generally very low and the only peak in cases occurred two weeks after a peak in temperature. While it is likely that some of the differences we observed in the spatial and temporal patterns between the different lineages are due to differences in transmissibility, particularly for B.1.1.7 (the alpha variant) (Volz et al., 2021; Hinch et al., 2022; Panovska-Griffiths et al., 2022), differences in the timing, location, and number of introductions are also likely to be a factor (Vöhringer et al., 2021). While the utility of examining covariate relationships for individual lineages over small geographic areas (with low numbers of sequenced tests) may appear to be low, the use of genomic sequencing to track community spread over such local scales holds great potential (du Plessis et al., 2021). Such forensic tracking could be used to further increase epidemiological understanding and perhaps target local interventions more effectively.

It has been well documented by large-scale studies that previous infections can confer natural immunity to subsequent infections (Flacco et al., 2022; Hall et al., 2022; Murugesan et al., 2022). Therefore, it is possible that natural immunity could explain some of the different spatial and temporal patterns between lineages in our data, particularly as the relative risk and the rate of increase in cases during the second wave tended to be higher in areas that had low relative risk and case numbers during the first wave. However, when we included the number of cases of first-wave lineages as a fixed effect in a GLMM modelling the number of cases of second-wave lineages, there was no apparent relationship. It seems likely that we were unable to detect a natural immunity effect because of the low numbers of sequenced positive tests in our study. This suggests that natural immunity, and possibly other disease processes, are easier to detect at larger spatial scales and with larger datasets.

Our study has demonstrated the effects of weather and government interventions on the number of SARS-CoV-2 positive tests at a sub-regional scale in Teesside, UK. The number of COVID-19 cases was negatively related to temperature and the first national lockdown. There were positive relationships between cases and total population, socio-economic deprivation, the second lockdown, and local tiered restrictions. While further research is needed to investigate the factors affecting lockdown success in different communities, we feel confident to make several recommendations regarding future epidemic policy responses in local/regional contexts, based on both ours and other’s findings. Firstly, school closures are one of the most important interventions in controlling transmission and mortality (Brauner et al., 2021; Davies et al., 2021; Hunter et al., 2021; Ge et al., 2022; Torres et al., 2022), therefore school closures should be included in national and local/regional lockdowns. Secondly, interventions applied at the local/regional scale are less effective if they are less strict or applied later (Davies et al., 2021; Torres et al., 2022), therefore all tier levels (not just the highest) should be stringent and they should be imposed early. It is also imperative that local restrictions are communicated clearly and effectively with the public, and that the rules are simple and consistent across areas, so as to facilitate adherence (Smith et al., 2022). Thirdly, transmission at the regional scale is dependent on introductions (da Silva Filipe et al., 2021; du Plessis et al., 2021; Lane et al., 2021; Vöhringer et al., 2021), particularly during periods of low restrictions (Lemey et al., 2021), so long-distance domestic and international travel restrictions should be imposed quickly at the start of an epidemic and the latter maintained. And finally, regional and local transmission is dependent on both the transmissibility, location, and number of introductions of different lineages (da Silva Filipe et al., 2021; du Plessis et al., 2021; Lane et al., 2021; Vöhringer et al., 2021), therefore the possibility of using genomic sequencing to conduct forensic tracking of community spread should be investigated.

## Data Availability

All data produced in the present study are available upon reasonable request to the authors

https://doi.org/10.25405/data.ncl.23815077

## Acknowledgements

The authors thank colleagues within COG-UK and three anonymous reviewers for constructive comments on earlier versions of this manuscript.

## Data, scripts, code, and supplementary information availability

Data, scripts, code, and supplementary information are available online: https://doi.org/10.25405/data.ncl.23815077

## Ethics approval

The surveillance activities within which this study was conducted are part of Public Health England’s responsibility to monitor COVID-19 during the current pandemic. Public Health England has legal permission, provided by Regulation 3 of The Health Service (Control of Patient Information) Regulations 2002 to process confidential patient information under Sections 3(i) a–c, 3(i) d(i and ii), and 3(iii) as part of its outbreak response activities. This study falls within the research activities approved by the Public Health England Research Ethics and Governance of Public Health Practice Group. Informed consent was obtained from all individual participants included in the study. No individual patient data in any form is included in this manuscript.

