## Supplementary Tables and Figures for "Spatial and temporal epidemiology of SARS-CoV-2 virus lineages in Teesside, UK, in 2020: effects of socio-economic deprivation, weather, and lockdown on lineage dynamics"

### 1. Supplementary Results Figures and Tables

#### 1.1 Data and Trends

The first recorded cases of COVID-19 in the UK were in January 2020. After cases rose rapidly in March, a complete social and economic lockdown was introduced in England on 23/03/2020 (UK Government, 2020a), which led to a swift reduction in cases (Covid-19 control measures differed in Scotland, Wales, and Northern Ireland and were decided by their own devolved parliaments). A second major outbreak in autumn 2020 led to the introduction of further interventions in England: a heterogenous “tier” system applied at local scales (UK Government, 2020c), and a short national lockdown in November 2020 (UK Government, 2020d). When these failed to reduce hospital admissions and deaths, a further national lockdown was introduced in January 2021 (UK Government, 2021).

**Table S1:** Total resident population and proportion of the population within the 10$^{th}$ decile of the Index of Multiple Deprivation (when assessed at the scale of England as a whole), for each Teesside postcode district.

| Postcode district | Population | Proportion 10th Decile |
|---|---|---|
| TS1 | 16994 | 0.997 |
| TS2 | 780 | 1 |
| TS3 | 29253 | 0.949 |
| TS4 | 16966 | 0.789 |
| TS5 | 39387 | 0.133 |
| TS6 | 30239 | 0.410 |
| TS7 | 22058 | 0.141 |
| TS8 | 21266 | 0.204 |
| TS9 | 12832 | 0 |
| TS10 | 36800 | 0.303 |
| TS11 | 11822 | 0 |
| TS12 | 22491 | 0.070 |
| TS13 | 9975 | 0.160 |
| TS14 | 17548 | 0.077 |
| TS15 | 12891 | 0 |
| TS16 | 10422 | 0 |
| TS17 | 44889 | 0.163 |
| TS18 | 24163 | 0.422 |
| TS19 | 37087 | 0.380 |
| TS20 | 20663 | 0.229 |
| TS21 | 12472 | 0.004 |
| TS22 | 8662 | 0 |
| TS23 | 30241 | 0.120 |
| TS24 | 22019 | 0.630 |
| TS25 | 38616 | 0.392 |
| TS26 | 25253 | 0.204 |
| TS27 | 12193 | 0.207 |
| TS28 | 5318 | 0.001 |
| TS29 | 6300 | 0.002 |

#### 1.2 Disease Mapping

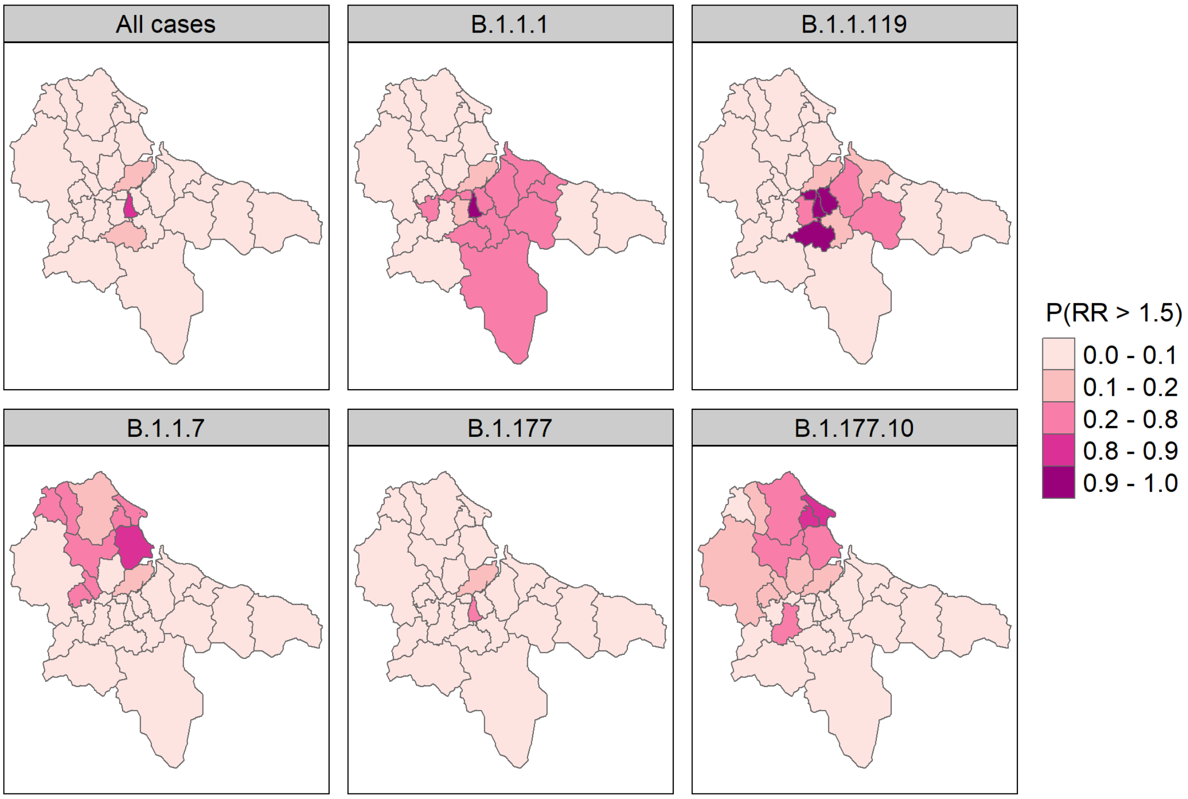

**Figure S1**: Map of the exceedance probabilities for relative risk threshold of 1.5. Relative risk is very likely to exceed 1.5 when probabilities are close to 1, and very unlikely when values are close to 0 (values around 0.5 have the highest uncertainty). Maps for 3 of the lineages are not shown due to poor model fit (B.1.1.309, B.1.1.315, and B.1.1.37).

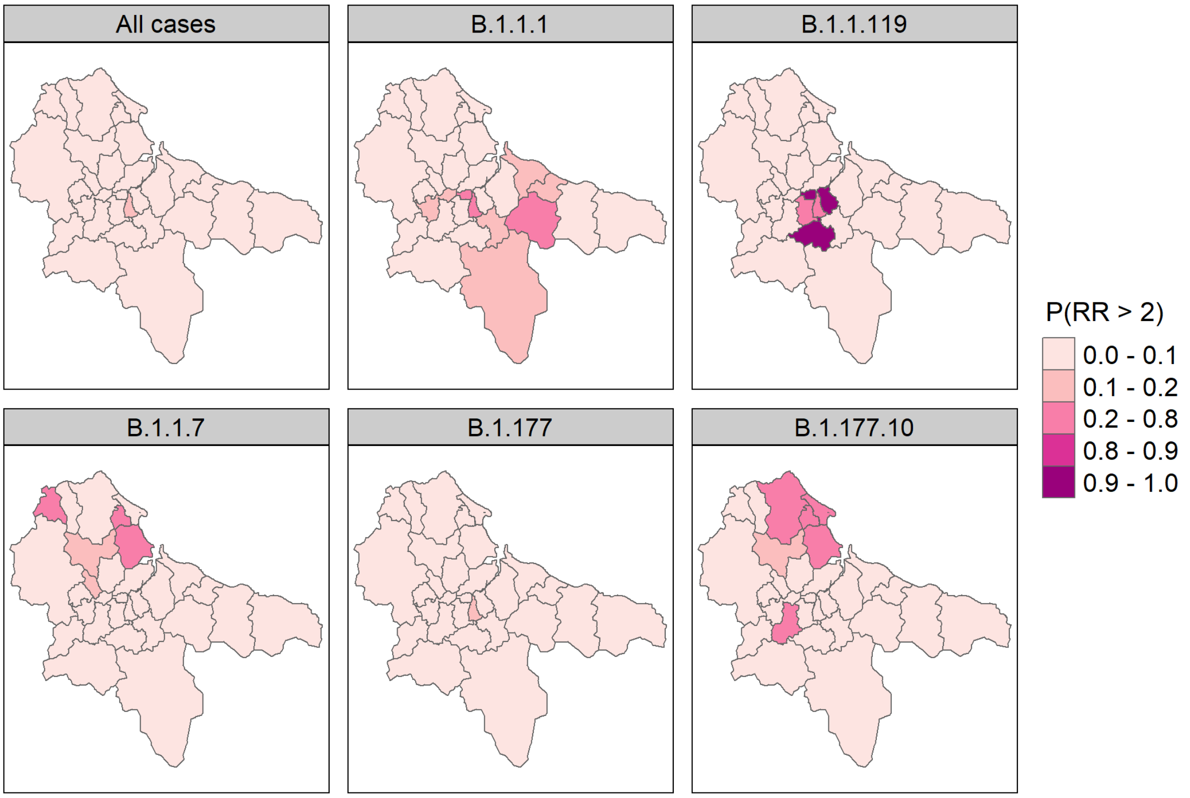

**Figure S2**: Map of the exceedance probabilities for relative risk threshold of 2. Relative risk is very likely to exceed 2 when probabilities are close to 1, and very unlikely when values are close to 0 (values around 0.5 have the highest uncertainty). Maps for 3 of the lineages are not shown due to poor model fit (B.1.1.309, B.1.1.315, and B.1.1.37).

**Table S2:** Mean, standard deviation (SD), and upper and lower limits of the 95% credible intervals (0.025 quant and 0.975 quant) of the posterior distributions of the fixed and random effects from the INLA disease mapping models. Output for 3 of the lineages are not shown due to poor model fit (B.1.1.309, B.1.1.315, and B.1.1.37).

| Lineage | Effect | Effect type | Mean | SD | 0.025 quant | 0.975 quant |
|---|---|---|---|---|---|---|
| Total cases | Intercept | Fixed | -0.073 | 0.044 | -0.159 | 0.015 |
| Total cases | Size for the nbinomial observations | Random | 1911.5 | 12911.2 | 32.051 | 12753.9 |
| Total cases | Precision for postcode | Random | 14.178 | 5.445 | 6.056 | 27.187 |
| Total cases | Phi for postcode | Random | 0.751 | 0.225 | 0.200 | 0.994 |
| B.1.1.1 | Intercept | Fixed | -0.287 | 0.183 | -0.673 | 0.049 |
| B.1.1.1 | Size for the nbinomial observations | Random | 903.7 | 9172.4 | 5.229 | 5869.4 |
| B.1.1.1 | Precision for postcode | Random | 2.984 | 1.694 | 0.962 | 7.381 |
| B.1.1.1 | Phi for postcode | Random | 0.631 | 0.242 | 0.138 | 0.974 |
| B.1.1.119 | Intercept | Fixed | -0.581 | 0.186 | -0.967 | -0.231 |
| B.1.1.119 | Size for the nbinomial observations | Random | 881.5 | 8916.5 | 6.237 | 5736.4 |
| B.1.1.119 | Precision for postcode | Random | 1.313 | 0.477 | 0.617 | 2.468 |
| B.1.1.119 | Phi for postcode | Random | 0.620 | 0.232 | 0.155 | 0.965 |
| B.1.1.7 | Intercept | Fixed | -0.201 | 0.137 | -0.487 | 0.052 |
| B.1.1.7 | Size for the nbinomial observations | Random | 1723.1 | 23154.0 | 9.132 | 10512.1 |
| B.1.1.7 | Precision for postcode | Random | 4.512 | 2.505 | 1.406 | 10.944 |
| B.1.1.7 | Phi for postcode | Random | 0.498 | 0.254 | 0.069 | 0.936 |
| B.1.177 | Intercept | Fixed | -0.032 | 0.061 | -0.154 | 0.087 |
| B.1.177 | Size for the nbinomial observations | Random | 315.9 | 895.2 | 18.129 | 1890.2 |
| B.1.177 | Precision for postcode | Random | 18.059 | 9.356 | 5.221 | 41.174 |
| B.1.177 | Phi for postcode | Random | 0.304 | 0.230 | 0.016 | 0.821 |
| B.1.177.10 | Intercept | Fixed | -0.432 | 0.197 | -0.849 | -0.076 |
| B.1.177.10 | Size for the nbinomial observations | Random | 589.8 | 4605.9 | 3.602 | 3957.5 |
| B.1.177.10 | Precision for postcode | Random | 2.660 | 1.382 | 0.916 | 6.194 |
| B.1.177.10 | Phi for postcode | Random | 0.635 | 0.231 | 0.159 | 0.968 |

#### 1.3 Mixed-effect Modelling

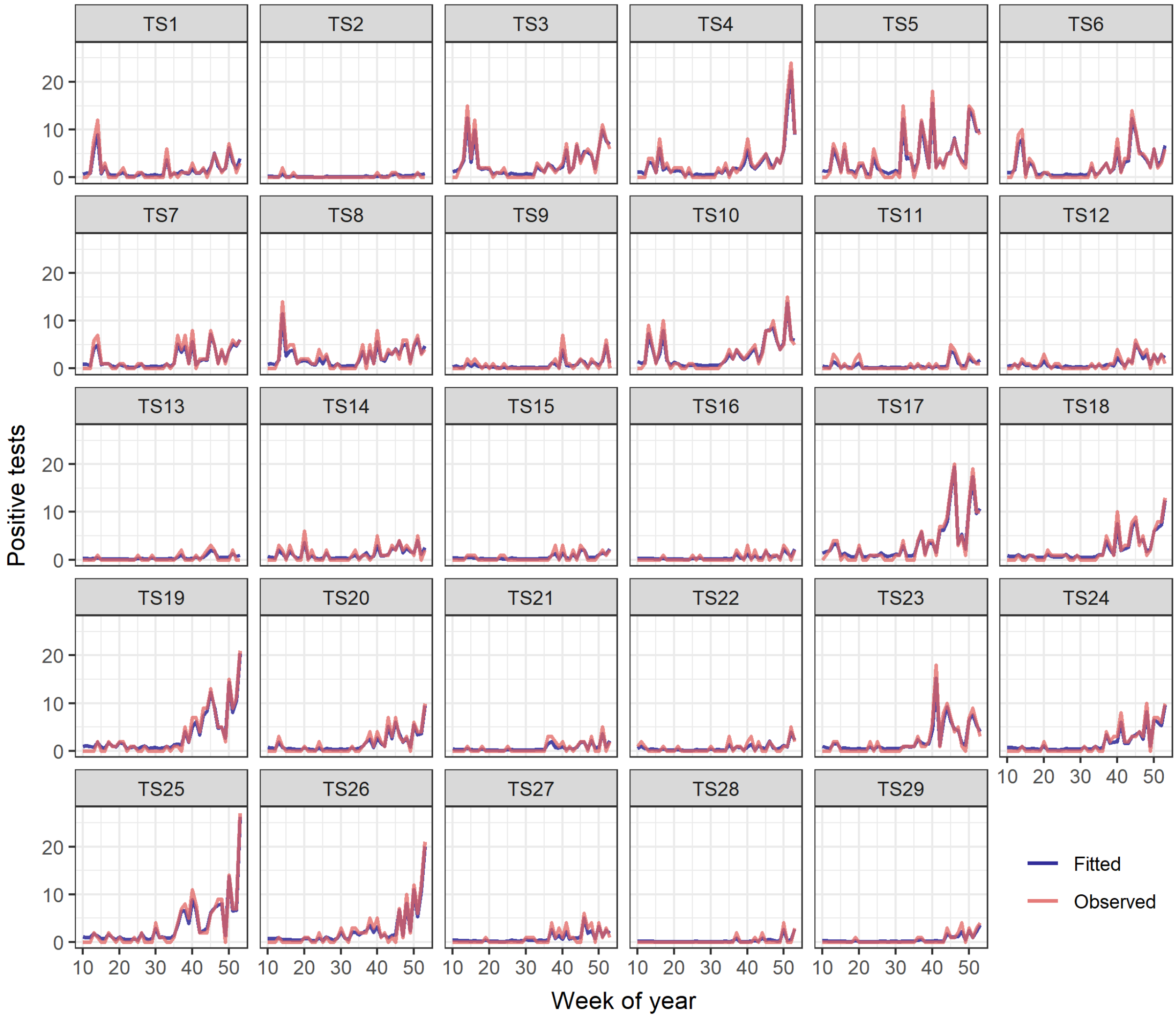

**Figure S3**: Predicted and observed total number of positive cases of COVID-19 (across all lineages) in Teesside during 2020. The predicted estimates (blue) are fitted values from the GLMM for all cases with an AR1 (autoregressive order 1) term for week for each postcode.

**Table S3**: Summary of the all-cases GLMM with an AR1 term for week for each postcode; effects of spatial (10th Decile of IMD, postcode district total population) and temporal variables (mean weekly temperature and rainfall, and government interventions and subsidy) on the total number of positive PCR tests recorded each week of 2020 in each postcode district. All temporal variables include a two-week time lag to account for the delay in symptom onset (and testing) after infection. VIF values for all variables were < 3. Estimates are on the original model scale (log).

| Variable | Estimate | Std. Error | z value | p value |
|---|---|---|---|---|
| Intercept | -1.479 | 0.236 | -6.276 | <0.001 |
| Temperature | -0.039 | 0.014 | -2.907 | 0.004 |
| Rainfall | 0.047 | 0.026 | 1.789 | 0.074 |
| Lockdown 1 | -0.063 | 0.018 | -3.486 | <0.001 |
| Lockdown 2 | 0.314 | 0.049 | 6.411 | <0.001 |
| Tier 2 | 0.551 | 0.061 | 9.079 | <0.001 |
| Tier 3 | 0.683 | 0.064 | 10.589 | <0.001 |
| Restaurant Subsidy | 0.082 | 0.061 | 1.340 | 0.180 |
| IMD 10th Decile | 0.502 | 0.211 | 2.380 | 0.017 |
| Total Population | 0.061 | 0.006 | 10.426 | <0.001 |

**Table S4**: Summary of the all-cases GLMM with a random gradient for week of year for each postcode; effects of spatial (10th Decile of IMD, postcode district total population) and temporal variables (mean weekly temperature and rainfall, and government interventions and subsidy) on the total number of positive PCR tests recorded each week of 2020 in each postcode district. All temporal variables include a two-week time lag to account for the delay in symptom onset (and testing) after infection. VIF values for all variables were < 3. Estimates are on the original model scale (log).

| Variable | Estimate | Std. Error | z value | p value |
|---|---|---|---|---|
| Intercept | -1.312 | 0.279 | -4.701 | <0.001 |
| Temperature | -0.121 | 0.014 | -8.421 | <0.001 |
| Rainfall | -0.089 | 0.028 | -3.230 | 0.001 |
| Lockdown 1 | -0.016 | 0.016 | -0.964 | 0.335 |
| Lockdown 2 | -0.243 | 0.061 | -3.990 | <0.001 |
| Week | 0.064 | 0.007 | 8.917 | <0.001 |
| Tier 2 | 0.040 | 0.066 | 0.605 | 0.545 |
| Tier 3 | -0.173 | 0.080 | -2.172 | 0.030 |
| Restaurant Subsidy | 0.258 | 0.059 | 4.380 | <0.001 |
| IMD 10th Decile | 0.348 | 0.226 | 1.536 | 0.124 |
| Total Population | 0.062 | 0.006 | 10.321 | <0.001 |

**Table S5**: Random effect estimates for the all-cases GLMM with a random gradient for week of year for each postcode. Estimates are on the original model scale (log).

| Postcode District | Estimate | Std. Error | 95% CI |
|---|---|---|---|
| TS1 | -0.037 | 0.012 | [-0.06, -0.014] |
| TS10 | -0.018 | 0.011 | [-0.04, 0.004] |
| TS11 | -0.023 | 0.014 | [-0.05, 0.005] |
| TS12 | -0.008 | 0.013 | [-0.034, 0.018] |
| TS13 | 0.003 | 0.017 | [-0.031, 0.037] |
| TS14 | -0.026 | 0.013 | [-0.051, -0.001] |
| TS15 | -0.003 | 0.016 | [-0.034, 0.027] |
| TS16 | 0.010 | 0.017 | [-0.024, 0.043] |
| TS17 | 0.013 | 0.012 | [-0.01, 0.037] |
| TS18 | 0.017 | 0.014 | [-0.009, 0.044] |
| TS19 | 0.027 | 0.013 | [0.002, 0.052] |
| TS2 | -0.010 | 0.019 | [-0.046, 0.027] |
| TS20 | 0.014 | 0.014 | [-0.013, 0.042] |
| TS21 | 0.006 | 0.016 | [-0.025, 0.038] |
| TS22 | -0.014 | 0.015 | [-0.043, 0.014] |
| TS23 | 0.019 | 0.014 | [-0.008, 0.046] |
| TS24 | 0.035 | 0.016 | [0.004, 0.065] |
| TS25 | 0.026 | 0.013 | [0.001, 0.052] |
| TS26 | 0.023 | 0.014 | [-0.004, 0.051] |
| TS27 | 0.019 | 0.017 | [-0.014, 0.052] |
| TS28 | 0.026 | 0.020 | [-0.014, 0.065] |
| TS29 | 0.031 | 0.019 | [-0.007, 0.069] |
| TS3 | -0.031 | 0.011 | [-0.052, -0.009] |
| TS4 | -0.018 | 0.011 | [-0.04, 0.005] |
| TS5 | -0.014 | 0.011 | [-0.036, 0.008] |
| TS6 | -0.019 | 0.011 | [-0.041, 0.003] |
| TS7 | -0.017 | 0.012 | [-0.04, 0.007] |
| TS8 | -0.042 | 0.011 | [-0.064, -0.019] |
| TS9 | -0.015 | 0.014 | [-0.042, 0.013] |

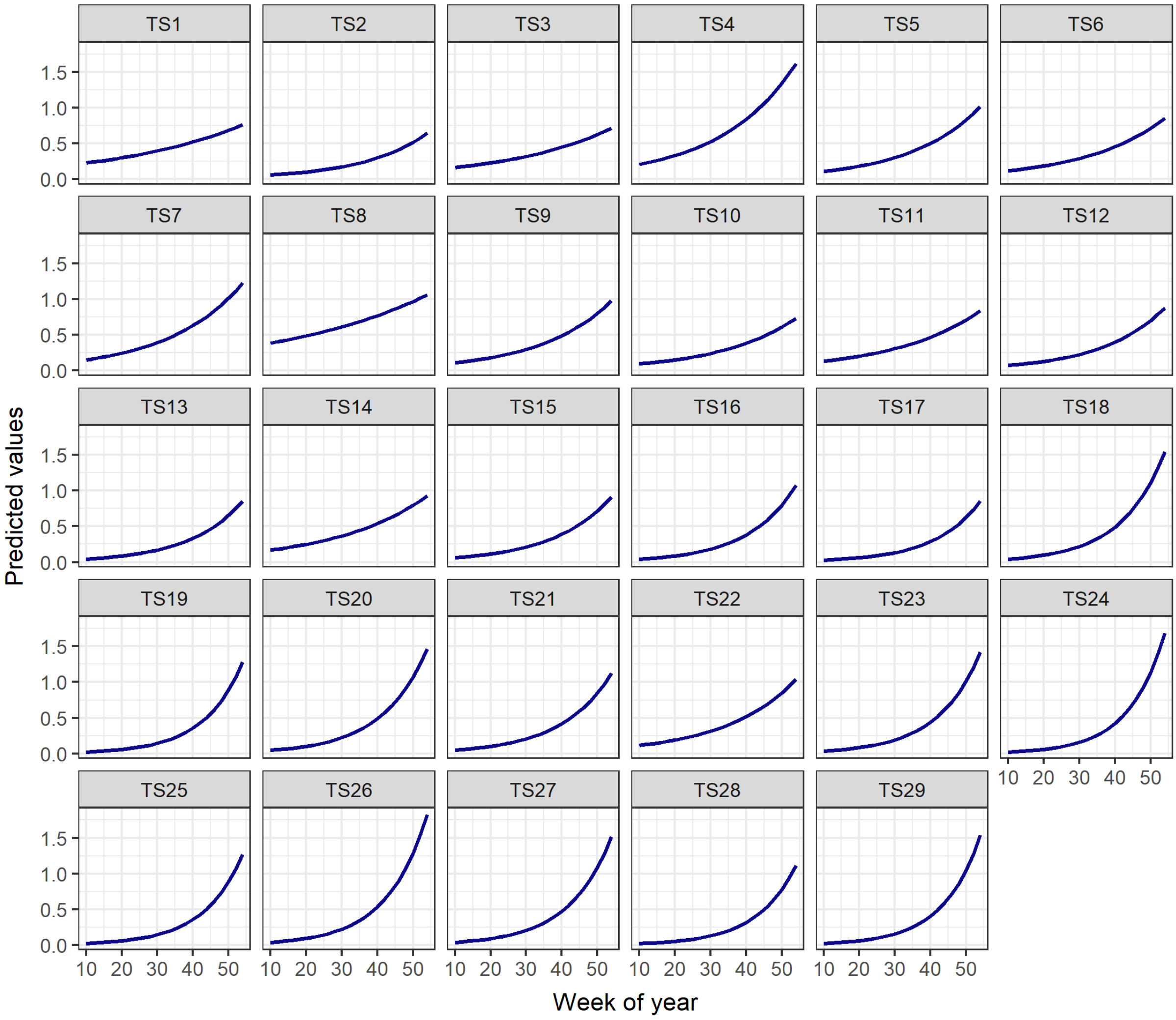

**Figure S4**: Rate of change in predicted positive tests of COVID-19 (across all lineages) in Teesside during 2020, from the GLMM with a random gradient for week for each postcode district. Predicted values represent the increase in cases over time relative to the mean, when all fixed effects are held constant, and only the random effects (week and postcode) are allowed to vary. Predicted values were calculated via the “ggeffects” R package.

**Table S6**: Summary of the final versions of the separate lineage GLMMs with an AR1 term for week for each postcode; effects of spatial (10th Decile of IMD, postcode district total population) and temporal variables (mean weekly temperature and rainfall, week of year, and government interventions and subsidy) on the total number of positive PCR tests recorded each week of 2020 in each postcode district, for each of the 8 most common lineages of SARS-CoV-2. All temporal variables include a two-week time lag to account for the delay in symptom onset (and testing) after infection. Variables with a VIF > 3 were removed from the models where present (B.1.1.309, B.1.1.315, B.1.1.37, B.1.1.7, B.1.177, B.1.177.10). Estimates are on the original model scale (log) (Continued on next page).

| Lineage | Variable | Estimate | Std. Error | z value | p value |
|---|---|---|---|---|---|
| B.1.1.1 | Intercept | -3.629 | 1.301 | -2.789 | 0.005 |
| B.1.1.1 | Temperature | -0.234 | 0.066 | -3.546 | <0.001 |
| B.1.1.1 | Rainfall | -0.184 | 0.140 | -1.319 | 0.187 |
| B.1.1.1 | Lockdown 1 | 0.131 | 0.057 | 2.286 | 0.022 |
| B.1.1.1 | Lockdown 2 | -1.097 | 0.622 | -1.763 | 0.078 |
| B.1.1.1 | Tier 2 | -0.260 | 0.365 | -0.711 | 0.477 |
| B.1.1.1 | Tier 3 | -1.371 | 0.852 | -1.609 | 0.108 |
| B.1.1.1 | Restaurant Subsidy | 0.550 | 0.321 | 1.713 | 0.087 |
| B.1.1.1 | IMD 10th Decile | 1.238 | 0.583 | 2.122 | 0.034 |
| B.1.1.1 | Total Population | 0.042 | 0.017 | 2.548 | 0.011 |
| B.1.1.119 | Intercept | -2.088 | 0.621 | -3.360 | <0.001 |
| B.1.1.119 | Temperature | -0.295 | 0.042 | -7.046 | <0.001 |
| B.1.1.119 | Rainfall | -0.134 | 0.082 | -1.633 | 0.103 |
| B.1.1.119 | Lockdown 1 | 0.131 | 0.037 | 3.509 | <0.001 |
| B.1.1.119 | Restaurant Subsidy | -0.337 | 0.356 | -0.947 | 0.343 |
| B.1.1.119 | IMD 10th Decile | 1.546 | 0.619 | 2.499 | 0.012 |
| B.1.1.119 | Total Population | 0.060 | 0.018 | 3.313 | <0.001 |
| B.1.1.309 | Intercept | -5.477 | 1.083 | -5.058 | <0.001 |
| B.1.1.309 | Temperature | 0.103 | 0.062 | 1.671 | 0.095 |
| B.1.1.309 | Rainfall | 0.140 | 0.055 | 2.540 | 0.011 |
| B.1.1.309 | Lockdown 2 | -1.605 | 0.913 | -1.757 | 0.079 |
| B.1.1.309 | Tier 2 | -0.170 | 0.252 | -0.677 | 0.498 |
| B.1.1.309 | IMD 10th Decile | -0.852 | 0.565 | -1.508 | 0.131 |
| B.1.1.309 | Total Population | 0.074 | 0.012 | 6.301 | <0.001 |
| B.1.1.315 | Intercept | -3.155 | 0.324 | -9.743 | <0.001 |
| B.1.1.315 | Rainfall | 0.243 | 0.056 | 4.329 | <0.001 |
| B.1.1.315 | Lockdown 2 | -0.165 | 0.104 | -1.578 | 0.115 |
| B.1.1.315 | Tier 2 | 0.357 | 0.096 | 3.725 | <0.001 |
| B.1.1.315 | Tier 3 | -1.307 | 0.309 | -4.227 | <0.001 |
| B.1.1.315 | Restaurant Subsidy | -0.580 | 0.116 | -5.020 | <0.001 |
| B.1.1.315 | IMD 10th Decile | 0.605 | 0.305 | 1.984 | 0.047 |
| B.1.1.315 | Total Population | 0.055 | 0.008 | 6.753 | <0.001 |
| B.1.1.37 | Intercept | -7.207 | 1.536 | -4.693 | <0.001 |
| B.1.1.37 | Temperature | 0.200 | 0.089 | 2.235 | 0.025 |
| B.1.1.37 | Rainfall | -0.228 | 0.109 | -2.089 | 0.037 |
| B.1.1.37 | Lockdown 2 | -0.039 | 0.272 | -0.144 | 0.885 |
| B.1.1.37 | Tier 2 | -0.162 | 0.319 | -0.507 | 0.612 |

| | | | | | |
|---|---|---|---|---|---|
| B.1.1.37 | IMD 10th Decile | 0.199 | 0.740 | 0.269 | 0.788 |
| B.1.1.37 | Total Population | 0.070 | 0.019 | 3.748 | <0.001 |
| B.1.1.7 | Intercept | -0.938 | 0.892 | -1.051 | 0.293 |
| B.1.1.7 | Temperature | -0.315 | 0.102 | -3.106 | 0.002 |
| B.1.1.7 | Rainfall | -0.485 | 0.157 | -3.083 | 0.002 |
| B.1.1.7 | Tier 2 | -1.187 | 0.778 | -1.525 | 0.127 |
| B.1.1.7 | Tier 3 | 1.035 | 0.113 | 9.136 | <0.001 |
| B.1.1.7 | IMD 10th Decile | 0.476 | 0.457 | 1.041 | 0.298 |
| B.1.1.7 | Total Population | 0.066 | 0.012 | 5.626 | <0.001 |
| B.1.177 | Intercept | -2.306 | 0.201 | -11.478 | <0.001 |
| B.1.177 | Rainfall | 0.198 | 0.039 | 5.030 | <0.001 |
| B.1.177 | Lockdown 2 | 0.430 | 0.052 | 8.263 | <0.001 |
| B.1.177 | Tier 2 | 0.525 | 0.066 | 7.947 | <0.001 |
| B.1.177 | Tier 3 | 0.510 | 0.067 | 7.631 | <0.001 |
| B.1.177 | Restaurant Subsidy | -0.399 | 0.077 | -5.164 | <0.001 |
| B.1.177 | IMD 10th Decile | 0.361 | 0.203 | 1.778 | 0.075 |
| B.1.177 | Total Population | 0.054 | 0.006 | 9.628 | <0.001 |
| B.1.177.10 | Intercept | -1.842 | 0.975 | -1.890 | 0.059 |
| B.1.177.10 | Temperature | -0.354 | 0.088 | -4.021 | <0.001 |
| B.1.177.10 | Rainfall | -0.056 | 0.133 | -0.423 | 0.672 |
| B.1.177.10 | Tier 2 | 0.231 | 0.189 | 1.223 | 0.221 |
| B.1.177.10 | Tier 3 | 0.409 | 0.158 | 2.591 | 0.010 |
| B.1.177.10 | IMD 10th Decile | 1.089 | 0.638 | 1.707 | 0.088 |
| B.1.177.10 | Total Population | 0.071 | 0.017 | 4.068 | <0.001 |

**Table S7**: Summary of the full versions of the separate lineage GLMMs with an AR1 term for week for each postcode; effects of spatial (10th Decile of IMD, postcode district total population) and temporal variables (mean weekly temperature and rainfall, week of year, and government interventions and subsidy) on the total number of positive PCR tests recorded each week of 2020 in each postcode district, for each of the 8 most common lineages of SARS-CoV-2. All temporal variables include a two-week time lag to account for the delay in symptom onset (and testing) after infection. The VIF of at least one variable in each model was > 3, which means all of the below standard errors, z values, and p values may be unreliable (though the estimates are accurate). Estimates are on the original model scale (log) (Continued on next page).

| Lineage | Variable | Estimate | Std. Error | z value | p value |
|---|---|---|---|---|---|
| B.1.1.309 | Intercept | -3.815 | 1.459 | -2.614 | 0.009 |
| B.1.1.309 | Temperature | -0.006 | 0.092 | -0.065 | 0.949 |
| B.1.1.309 | Rainfall | -0.002 | 0.103 | -0.022 | 0.983 |
| B.1.1.309 | Lockdown 2 | -2.012 | 1.010 | -1.991 | 0.047 |
| B.1.1.309 | Tier 2 | -0.347 | 0.273 | -1.270 | 0.204 |
| B.1.1.309 | Restaurant Subsidy | 0.299 | 0.188 | 1.588 | 0.112 |
| B.1.1.309 | IMD 10th Decile | -0.880 | 0.568 | -1.549 | 0.121 |
| B.1.1.309 | Total Population | 0.075 | 0.012 | 6.323 | <0.001 |
| B.1.1.315 | Intercept | -1.735 | 0.873 | -1.987 | 0.047 |
| B.1.1.315 | Temperature | -0.096 | 0.056 | -1.701 | 0.089 |
| B.1.1.315 | Rainfall | 0.156 | 0.075 | 2.091 | 0.037 |
| B.1.1.315 | Lockdown 2 | -0.377 | 0.162 | -2.323 | 0.020 |
| B.1.1.315 | Tier 2 | 0.205 | 0.129 | 1.587 | 0.113 |
| B.1.1.315 | Tier 3 | -1.719 | 0.412 | -4.176 | <0.001 |
| B.1.1.315 | Restaurant Subsidy | -0.404 | 0.154 | -2.619 | 0.009 |
| B.1.1.315 | IMD 10th Decile | 0.611 | 0.305 | 2.003 | 0.045 |
| B.1.1.315 | Total Population | 0.055 | 0.008 | 6.743 | <0.001 |
| B.1.1.37 | Intercept | -6.705 | 2.145 | -3.126 | 0.002 |
| B.1.1.37 | Temperature | 0.167 | 0.134 | 1.247 | 0.212 |
| B.1.1.37 | Rainfall | -0.287 | 0.213 | -1.343 | 0.179 |
| B.1.1.37 | Lockdown 2 | -0.107 | 0.341 | -0.314 | 0.754 |
| B.1.1.37 | Tier 2 | -0.200 | 0.338 | -0.592 | 0.554 |
| B.1.1.37 | Restaurant Subsidy | 0.130 | 0.404 | 0.322 | 0.748 |
| B.1.1.37 | IMD 10th Decile | 0.197 | 0.739 | 0.267 | 0.790 |
| B.1.1.37 | Total Population | 0.070 | 0.019 | 3.759 | <0.001 |
| B.1.1.7 | Intercept | -11.428 | 3.550 | -3.219 | 0.001 |
| B.1.1.7 | Temperature | 0.123 | 0.163 | 0.751 | 0.453 |
| B.1.1.7 | Rainfall | 1.537 | 0.670 | 2.295 | 0.022 |
| B.1.1.7 | Lockdown 2 | 1.995 | 0.666 | 2.997 | 0.003 |
| B.1.1.7 | Tier 2 | 0.208 | 0.926 | 0.224 | 0.822 |
| B.1.1.7 | Tier 3 | 2.474 | 0.522 | 4.737 | <0.001 |
| B.1.1.7 | IMD 10th Decile | 0.412 | 0.475 | 0.868 | 0.386 |
| B.1.1.7 | Total Population | 0.065 | 0.012 | 5.365 | <0.001 |
| B.1.177 | Intercept | 0.497 | 0.460 | 1.079 | 0.281 |
| B.1.177 | Temperature | -0.189 | 0.029 | -6.433 | <0.001 |
| B.1.177 | Rainfall | -0.027 | 0.052 | -0.518 | 0.605 |

| | | | | | |
|---|---|---|---|---|---|
| B.1.177 | Lockdown 2 | 0.031 | 0.077 | 0.410 | 0.682 |
| B.1.177 | Tier 2 | 0.275 | 0.071 | 3.853 | <0.001 |
| B.1.177 | Tier 3 | 0.060 | 0.093 | 0.640 | 0.522 |
| B.1.177 | Restaurant Subsidy | 0.048 | 0.102 | 0.467 | 0.640 |
| B.1.177 | IMD 10th Decile | 0.356 | 0.212 | 1.677 | 0.094 |
| B.1.177 | Total Population | 0.054 | 0.006 | 9.299 | <0.001 |
| B.1.177.10 | Intercept | -1.887 | 1.561 | -1.209 | 0.227 |
| B.1.177.10 | Temperature | -0.352 | 0.103 | -3.412 | <0.001 |
| B.1.177.10 | Rainfall | -0.049 | 0.244 | -0.200 | 0.842 |
| B.1.177.10 | Lockdown 2 | 0.010 | 0.267 | 0.037 | 0.971 |
| B.1.177.10 | Tier 2 | 0.237 | 0.247 | 0.959 | 0.337 |
| B.1.177.10 | Tier 3 | 0.416 | 0.247 | 1.684 | 0.092 |
| B.1.177.10 | IMD 10th Decile | 1.089 | 0.638 | 1.707 | 0.088 |
| B.1.177.10 | Total Population | 0.071 | 0.017 | 4.067 | <0.001 |

**Table S8**: Summary of the final version of the second-wave lineages GLMM with an AR1 term for week (grouped by postcode); effects of spatial (10th Decile of IMD, postcode district total population, and total cases of lineages B.1.1.1 and B.1.1.119 during the first wave (up to week 28)) and temporal variables (mean weekly temperature and rainfall, and government interventions and subsidy) on the total number of positive PCR tests of the six second wave lineages (B.1.1.309, B.1.1.315, B.1.1.37, B.1.1.7, B.1.177, and B.1.177.10) recorded each week of 2020 in each postcode district. All temporal variables include a two-week time lag to account for the delay in symptom onset (and testing) after infection. Variables with a VIF > 3 were removed from this model (Temperature). Estimates are on the original model scale (log).

| Variable | Estimate | Std. Error | z value | p value |
|---|---|---|---|---|
| Intercept | -1.187 | 0.163 | -7.299 | <0.001 |
| First-wave cases | 0.006 | 0.008 | 0.842 | 0.400 |
| Rainfall | 0.122 | 0.025 | 4.831 | <0.001 |
| Lockdown 2 | 0.188 | 0.037 | 5.051 | <0.001 |
| Tier 2 | 0.310 | 0.048 | 6.511 | <0.001 |
| Tier 3 | 0.386 | 0.049 | 7.953 | <0.001 |
| Restaurant Subsidy | -0.220 | 0.048 | -4.606 | <0.001 |
| IMD 10th Decile | 0.245 | 0.232 | 1.053 | 0.292 |
| Total Population | 0.059 | 0.006 | 10.200 | <0.001 |

**Table S9**: Summary of the full version of the second-wave lineages GLMM with an AR1 term for week (grouped by postcode); effects of spatial (10th Decile of IMD, postcode district total population, and total cases of lineages B.1.1.1 and B.1.1.119 during the first wave (up to week 28)) and temporal variables (mean weekly temperature and rainfall, week of year, and government interventions and subsidy) on the total number of positive PCR tests of the six second wave lineages (B.1.1.309, B.1.1.315, B.1.1.37, B.1.1.7, B.1.177, and B.1.177.10) recorded each week of 2020 in each postcode district. All temporal variables include a two-week time lag to account for the delay in symptom onset (and testing) after infection. The VIF of Temperature was > 3, which means all of the below standard errors, z values, and p values may be unreliable (though the estimates are accurate). Estimates are on the original model scale (log).

| Variable | Estimate | Std. Error | z value | p value |
|---|---|---|---|---|
| Intercept | -0.063 | 0.347 | -0.183 | 0.855 |
| First-wave cases | 0.006 | 0.008 | 0.839 | 0.402 |
| Temperature | -0.075 | 0.021 | -3.596 | <0.001 |
| Rainfall | 0.037 | 0.034 | 1.072 | 0.284 |
| Lockdown 2 | 0.023 | 0.058 | 0.403 | 0.687 |
| Tier 2 | 0.198 | 0.056 | 3.516 | <0.001 |
| Tier 3 | 0.196 | 0.071 | 2.738 | 0.006 |
| Restaurant Subsidy | -0.058 | 0.066 | -0.888 | 0.375 |
| IMD 10th Decile | 0.246 | 0.233 | 1.054 | 0.292 |
| Total Population | 0.059 | 0.006 | 10.147 | <0.001 |

### 2. Model Specifications

#### 2.1 Mixed Effects Models

- Estimation method: maximum likelihood (the default).
- Fixed effects: total population per postcode, proportion of the population in the $10^{th}$ decile of the Index of Multiple Deprivation (IMD) per postcode, mean weekly temperature and rainfall, government restaurant subsidy, first national lockdown, second national lockdown, and tier 2 and tier 3 restrictions in Teesside.
    - Total population was rescaled by dividing by 1000, to measure population in thousands rather than single people.
    - The interventions and subsidy were coded numerically according to the procedure of Hunter et al., 2021 (https://www.eurosurveillance.org/content/10.2807/1560-7917.ES.2021.26.28.2001401): 0 before they were imposed, 1 for the first week of imposition, increasing by 1 for each further week of duration, and reverting to 0 after the end of the intervention. This was to allow the effect to incorporate duration as well as presence/absence.
    - Two-week time lags were applied to the temporal variables: temperature, rainfall, lockdown 1, lockdown 2, tier 2, tier 3, and the restaurant subsidy (the values of these variables were shifted forward in time by 2 weeks, e.g. the values at week 1 were moved to week 3 etc.).

##### AR1 term for week (grouped by postcode):

- Family: Poisson (with log link function).
- Random effects: autoregressive term of order 1, for week of year, grouped by postcode; and a separate random intercept for postcode. The AR1 term was fitted without an intercept, as recommended in glmmTMB package documentation (https://cran.r-project.org/web/packages/glmmTMB/vignettes/covstruct.html#construction-of-structured-covariance-matrices).
  e.g. + (1|Postcode) + ar1(Week + 0|Postcode)
    - The model for Lineage B.1.1.309 demonstrated poor fit in terms of residual autocorrelation, and so it was refit without the random intercept term for postcode (this term had a variance that was almost 0).
- Additional fixed effects: the model examining cases of the second-wave lineages included the total number of cases of the two first-wave lineages (B.1.1.1, B.1.1.119), summed across weeks 11 to 27, per postcode as a fixed effect.

##### Random gradient for week for each postcode:

- Family: negative binomial (with log link function.
- Additional fixed effects: this model included week of year as a fixed effect to attempt to control for the effect of time on cases.
- Random effects: a random gradient for week of year, grouped by postcode; and a random intercept for postcode.
  e.g. + (Week + 1|Postcode)

#### 2.2 Disease Mapping Models

- The adjacency matrix for the Teesside postcode districts was constructed using rook contiguity: shared borders between adjacent areas must be edges (multiple points) rather than single vertices (single points). This is appropriate in the context of disease transmission between UK postcodes districts, as a single point would represent a tiny area of linear contact that is not representative of a true geographical boundary. However, the arrangement and irregular shape of the postcode polygons in the shapefile means that it would make no difference whether or not we used queen or rook contiguity, as all polygons that connect do so by more than one point.
  e.g. poly2nb(shape_file, queen = FALSE)
- Family: negative binomial.
- Link function: log (the default).
- Approximation strategy: Laplace.
  e.g. control.inla = list(strategy = "laplace")
- Integration strategy: CCD (complete composite design). This was the default for our models as the number of hyperparameters was greater than 2 ("size for the nbinomial observations (1/overdispersion)", "Precision for PC_num", “Phi for PC_num").
  e.g. control.inla = list(int.strategy = "auto")
- Priors
  - Negative binomial: we used the default values for this family, and the default variant of it (variant = 0). Hyperid: 63001, name: "size", initial: 2.3, fixed: FALSE, prior: "pc.mgamma", param: 7.
  - BYM2 Hyperparameters: we used the default values. prec = list(prior = "pc.prec", param = c(1, 0.01)), phi = list(prior = "pc", param = c(0.5, 0.5)).
  - The sensitivity analysis for the BYM2 phi and precision hyperparameters used the following values.
    Default: prec = list(prior = "pc.prec", param = c(1, 0.01)), phi = list(prior = "pc", param = c(0.5, 0.5)).
    Larger_phi: prec = list(prior = "pc.prec", param = c(1, 0.01)), phi = list(prior = "pc", param = c(0.5, 2 / 3)).
    Larger_prec: prec = list(prior = "pc.prec", param = c(0.5 / 0.31, 0.01)), phi = list(prior = "pc", param = c(0.5, 0.5)).
    Larger_phiprec: prec = list(prior = "pc.prec", param = c(0.5 / 0.31, 0.01)), phi = list(prior = "pc", param = c(0.5, 2 / 3)).
  - Full details of all hyperparameters of the all-cases model are given as a print-out from the R console below.

```
List of 4
 $ predictor:List of 1
   $ hyper:List of 1
     $ theta:List of 9
       $ hyperid   : num 53001
       $ name      : chr "log precision"
       $ short.name: chr "prec"
       $ initial   : num 13.8
```

```
      $ fixed     : logi TRUE
      $ prior     : chr "loggamma"
      $ param     : num [1:2] 1e+00 1e-05
 $ family   :List of 1
  $ :List of 4
   $ hyperid: chr "INLA.Data1"
   $ label  : chr "nbinomial"
   $ hyper  :List of 1
    $ theta:List of 9
     $ hyperid   : num 63001
     $ name      : chr "size"
     $ short.name: chr "size"
     $ initial   : num 2.3
     $ fixed     : logi FALSE
     $ prior     : chr "pc.mgamma"
     $ param     : num 7
   $ link   :List of 1
    $ hyper: list()
 $ fixed    :List of 1
  $ :List of 3
   $ label     : chr "(Intercept)"
   $ prior.mean: num 0
   $ prior.prec: num 0
 $ random   :List of 1
  $ :List of 3
   $ hyperid    : chr "PC_num"
   $ hyper      :List of 2
    $ theta1:List of 9
     $ hyperid   : num 11001
     $ name      : chr "log precision"
     $ short.name: chr "prec"
     $ prior     : chr "pc.prec"
     $ param     : num [1:2] 1 0.01
     $ initial   : num 4
     $ fixed     : logi FALSE
    $ theta2:List of 9
     $ hyperid   : num 11002
     $ name      : chr "logit phi"
     $ short.name: chr "phi"
     $ prior     : chr "table: -12.6923076923077 -12.6898382145907 -
12.6873687368737 -12.6848992591567 -12.6824297814397 -12.6799603037"|
__truncated__
     $ param     : num(0)
     $ initial   : num -3
     $ fixed     : logi FALSE
   $ group.hyper:List of 1
    $ theta:List of 9
     $ hyperid   : num 40001
     $ name      : chr "logit correlation"
     $ short.name: chr "rho"
     $ initial   : num 1
```

```
$ fixed     : logi FALSE
$ prior     : chr "normal"
$ param     : num [1:2] 0 0.2
```

#### 3. Model Validation

##### 3.1 Disease Mapping Models

All-cases

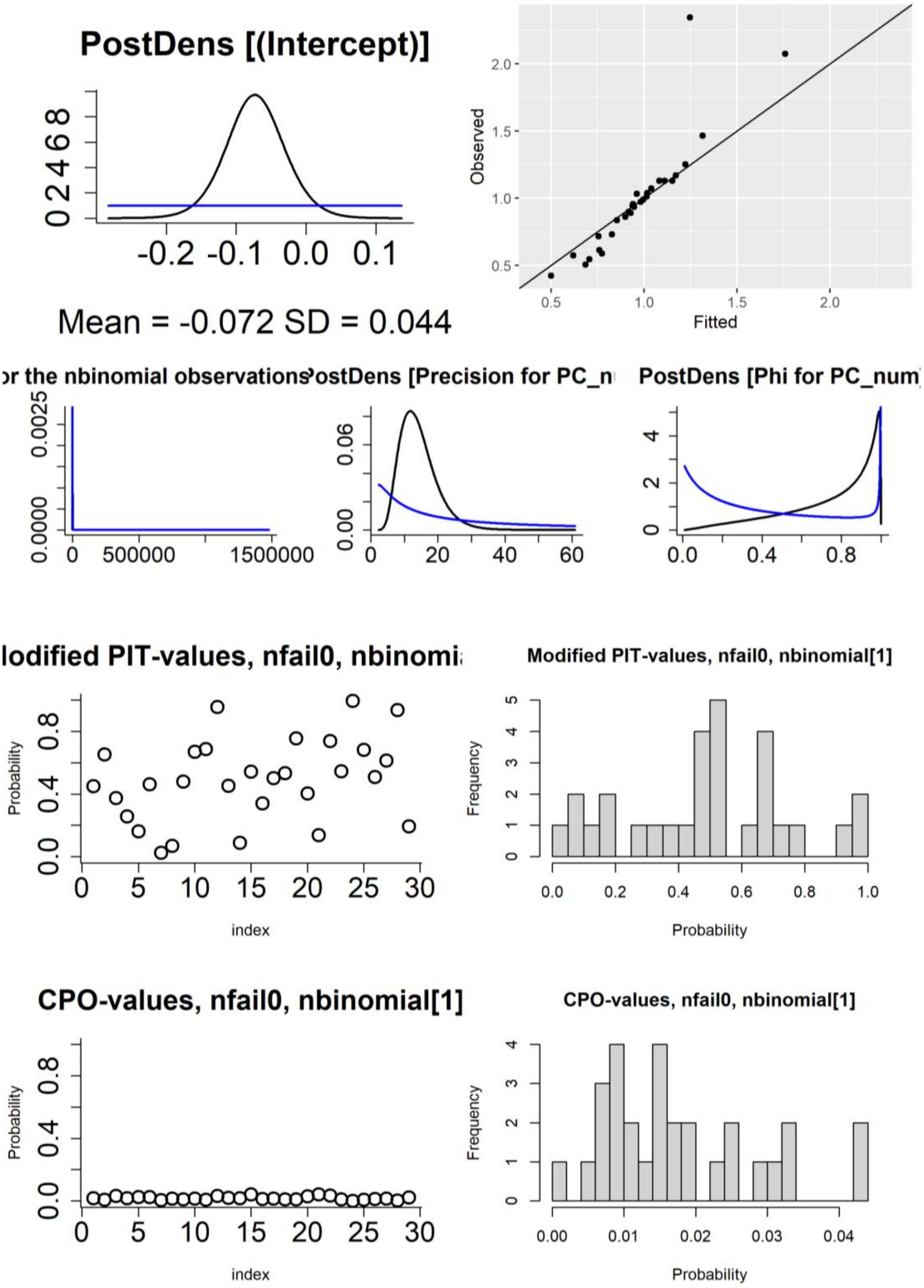

**Figure S5**: Figures used to assess model fit for the all-cases disease mapping model, fit with default priors and other settings. First row: density of the posterior marginals of the intercept (including prior), observed vs fitted values. Second row: posterior density (and priors) of the hyperparameters (size for the negative binomial observations, and precision and phi for the BYM2 random effect). Third row: probability integral transforms (PIT) values (modified/adjusted version appropriate for count data) per postcode area, histogram of PIT values. Fourth row: conditional predictive ordinate (CPO) values per postcode area, histogram of CPO values.

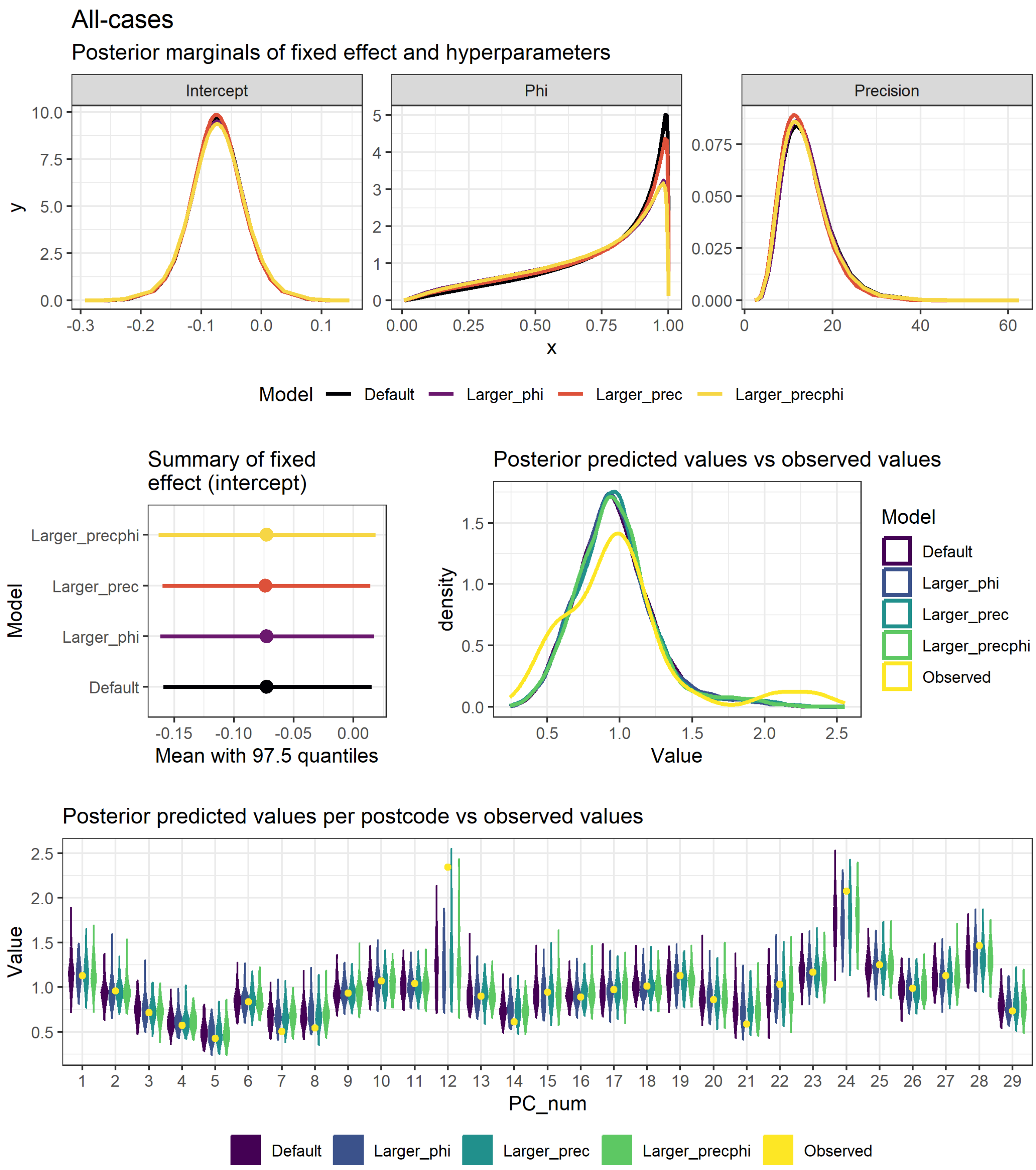

**Figure S6**: Figures used to assess model fit and sensitivity to hyperparameter values for the all-cases disease mapping model, fit with the "laplace" approximation method. These plots compare the values for 4 models, each fit with different hyperparameter values (see section 2.2 above and model code in the analysis repository (https://doi.org/10.25405/data.ncl.23815077). First row: density of the posterior marginals of the intercept and phi and precision hyperparameters. Second row: Mean and 97.5% quantiles for the intercept, comparison of the observed values to a sample generated from the posterior distribution.  Third row: comparison of the observed values per postcode area to a sample generated from the posterior distribution (note that the numbers on the x axis do not correspond to the actual postcode numbers).

#### Lineage B.1.1.1

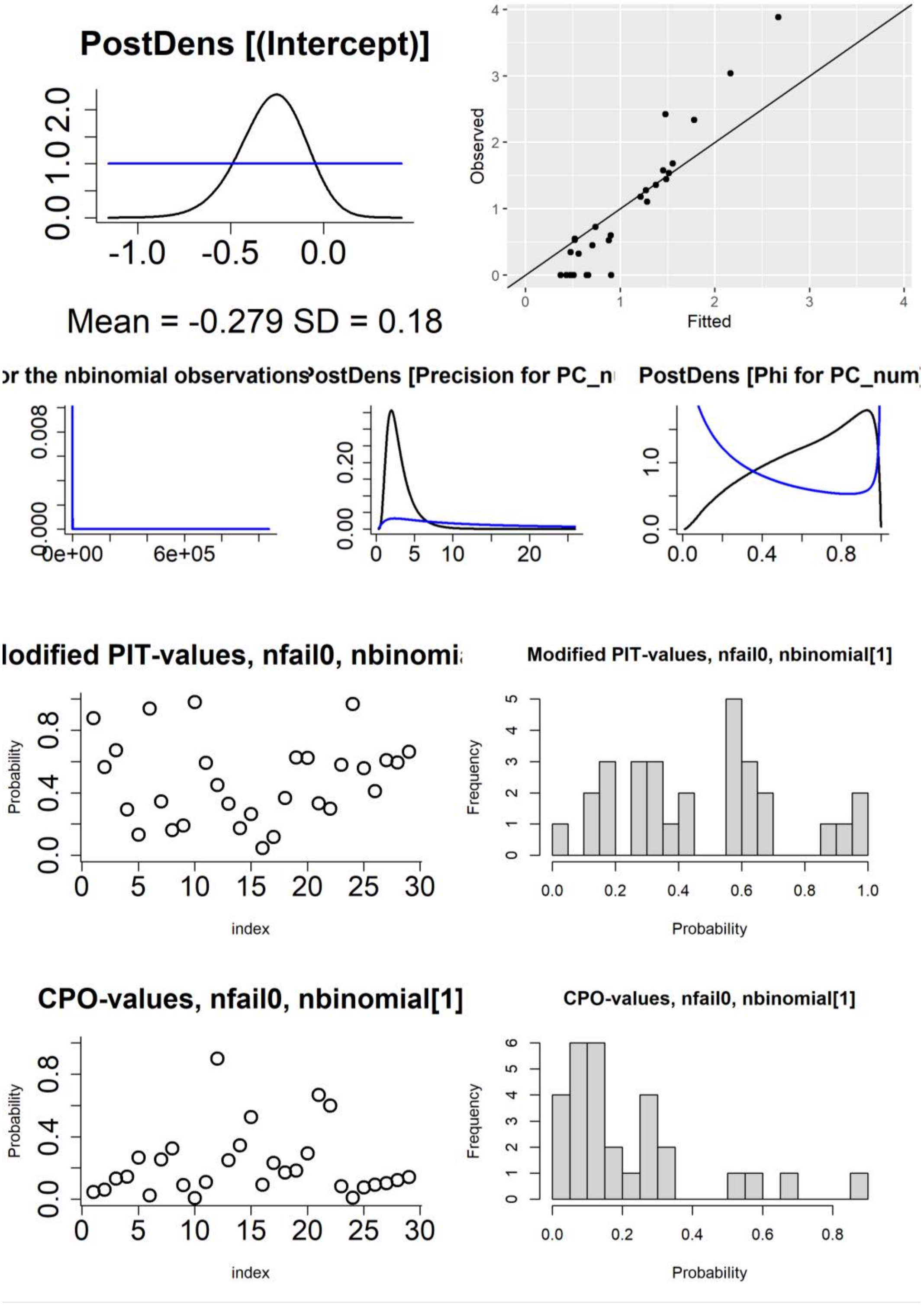

**Figure S7**: Figures used to assess model fit for the Lin6_B.1.1.1 disease mapping model, fit with default priors and other settings. First row: density of the posterior marginals of the intercept (including prior), observed vs fitted values. Second row: posterior density (and priors) of the hyperparameters (size for the negative binomial observations, and precision and phi for the BYM2 random effect). Third row: probability integral transforms (PIT) values (modified/adjusted version appropriate for count data) per postcode area, histogram of PIT values. Fourth row: conditional predictive ordinate (CPO) values per postcode area, histogram of CPO values.

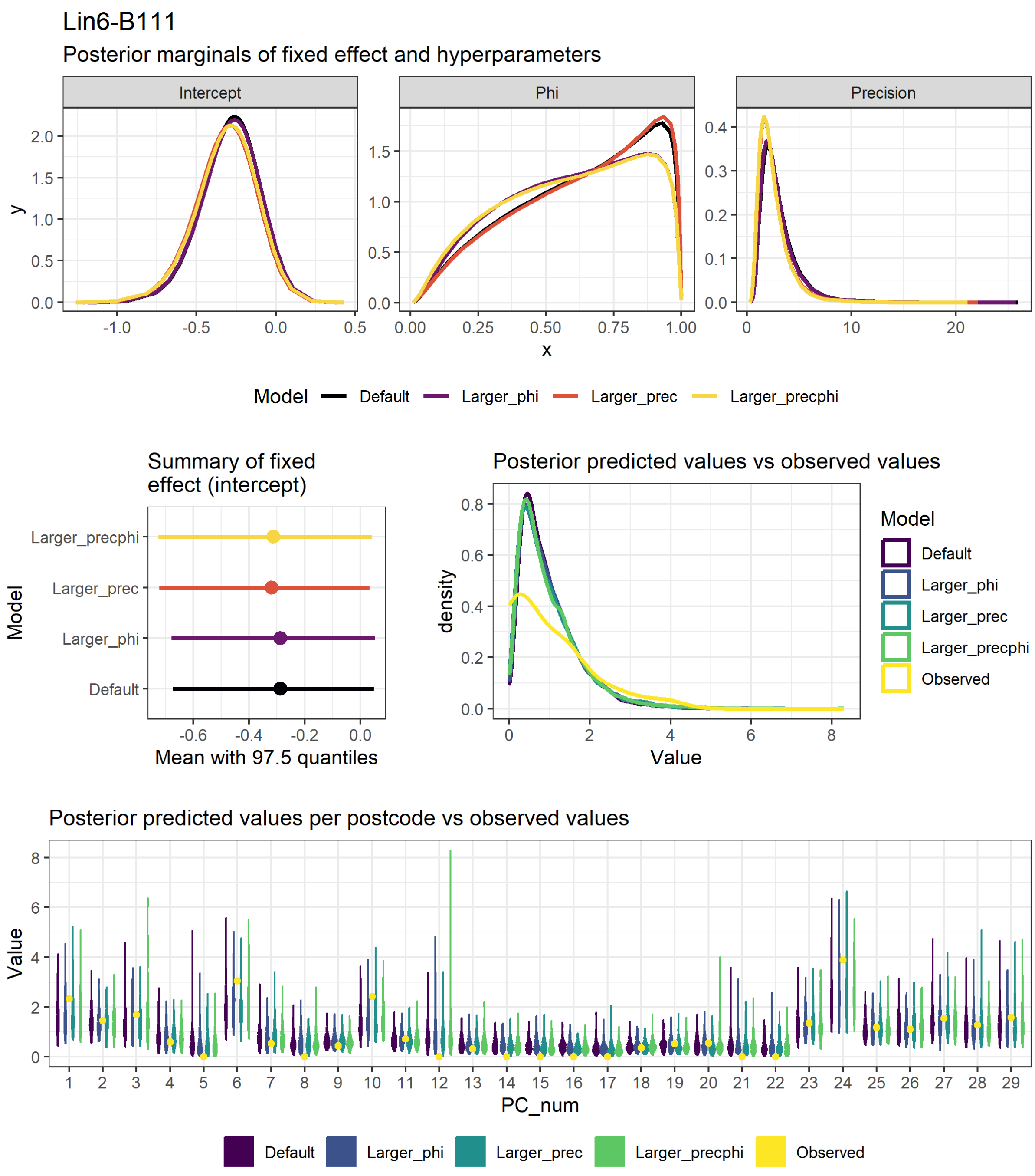

**Figure S8**: Figures used to assess model fit and sensitivity to hyperparameter values for the Lin6_B.1.1.1 disease mapping model, fit with the "laplace" approximation method. These plots compare the values for 4 models, each fit with different hyperparameter values (see section 2.2 above and model code in the analysis repository (https://doi.org/10.25405/data.ncl.23815077). First row: density of the posterior marginals of the intercept and phi and precision hyperparameters. Second row: Mean and 97.5% quantiles for the intercept, comparison of the observed values to a sample generated from the posterior distribution. Third row: comparison of the observed values per postcode area to a sample generated from the posterior distribution (note that the numbers on the x axis do not correspond to the actual postcode numbers).

#### Lineage B.1.1.119

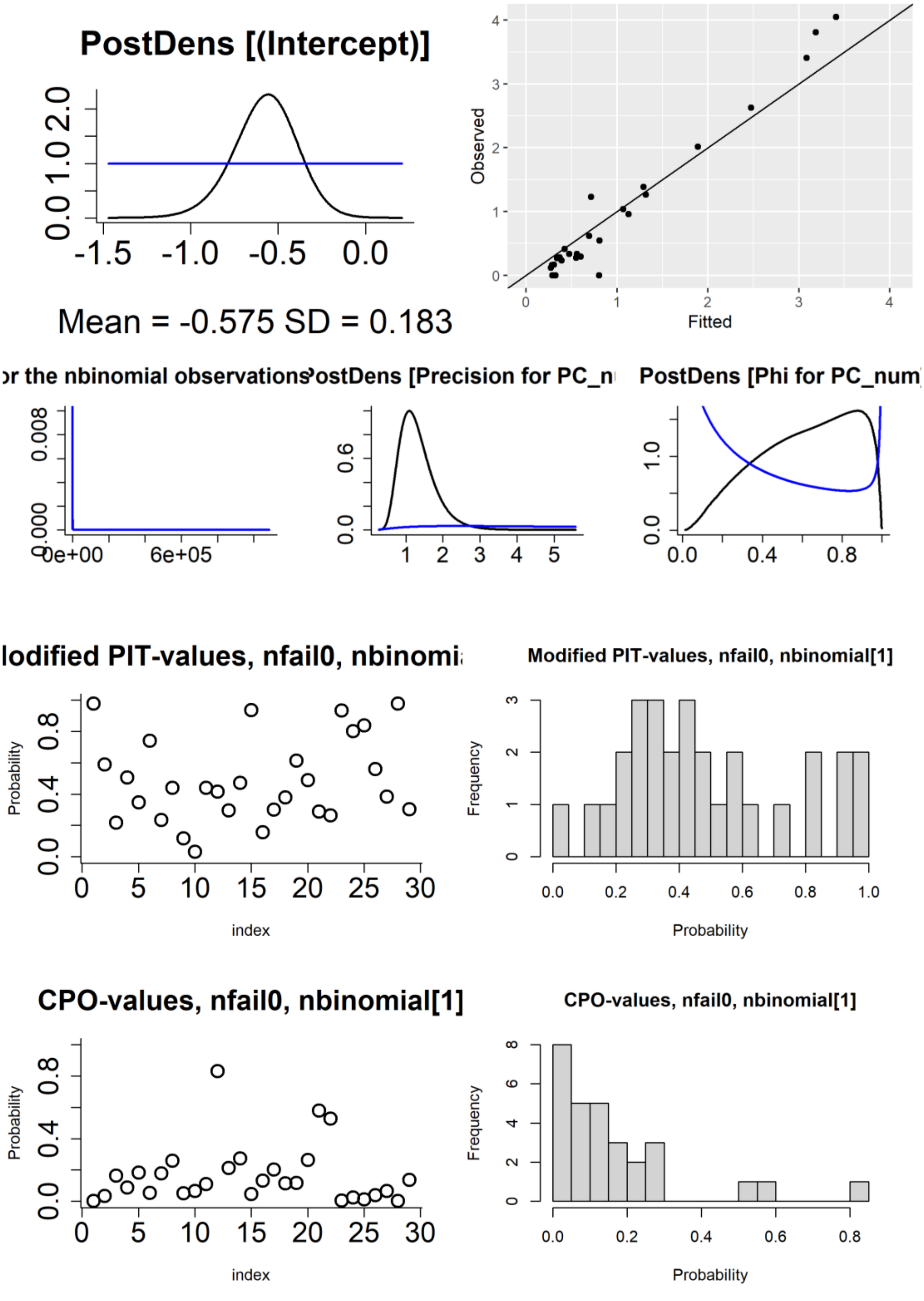

**Figure S9**: Figures used to assess model fit for the Lin10_B.1.1.119 disease mapping model, fit with default priors and other settings. First row: density of the posterior marginals of the intercept (including prior), observed vs fitted values. Second row: posterior density (and priors) of the hyperparameters (size for the negative binomial observations, and precision and phi for the BYM2 random effect). Third row: probability integral transforms (PIT) values (modified/adjusted version appropriate for count data) per postcode area, histogram of PIT values. Fourth row: conditional predictive ordinate (CPO) values per postcode area, histogram of CPO values.

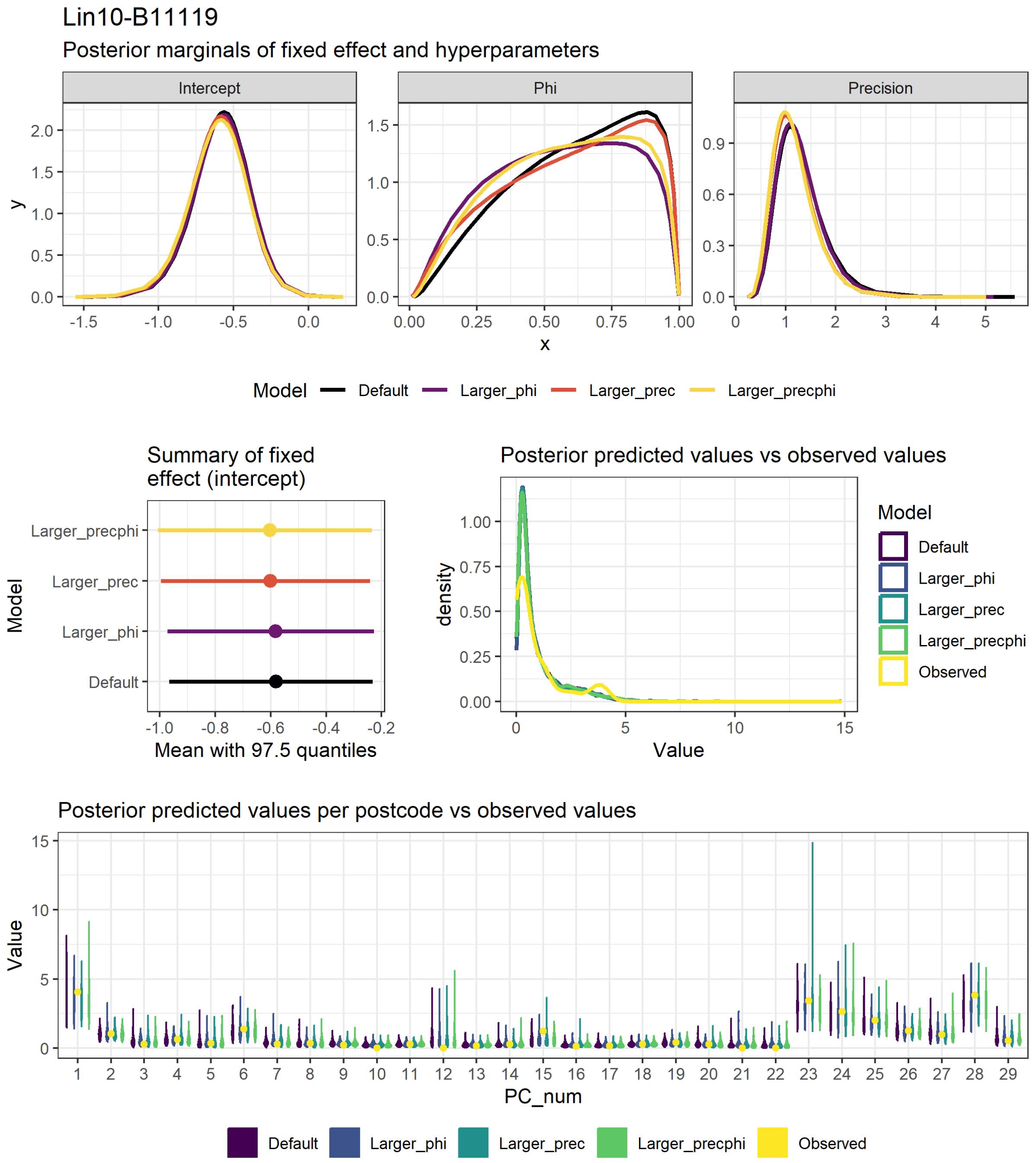

**Figure S10**: Figures used to assess model fit and sensitivity to hyperparameter values for the Lin10_B.1.1.119 disease mapping model, fit with the "laplace" approximation method. These plots compare the values for 4 models, each fit with different hyperparameter values (see section 2.2 above and model code in the analysis repository (https://doi.org/10.25405/data.ncl.23815077). First row: density of the posterior marginals of the intercept and phi and precision hyperparameters. Second row: Mean and 97.5% quantiles for the intercept, comparison of the observed values to a sample generated from the posterior distribution. Third row: comparison of the observed values per postcode area to a sample generated from the posterior distribution (note that the numbers on the x axis do not correspond to the actual postcode numbers).

#### Lineage B.1.1.309

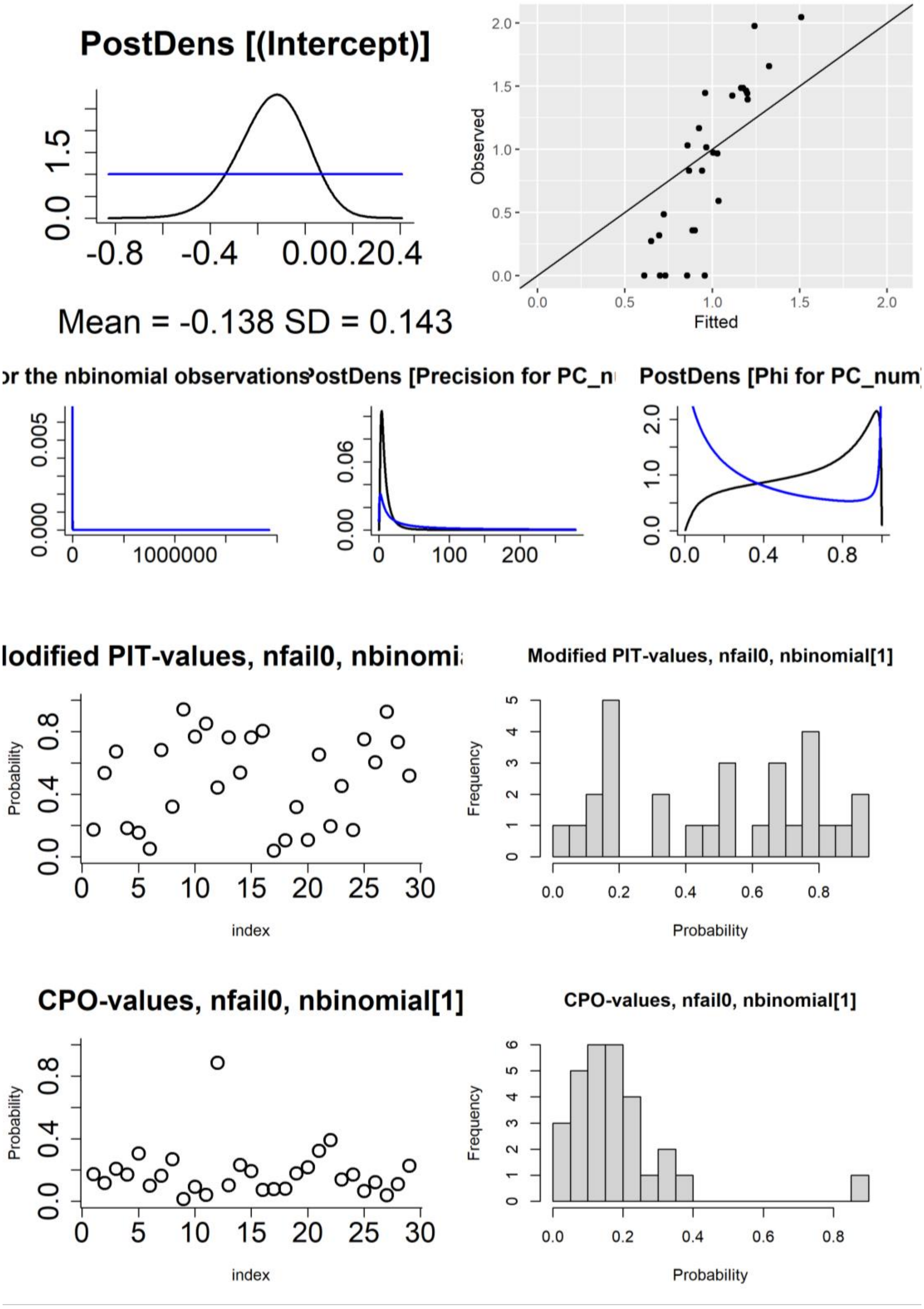

**Figure S11**: Figures used to assess model fit for the Lin35_B.1.1.309 disease mapping model, fit with default priors and other settings. First row: density of the posterior marginals of the intercept (including prior), observed vs fitted values. Second row: posterior density (and priors) of the hyperparameters (size for the negative binomial observations, and precision and phi for the BYM2 random effect). Third row: probability integral transforms (PIT) values (modified/adjusted version appropriate for count data) per postcode area, histogram of PIT values. Fourth row: conditional predictive ordinate (CPO) values per postcode area, histogram of CPO values.

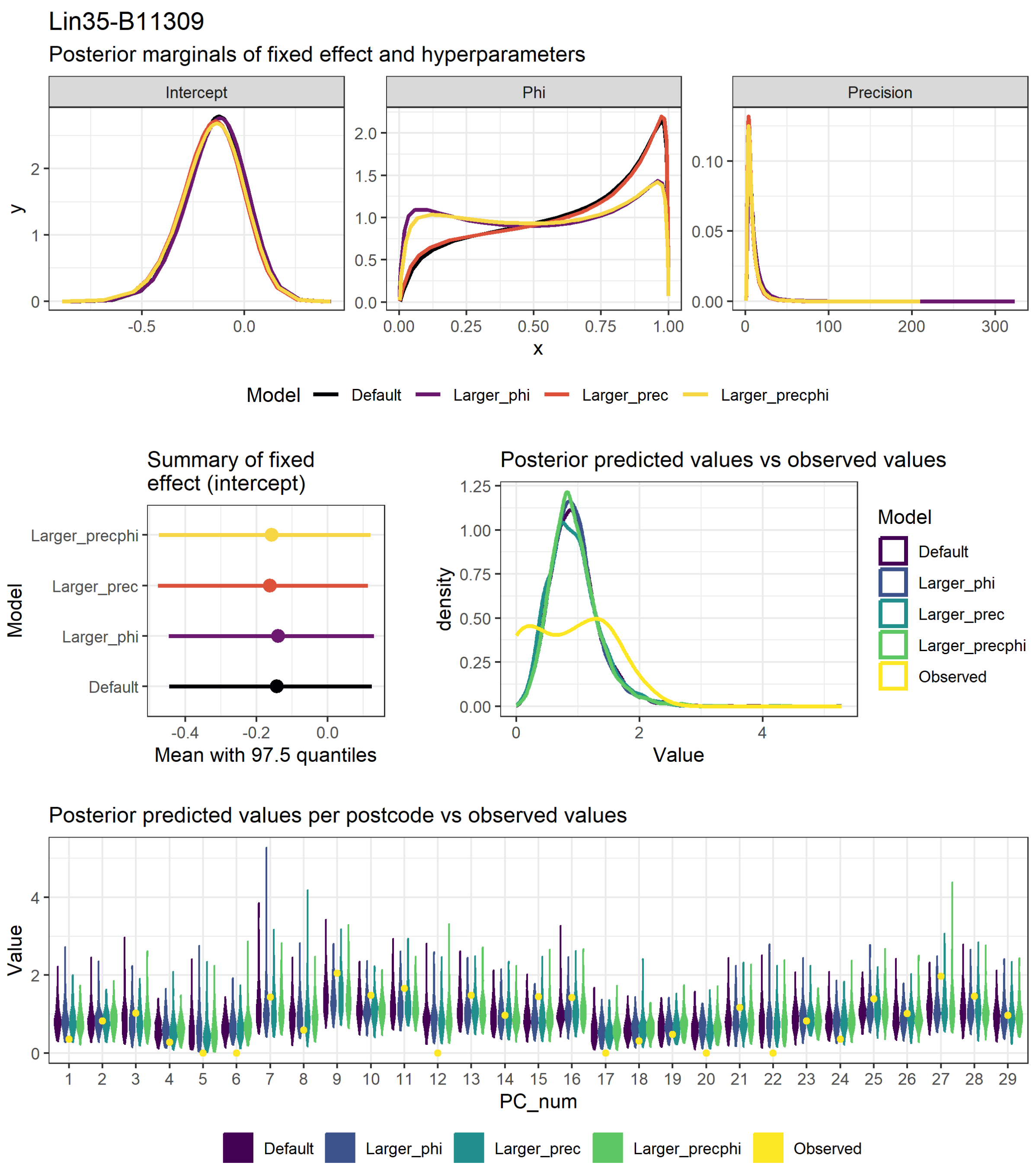

**Figure S12**: Figures used to assess model fit and sensitivity to hyperparameter values for the Lin35_B.1.1.309 disease mapping model, fit with the "laplace" approximation method. These plots compare the values for 4 models, each fit with different hyperparameter values (see section 2.2 above and model code in the analysis repository (https://doi.org/10.25405/data.ncl.23815077). First row: density of the posterior marginals of the intercept and phi and precision hyperparameters. Second row: Mean and 97.5% quantiles for the intercept, comparison of the observed values to a sample generated from the posterior distribution. Third row: comparison of the observed values per postcode area to a sample generated from the posterior distribution (note that the numbers on the x axis do not correspond to the actual postcode numbers).

#### Lineage B.1.1.315

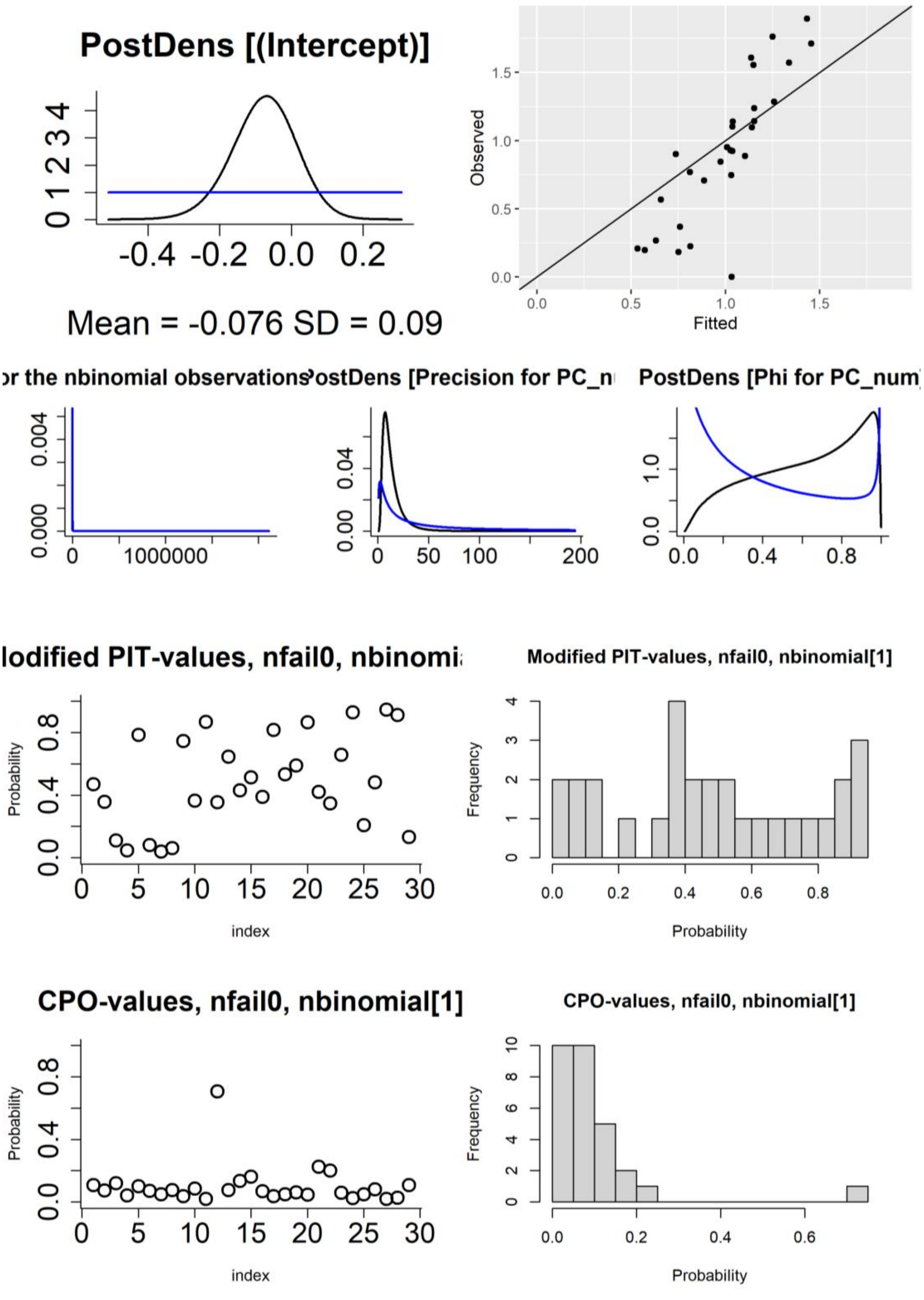

**Figure S13**: Figures used to assess model fit for the Lin37_B.1.1.315 disease mapping model, fit with default priors and other settings. First row: density of the posterior marginals of the intercept (including prior), observed vs fitted values. Second row: posterior density (and priors) of the hyperparameters (size for the negative binomial observations, and precision and phi for the BYM2 random effect). Third row: probability integral transforms (PIT) values (modified/adjusted version appropriate for count data) per postcode area, histogram of PIT values. Fourth row: conditional predictive ordinate (CPO) values per postcode area, histogram of CPO values.

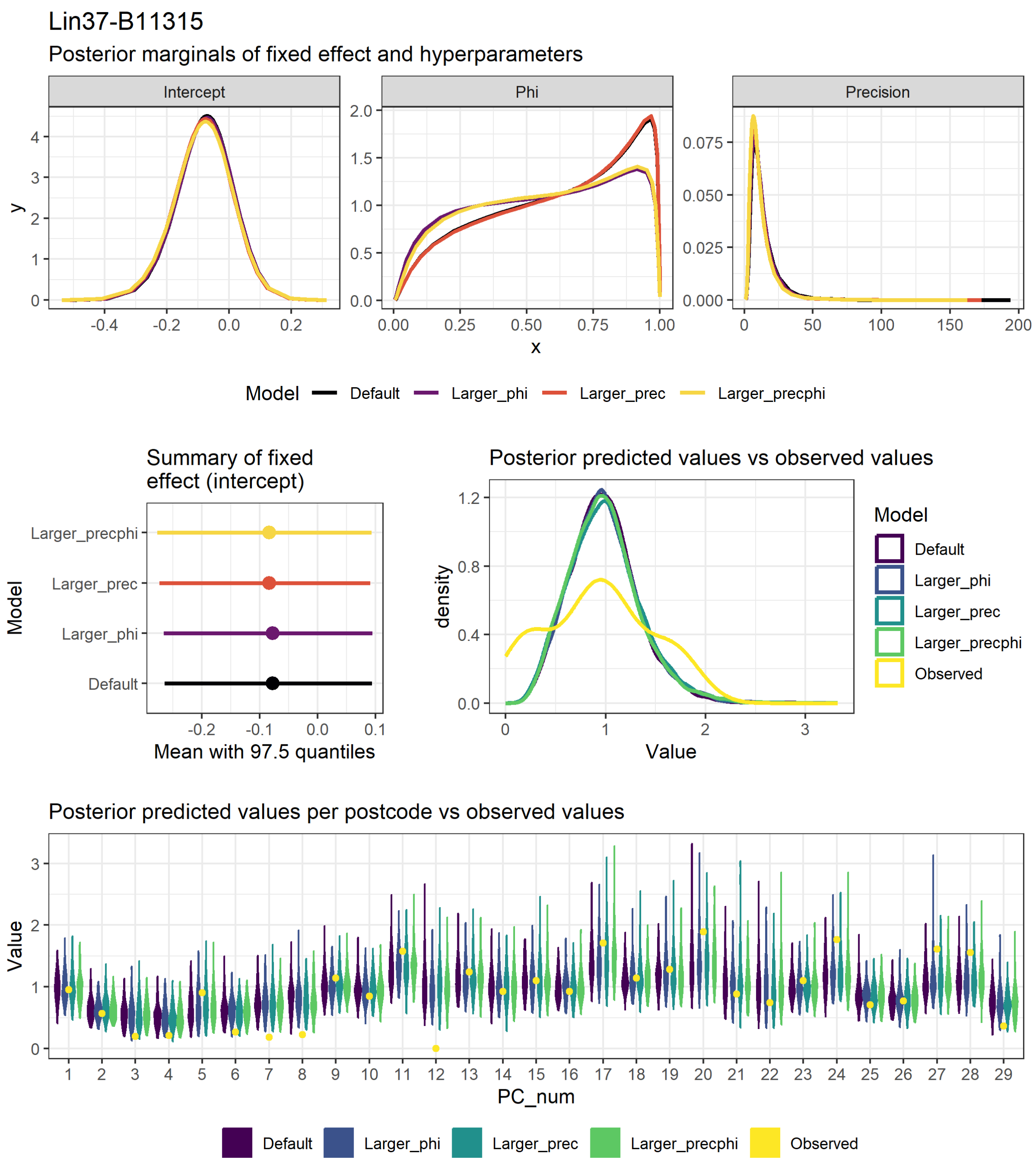

**Figure S14**: Figures used to assess model fit and sensitivity to hyperparameter values for the Lin37_B.1.1.315 disease mapping model, fit with the "laplace" approximation method. These plots compare the values for 4 models, each fit with different hyperparameter values (see section 2.2 above and model code in the analysis repository (https://doi.org/10.25405/data.ncl.23815077). First row: density of the posterior marginals of the intercept and phi and precision hyperparameters. Second row: Mean and 97.5% quantiles for the intercept, comparison of the observed values to a sample generated from the posterior distribution. Third row: comparison of the observed values per postcode area to a sample generated from the posterior distribution (note that the numbers on the x axis do not correspond to the actual postcode numbers).

#### Lineage B.1.1.37

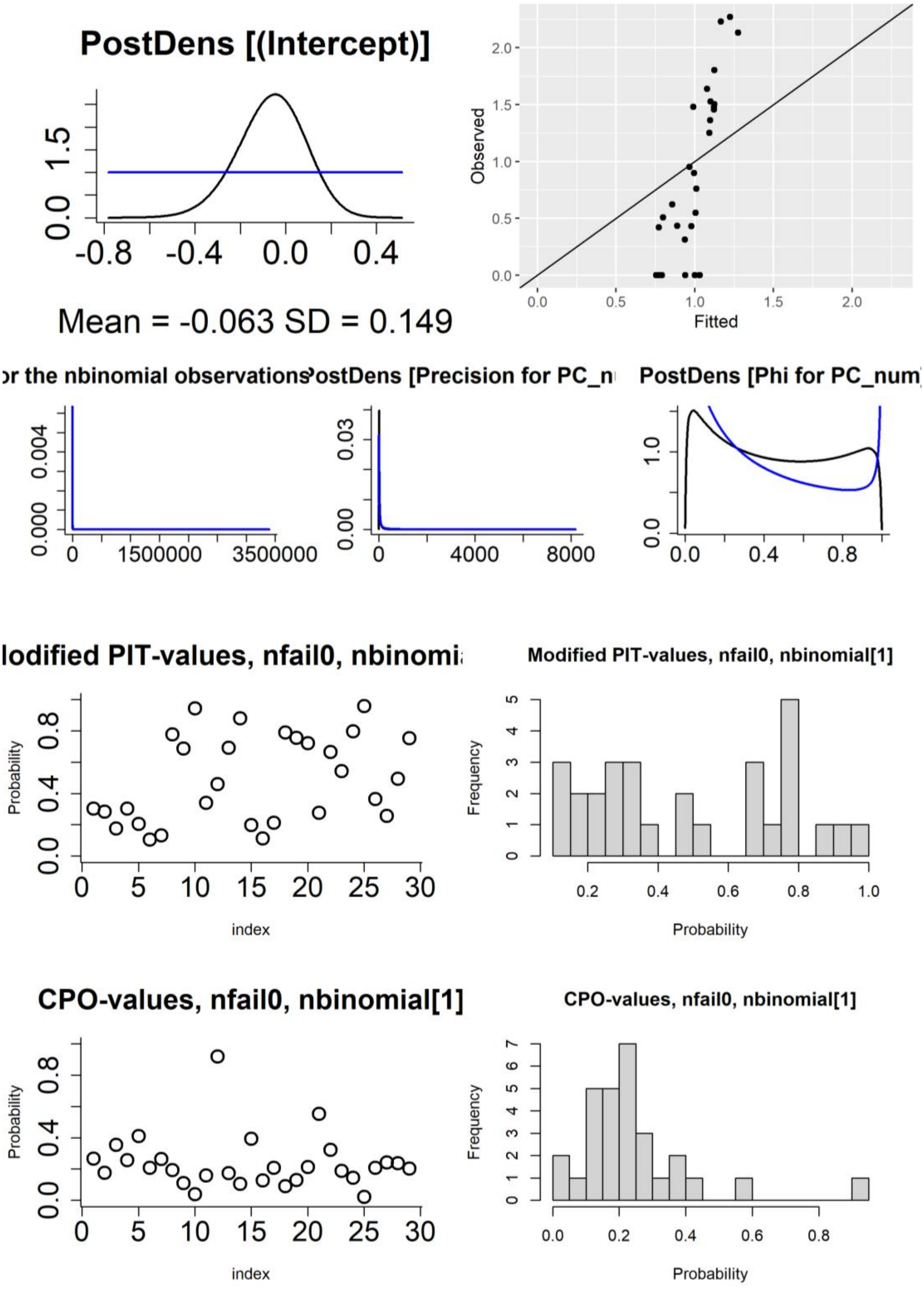

**Figure S15**: Figures used to assess model fit for the Lin39_B.1.1.37 disease mapping model, fit with default priors and other settings. First row: density of the posterior marginals of the intercept (including prior), observed vs fitted values. Second row: posterior density (and priors) of the hyperparameters (size for the negative binomial observations, and precision and phi for the BYM2 random effect). Third row: probability integral transforms (PIT) values (modified/adjusted version appropriate for count data) per postcode area, histogram of PIT values. Fourth row: conditional predictive ordinate (CPO) values per postcode area, histogram of CPO values.

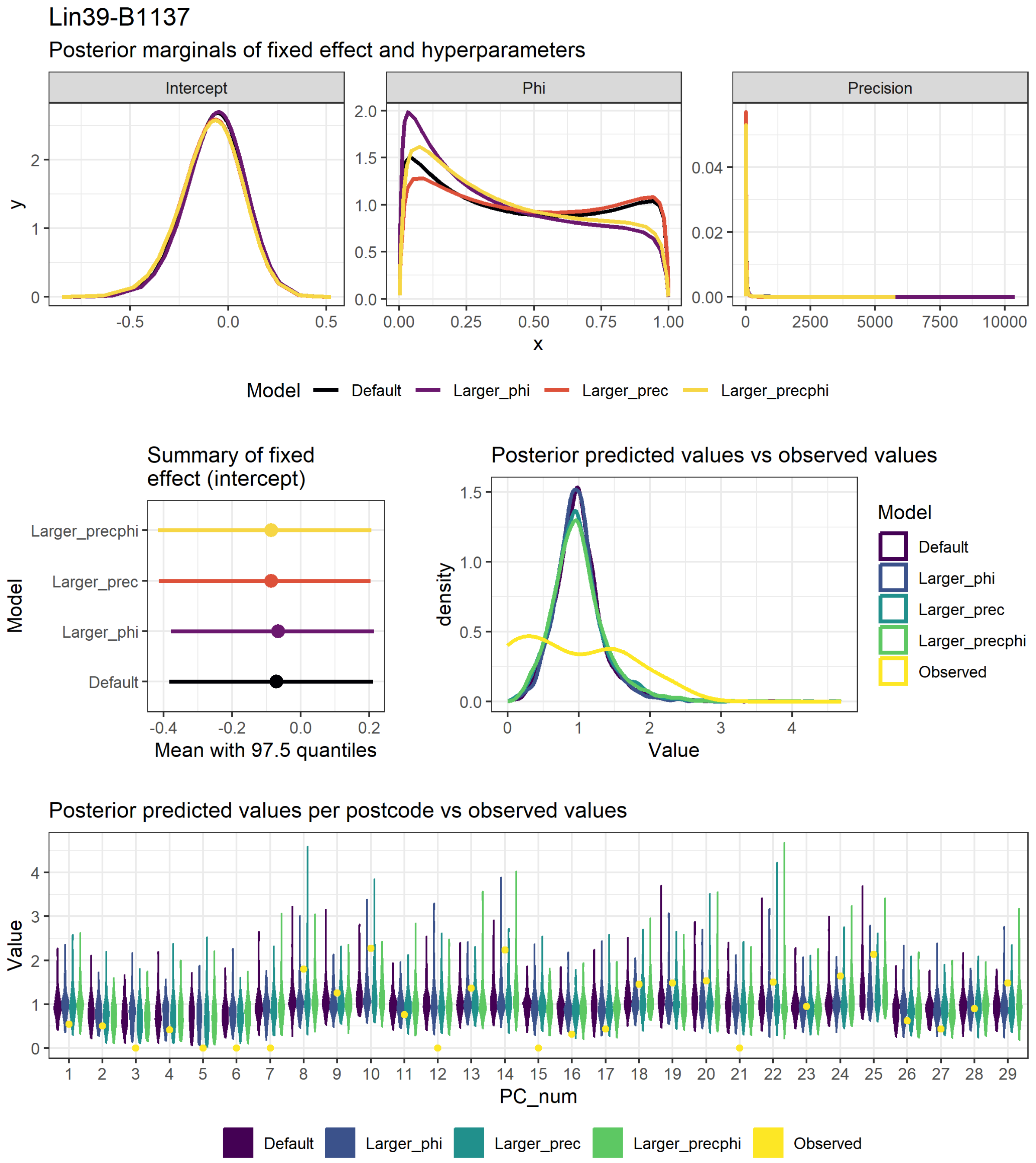

**Figure S16**: Figures used to assess model fit and sensitivity to hyperparameter values for the Lin39_B.1.1.37 disease mapping model, fit with the “laplace” approximation method. These plots compare the values for 4 models, each fit with different hyperparameter values (see section 2.2 above and model code in the analysis repository (https://doi.org/10.25405/data.ncl.23815077). First row: density of the posterior marginals of the intercept and phi and precision hyperparameters. Second row: Mean and 97.5% quantiles for the intercept, comparison of the observed values to a sample generated from the posterior distribution.  Third row: comparison of the observed values per postcode area to a sample generated from the posterior distribution (note that the numbers on the x axis do not correspond to the actual postcode numbers).

#### Lineage B.1.1.7

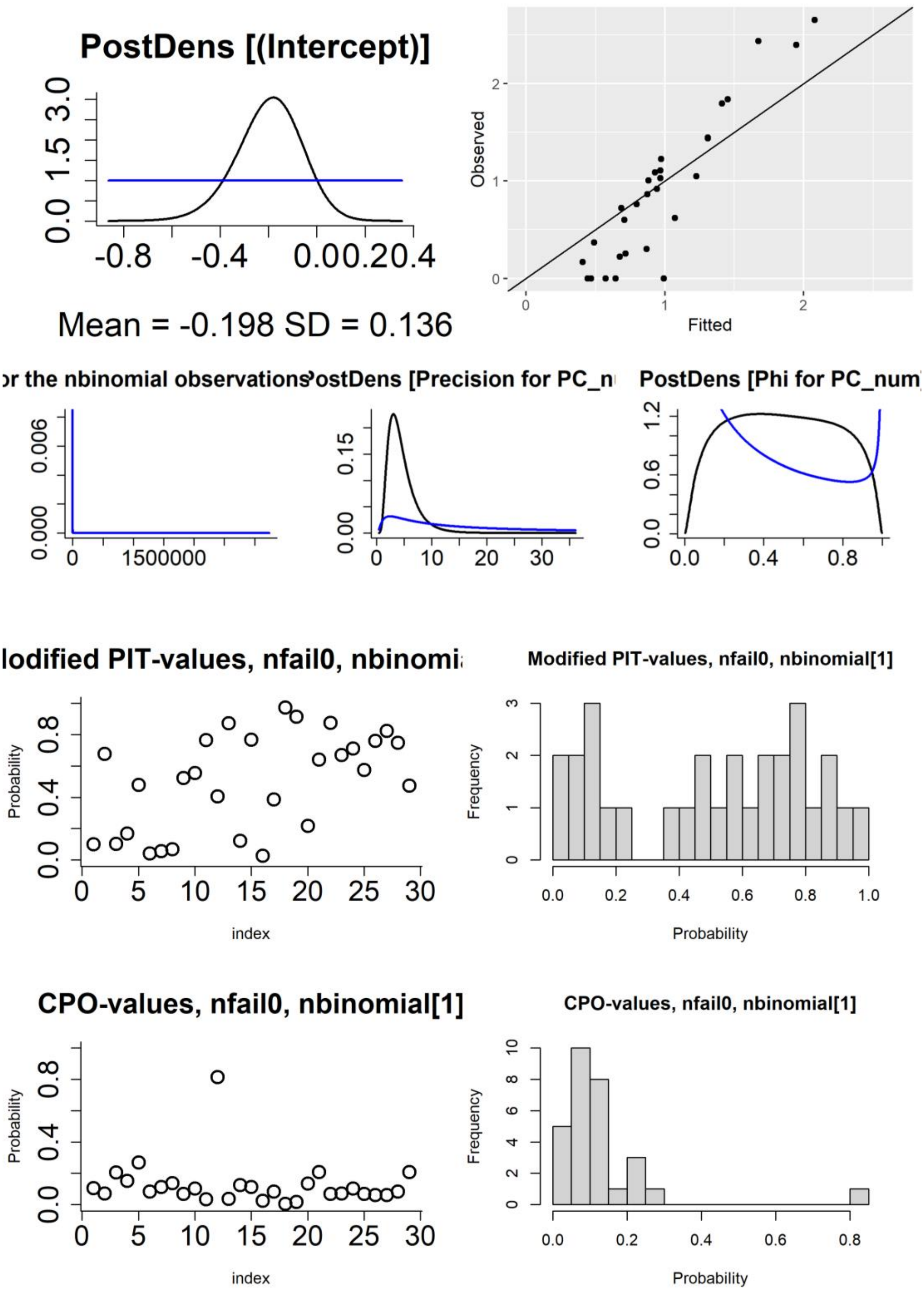

**Figure S17**: Figures used to assess model fit for the Lin45_B.1.1.7 disease mapping model, fit with default priors and other settings. First row: density of the posterior marginals of the intercept (including prior), observed vs fitted values. Second row: posterior density (and priors) of the hyperparameters (size for the negative binomial observations, and precision and phi for the BYM2 random effect). Third row: probability integral transforms (PIT) values (modified/adjusted version appropriate for count data) per postcode area, histogram of PIT values. Fourth row: conditional predictive ordinate (CPO) values per postcode area, histogram of CPO values.

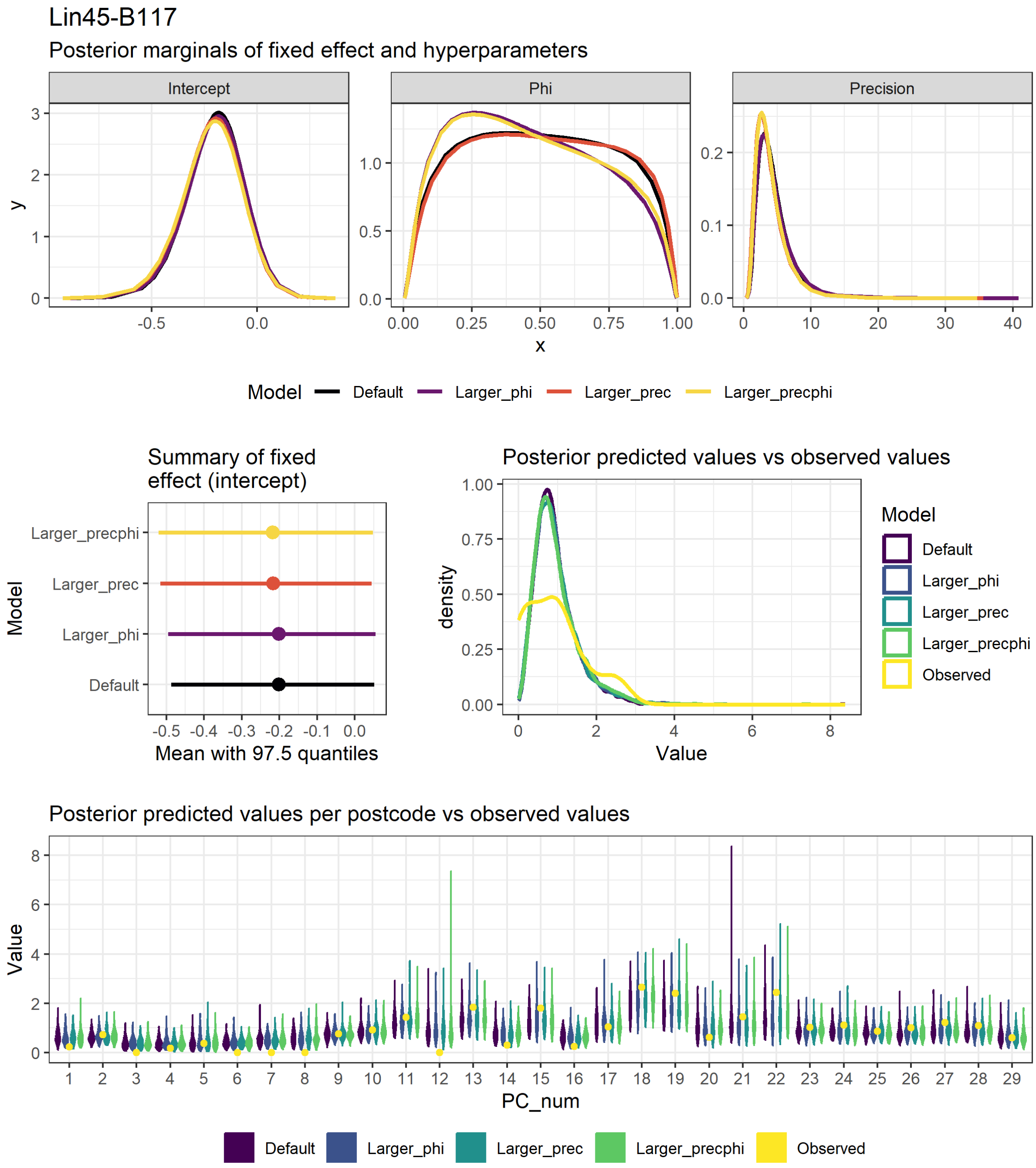

**Figure S18**: Figures used to assess model fit and sensitivity to hyperparameter values for the Lin45_B.1.1.7 disease mapping model, fit with the "laplace" approximation method. These plots compare the values for 4 models, each fit with different hyperparameter values (see section 2.2 above and model code in the analysis repository (https://doi.org/10.25405/data.ncl.23815077). First row: density of the posterior marginals of the intercept and phi and precision hyperparameters. Second row: Mean and 97.5% quantiles for the intercept, comparison of the observed values to a sample generated from the posterior distribution. Third row: comparison of the observed values per postcode area to a sample generated from the posterior distribution (note that the numbers on the x axis do not correspond to the actual postcode numbers).

#### Lineage B.1.177

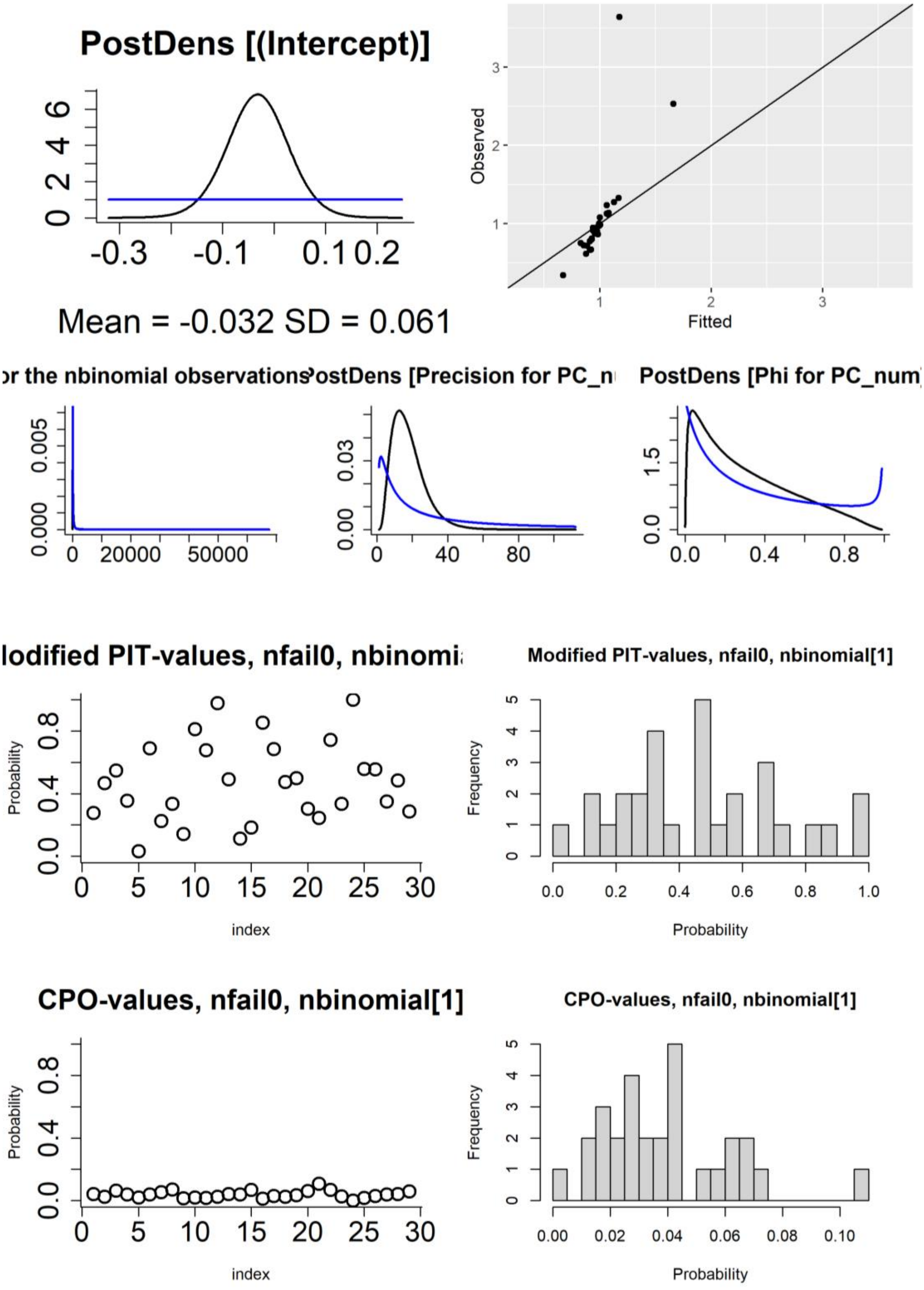

**Figure S19**: Figures used to assess model fit for the Lin51_B.1.177 disease mapping model, fit with default priors and other settings. First row: density of the posterior marginals of the intercept (including prior), observed vs fitted values. Second row: posterior density (and priors) of the hyperparameters (size for the negative binomial observations, and precision and phi for the BYM2 random effect). Third row: probability integral transforms (PIT) values (modified/adjusted version appropriate for count data) per postcode area, histogram of PIT values. Fourth row: conditional predictive ordinate (CPO) values per postcode area, histogram of CPO values.

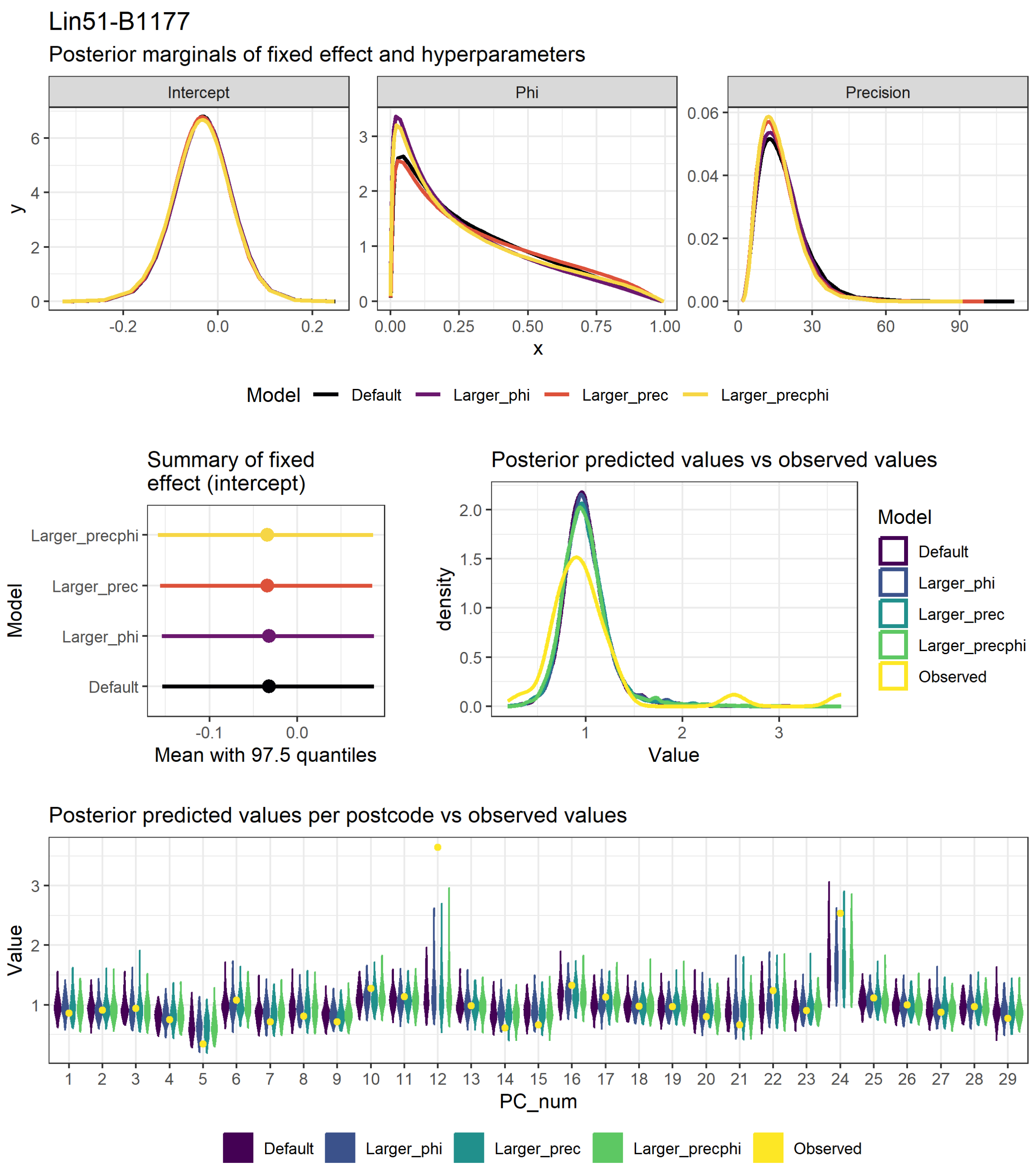

**Figure S20**: Figures used to assess model fit and sensitivity to hyperparameter values for the Lin51_B.1.177 disease mapping model, fit with the "laplace" approximation method. These plots compare the values for 4 models, each fit with different hyperparameter values (see section 2.2 above and model code in the analysis repository (https://doi.org/10.25405/data.ncl.23815077). First row: density of the posterior marginals of the intercept and phi and precision hyperparameters. Second row: Mean and 97.5% quantiles for the intercept, comparison of the observed values to a sample generated from the posterior distribution.  Third row: comparison of the observed values per postcode area to a sample generated from the posterior distribution (note that the numbers on the x axis do not correspond to the actual postcode numbers).

#### Lineage B.1.177.10

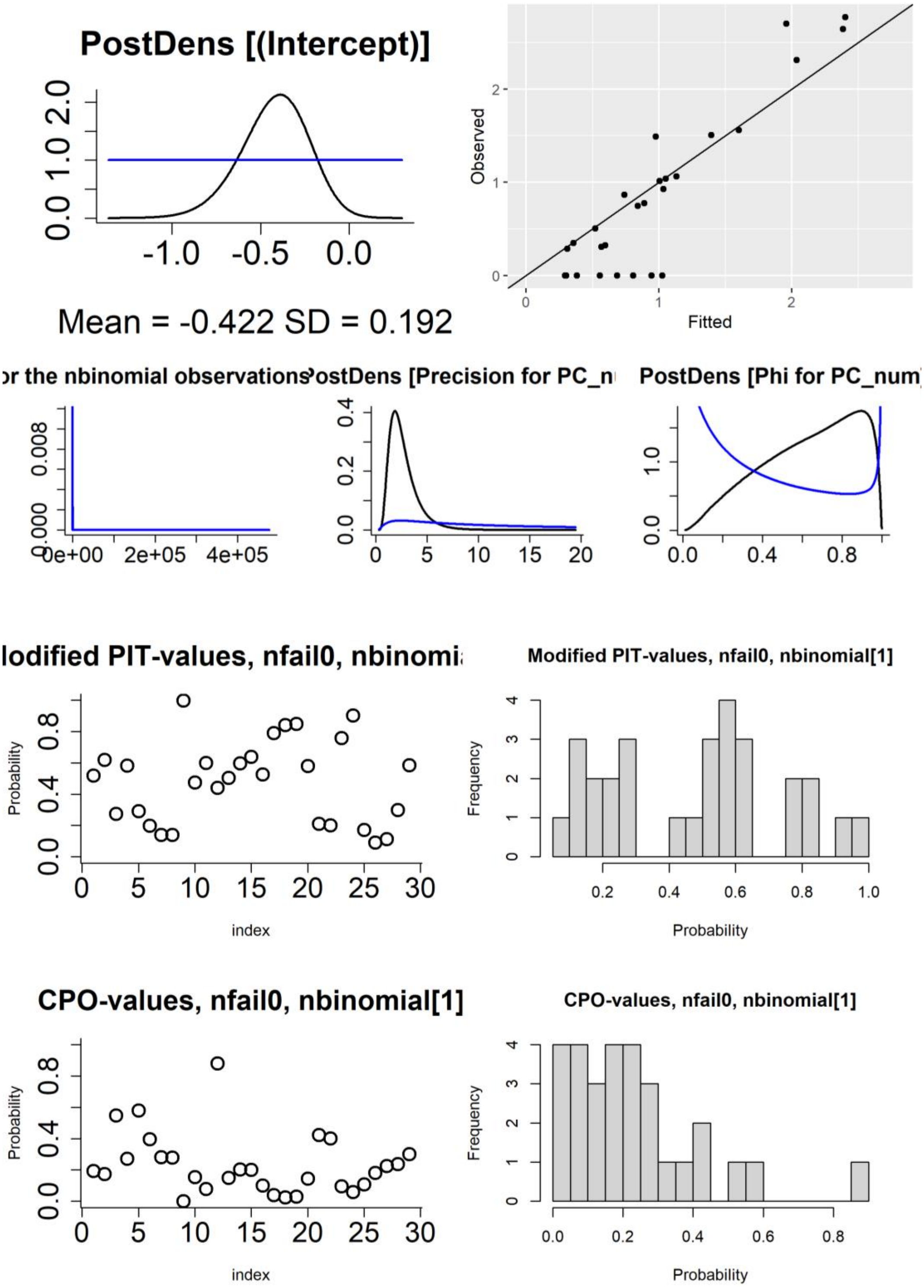

**Figure S21**: Figures used to assess model fit for the Lin52_B.1.177.10 disease mapping model, fit with default priors and other settings. First row: density of the posterior marginals of the intercept (including prior), observed vs fitted values. Second row: posterior density (and priors) of the hyperparameters (size for the negative binomial observations, and precision and phi for the BYM2 random effect). Third row: probability integral transforms (PIT) values (modified/adjusted version appropriate for count data) per postcode area, histogram of PIT values. Fourth row: conditional predictive ordinate (CPO) values per postcode area, histogram of CPO values.

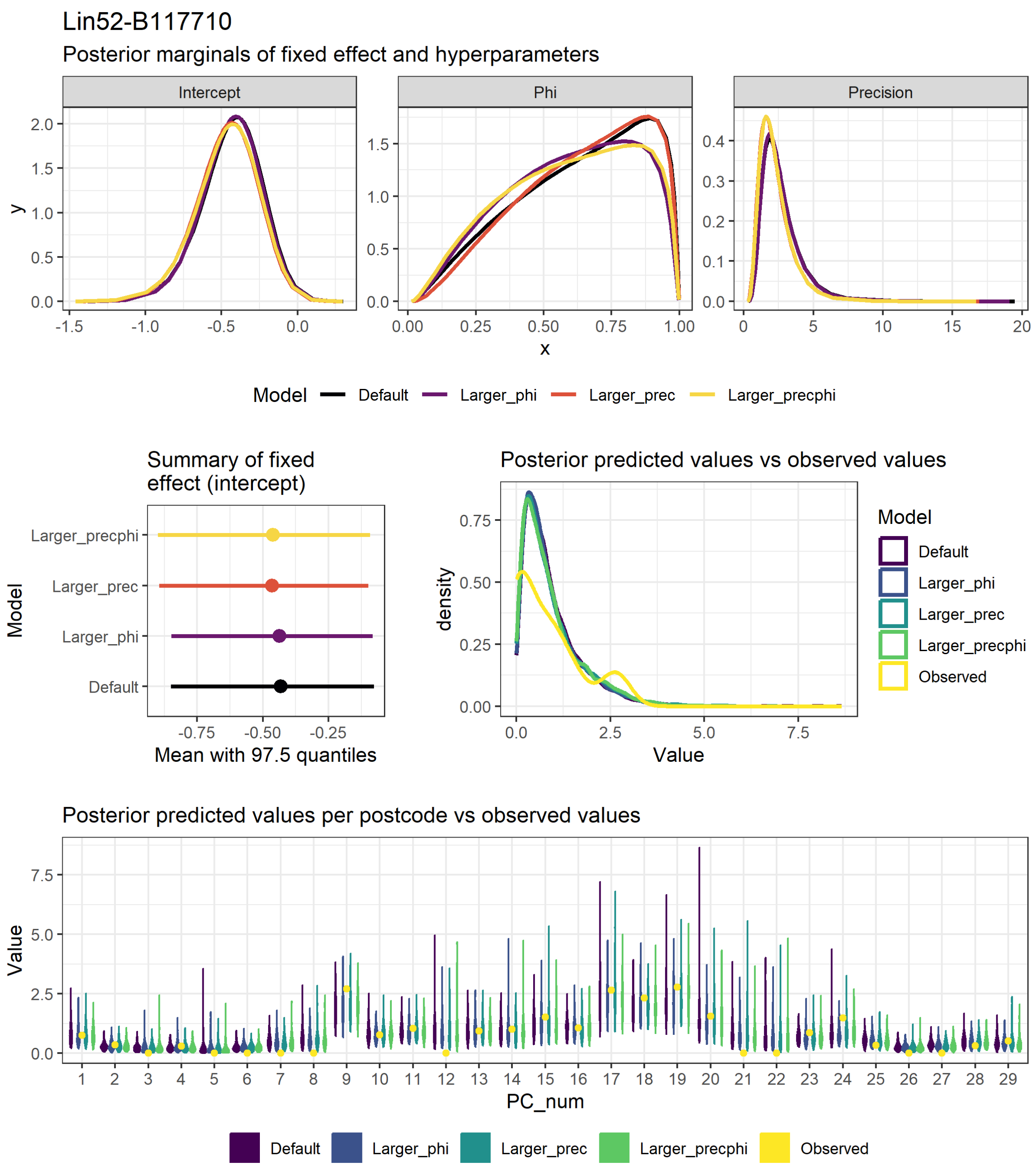

**Figure S22**: Figures used to assess model fit and sensitivity to hyperparameter values for the Lin52_B.1.177.10 disease mapping model, fit with the "laplace" approximation method. These plots compare the values for 4 models, each fit with different hyperparameter values (see section 2.2 above and model code in the analysis repository (https://doi.org/10.25405/data.ncl.23815077). First row: density of the posterior marginals of the intercept and phi and precision hyperparameters. Second row: Mean and 97.5% quantiles for the intercept, comparison of the observed values to a sample generated from the posterior distribution. Third row: comparison of the observed values per postcode area to a sample generated from the posterior distribution (note that the numbers on the x axis do not correspond to the actual postcode numbers).

#### 3.2 Mixed Effects Models

The GLMM models were validated using tests in the DHARMa R package and by comparing the observed values to those fitted by the models. The figures below contain plots created from the DHARMa test functions run on a simulated residuals object (https://cran.r-project.org/web/packages/DHARMa/vignettes/DHARMa.html): QQ plot, including Kolmogorov-Smirnov p-value; residuals against predicted values, including assessment for deviation from the expected mean or quantiles; histogram of the residuals, including assessment for outliers; dispersion test comparing the observed data (red line) with a histogram of residuals; residuals vs time; and the ACF (autocorrelation function), including a Durbin-Watson p-value. The validation figures also contain a VIF (variance inflation factor) plot for each variable in the model, created using the "performance" R package (https://easystats.github.io/performance/reference/check_collinearity.html).

All data sets were modelled using a range of different specifications: a basic model with only a random intercept for postcode; a restricted cubic spline for time (using both 3 and 4 knots) (implemented via the "splines" R package); a smooth spline for time (implemented via the "mgcv" R package); an AR1 term for time (without any grouping); and an AR1 term for time grouped by Postcode. Each of these was compared using the validation plots. For each dataset, the AR1 model that was grouped by postcode proved to be the best fit. The most appropriate distribution family was also assessed for the AR1 models in the same way, with Poisson being the best fit in all cases.

Because collinearity was present in most of our datasets, we used variance inflation factors (VIF) to identify variables to be dropped from the full models (where VIF > 3). In addition to VIF, we also examined variable clustering and redundancy via the "varclus" and "redun" functions of the Hmisc R package (https://www.rdocumentation.org/packages/Hmisc/versions/5.1-2). The output of the redundancy analyses for each of the datasets is in table S10. The redundant variables identified in these analyses matched those with the highest VIF values. Hierarchical cluster analysis plots for each dataset are included below, using both squared Pearson (S23) and Spearman (S24) correlations.

The validation figures for the final (best) models for each dataset are included below (S25, S28: S37). In addition, the all-cases models that include splines for time (a restricted cubic spline (using 4 knots) and a smooth spline) have also been included as an example of the poorer fit that these approaches provide (S26, S27). All of the model validation figures for all of the different models and datasets can be found in the analysis repository (https://doi.org/10.25405/data.ncl.23815077).

**Table S10**: Redundancy analysis of the variables included in the full versions of the all-cases, separate lineage, and second-wave lineage cases GLMMs. All temporal variables include a two-week time lag. Values are the $R^2$ with which each variable can be predicted from all other variables. Variables with an asterisk were identified as being redundant.

| Variable | All-cases | B.1.1.1 | B.1.1.119 | B.1.1.309 | B.1.1.315 | B.1.1.37 | B.1.1.7 | B.1.177 | B.1.177.10 | Second-wave lineage cases |
|---|---|---|---|---|---|---|---|---|---|---|
| Temperature | 0.319 | 0.408 | 0.103 | 0.748 | 0.794* | 0.682 | 0.738 | 0.794* | 0.585 | 0.794* |
| Rainfall | 0.338 | 0.355 | 0.358 | 0.677 | 0.686 | 0.761 | 0.903 | 0.686 | 0.515 | 0.686 |
| Lockdown 1 | 0.123 | 0.133 | 0.054 | NA | NA | NA | NA | NA | NA | NA |
| Lockdown 2 | 0.168 | 0.252 | NA | 0.592 | 0.672 | 0.666 | 0.941* | 0.672 | 0.706 | 0.672 |
| Tier 2 | 0.06 | 0.102 | NA | 0.455 | 0.363 | 0.311 | 0.778 | 0.363 | 0.382 | 0.363 |
| Tier 3 | 0.136 | 0.219 | NA | NA | 0.566 | NA | 0.807 | 0.566 | 0.587 | 0.566 |
| Eat-out Subsidy | 0.369 | 0.38 | 0.408 | 0.698 | 0.689 | 0.765* | NA | 0.689 | NA | 0.689 |
| IMD 10th Decile | 0.014 | 0.014 | 0.014 | 0.014 | 0.014 | 0.014 | 0.014 | 0.014 | 0.014 | 0.212 |
| Total Population | 0.014 | 0.014 | 0.014 | 0.014 | 0.014 | 0.014 | 0.014 | 0.014 | 0.014 | 0.174 |
| First-wave cases | NA | NA | NA | NA | NA | NA | NA | NA | NA | 0.335 |

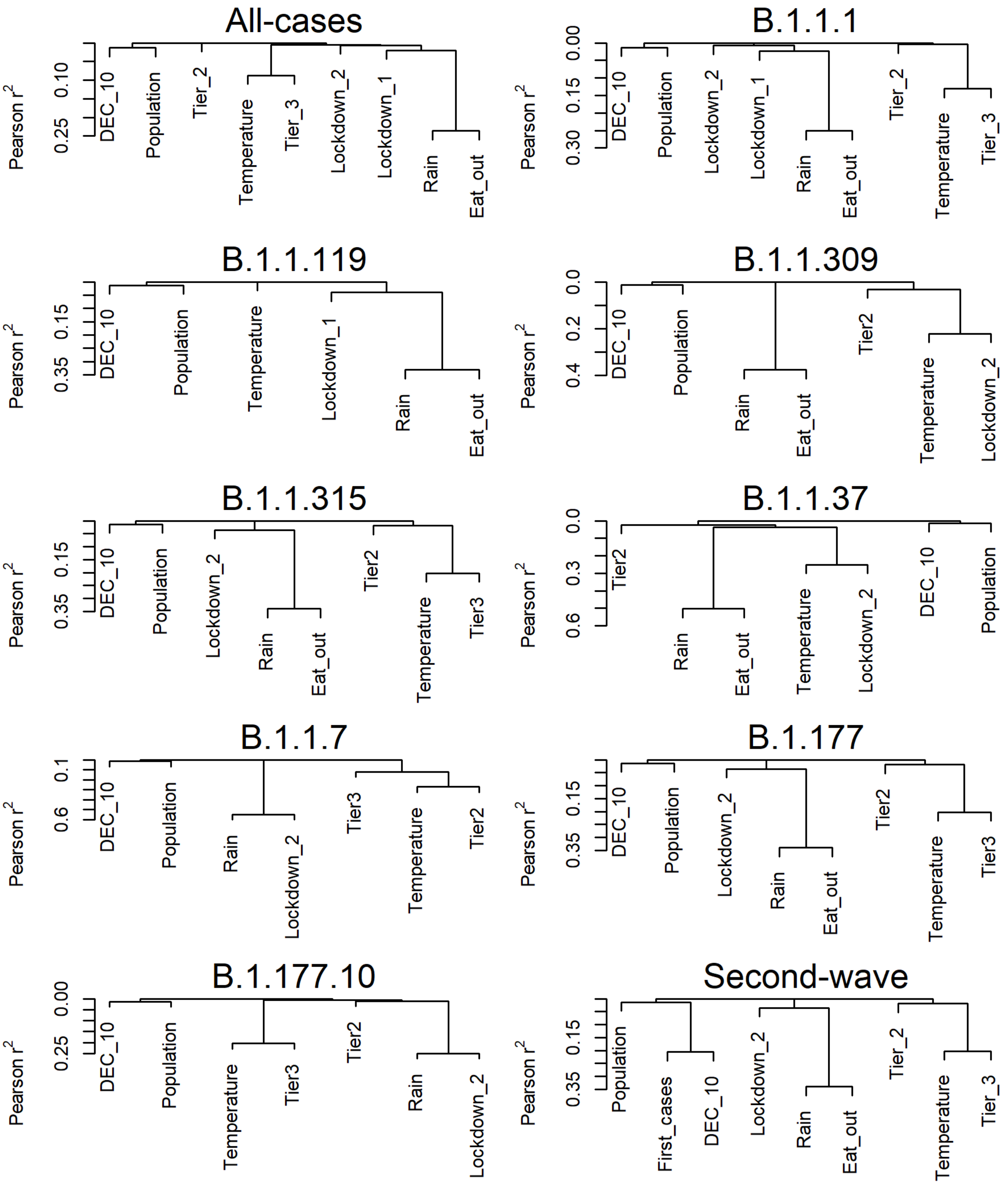

**Figure S23**: Hierarchical cluster analysis plots of the variables included in the full versions of the GLMMs for each dataset, using squared Pearson correlation. Created with the "varclus" function of the Hmisc R package. Absent variables are where they did not overlap temporally with cases of the given lineage.

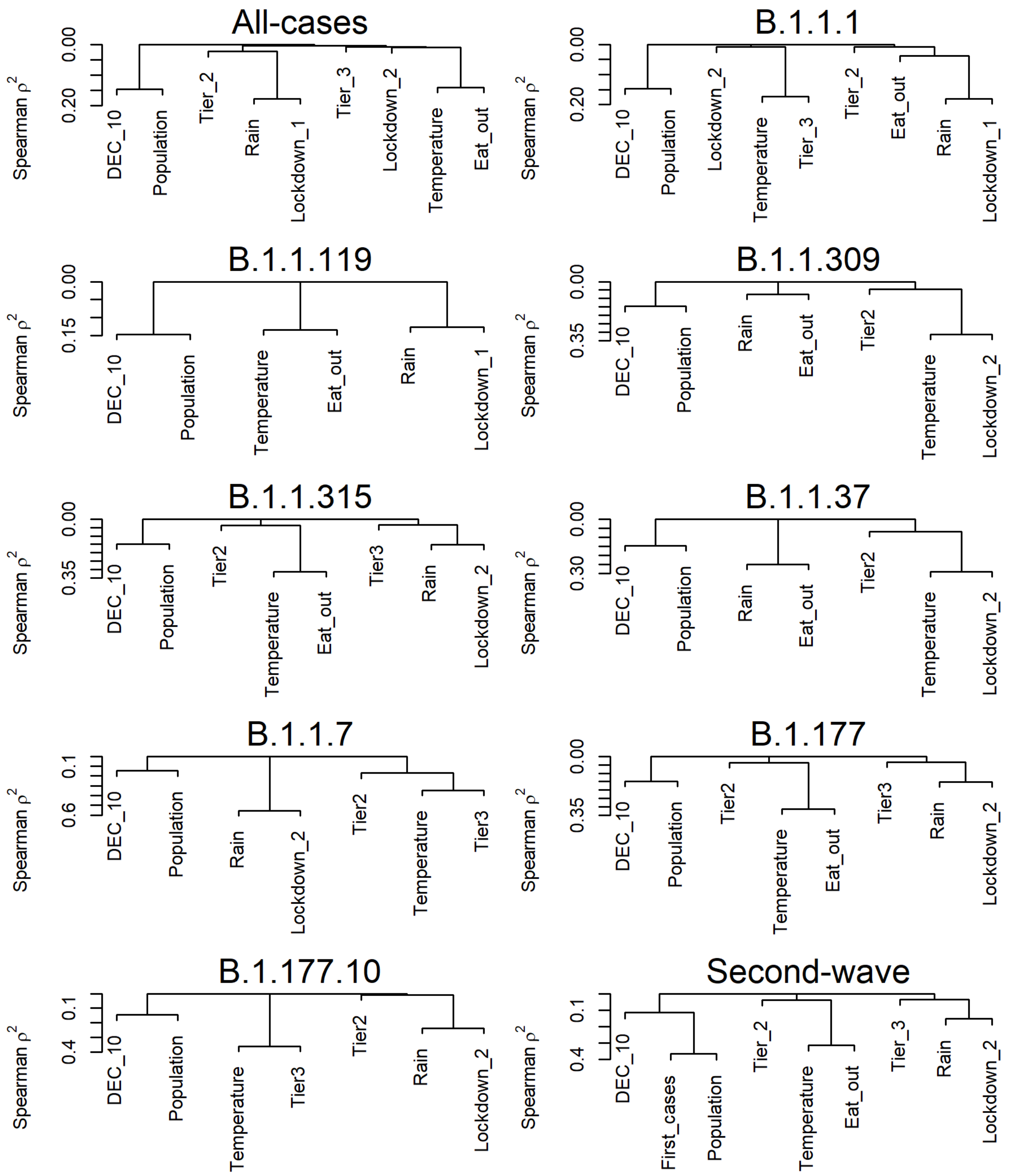

**Figure S24**: Hierarchical cluster analysis plots of the variables included in the full versions of the GLMMs for each dataset, using squared Spearman correlation. Created with the “varclus” function of the Hmisc R package. Absent variables are where they did not overlap temporally with cases of the given lineage.

#### All-cases - AR1 term for week (grouped by postcode)

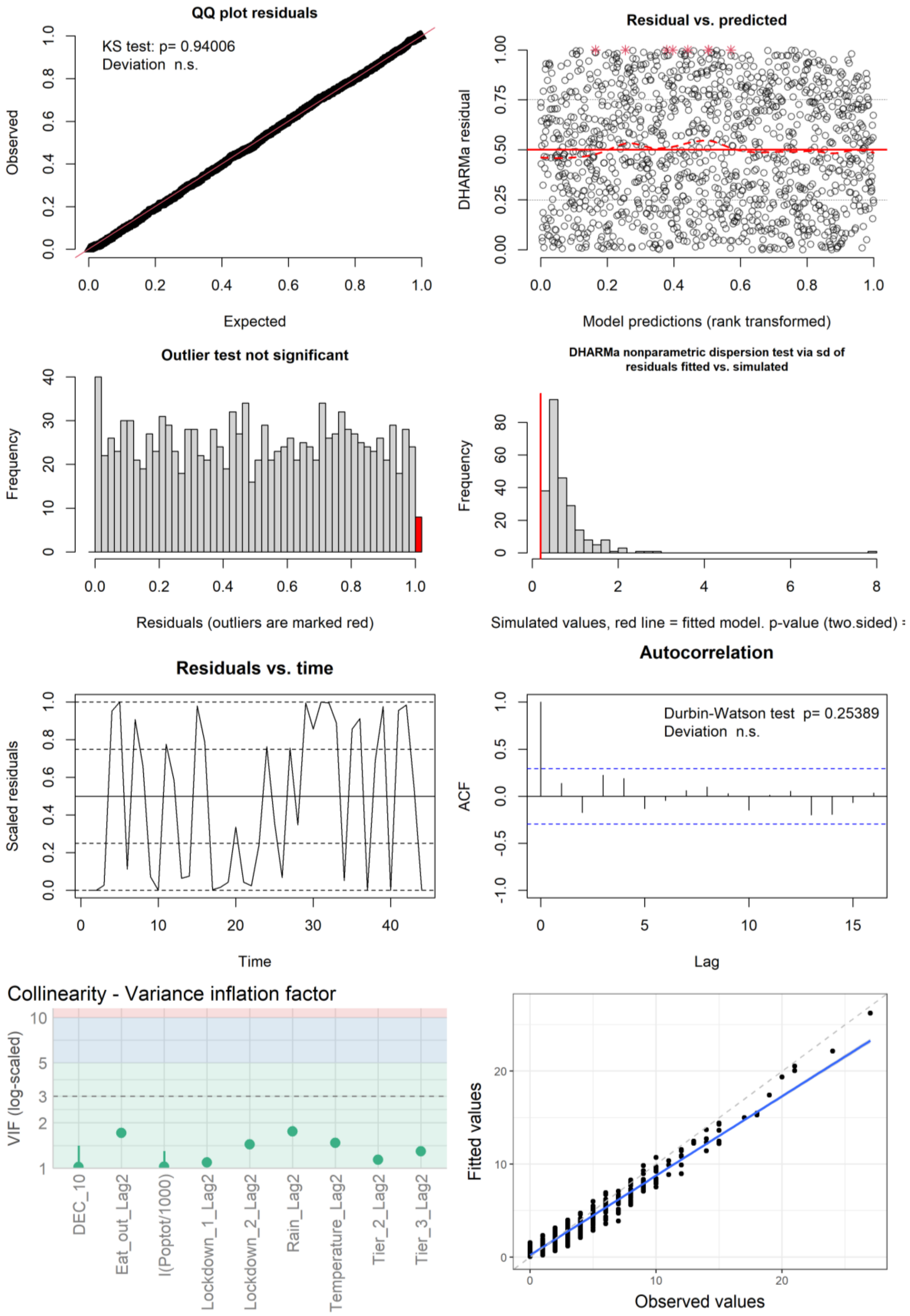

**Figure S25**: Figures used to assess model fit for the all-cases GLMM with an AR1 term for week grouped by postcode. First row: QQ plot, residuals vs predicted. Second row: outlier check, dispersion check. Third row: residuals vs time, ACF. Bottom row: VIF, observed values vs fitted values from the model.

#### All-cases - Restricted cubic spline for time (4 knots)

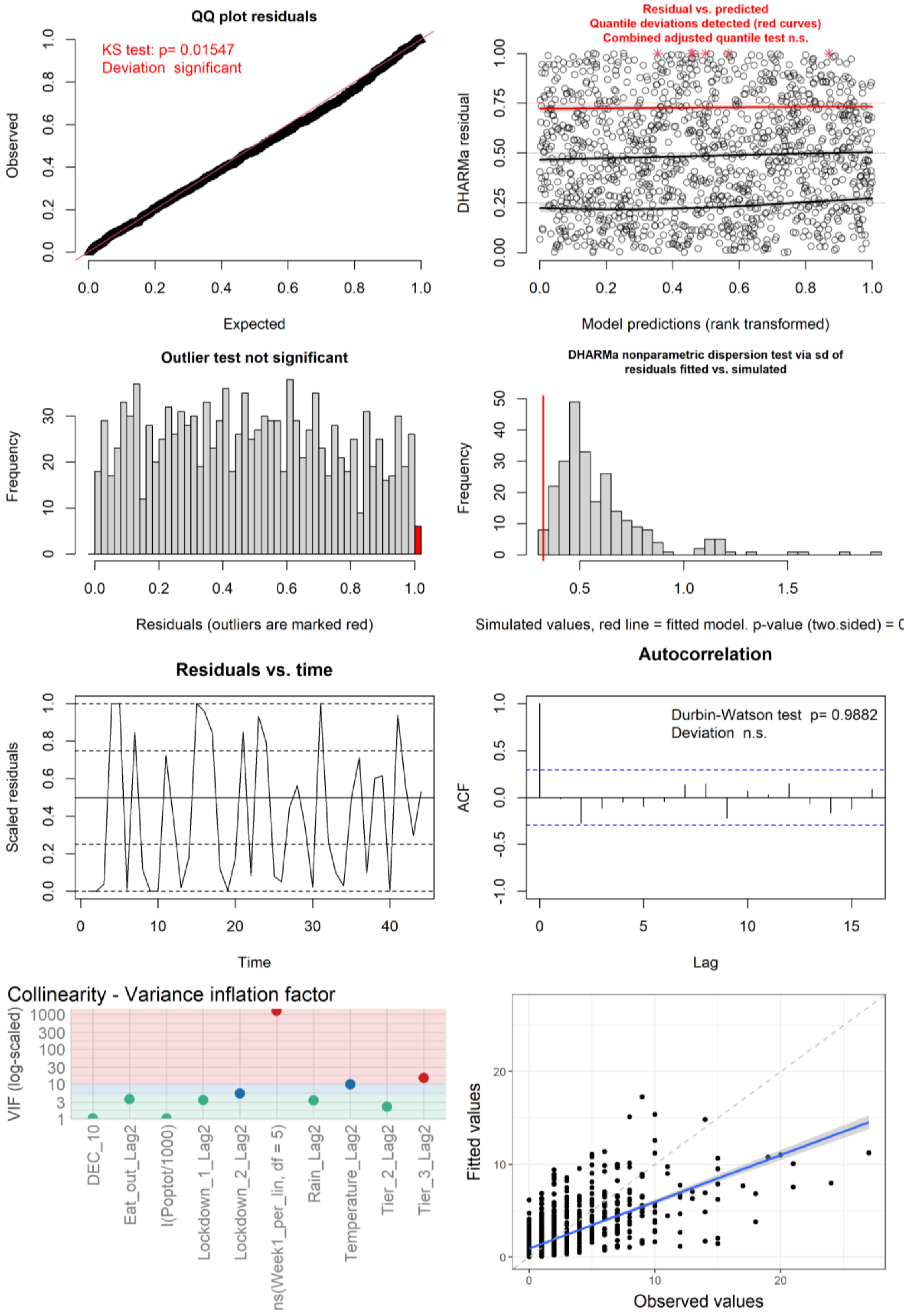

**Figure S26**: Figures used to assess model fit for the all-cases GLMM with a restricted cubic spline for week with 4 knots. First row: QQ plot, residuals vs predicted. Second row: outlier check, dispersion check. Third row: residuals vs time, ACF. Bottom row: VIF, observed values vs fitted values from the model.

#### All-cases - Smooth spline for time

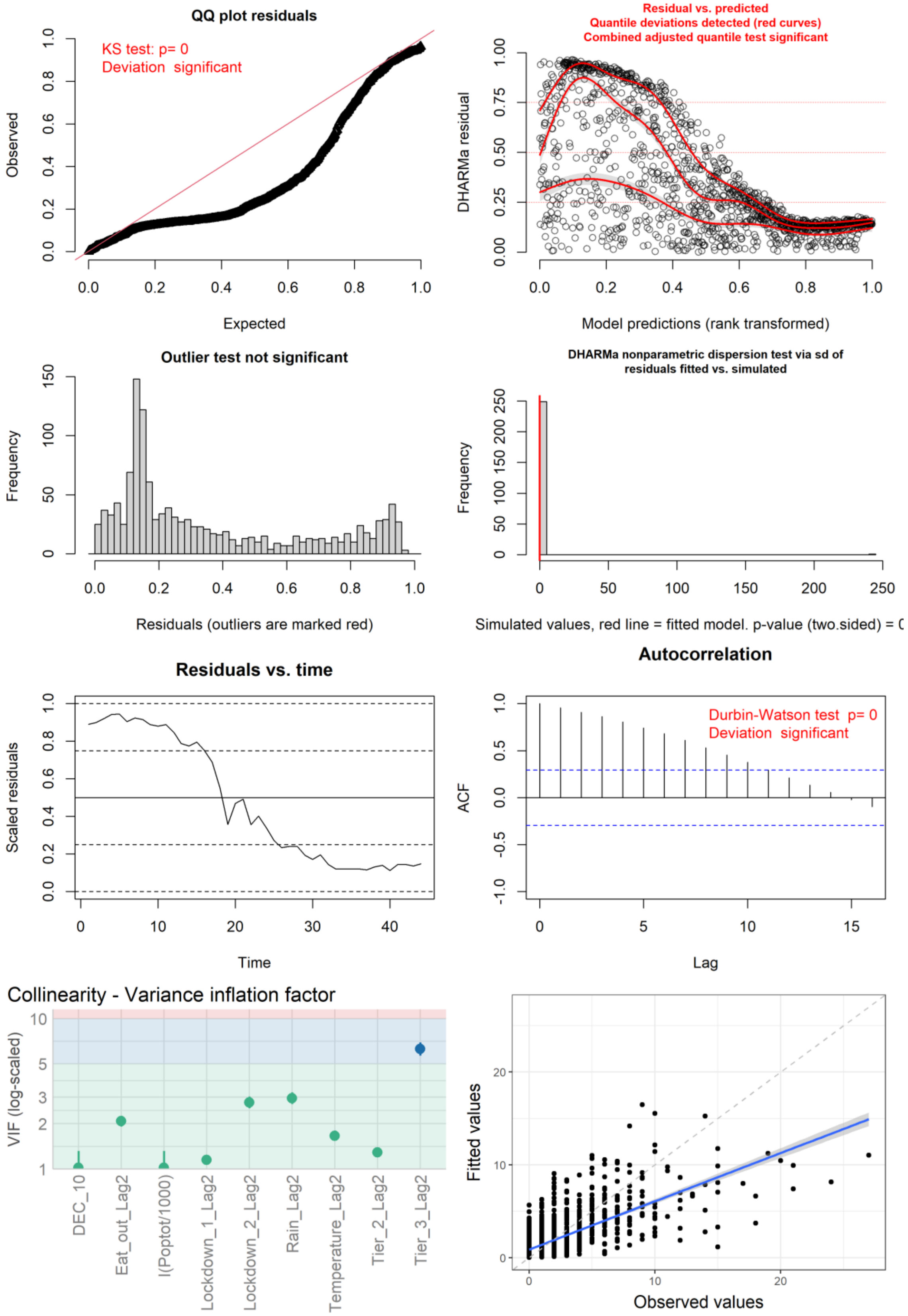

**Figure S27**: Figures used to assess model fit for the all-cases GLMM with a smooth spline for week. First row: QQ plot, residuals vs predicted. Second row: outlier check, dispersion check. Third row: residuals vs time, ACF. Bottom row: VIF, observed values vs fitted values from the model.

#### Lineage B.1.1.1 - AR1 term for week (grouped by postcode)

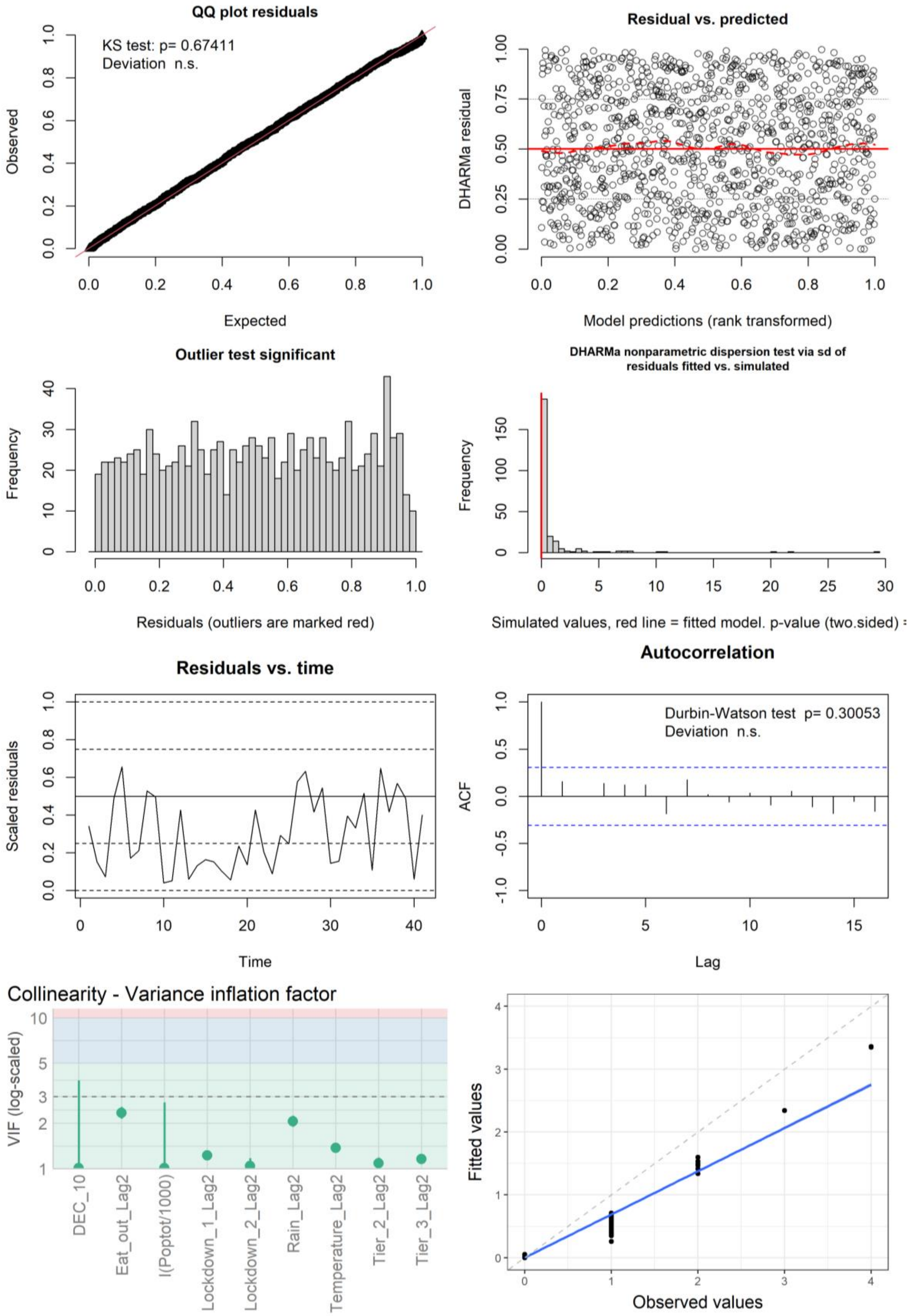

**Figure S28**: Figures used to assess model fit for the lineage B.1.1.1 GLMM with an AR1 term for week grouped by postcode. First row: QQ plot, residuals vs predicted. Second row: outlier check, dispersion check. Third row: residuals vs time, ACF. Bottom row: VIF, observed values vs fitted values from the model.

#### Lineage B.1.1.119 - AR1 term for week (grouped by postcode)

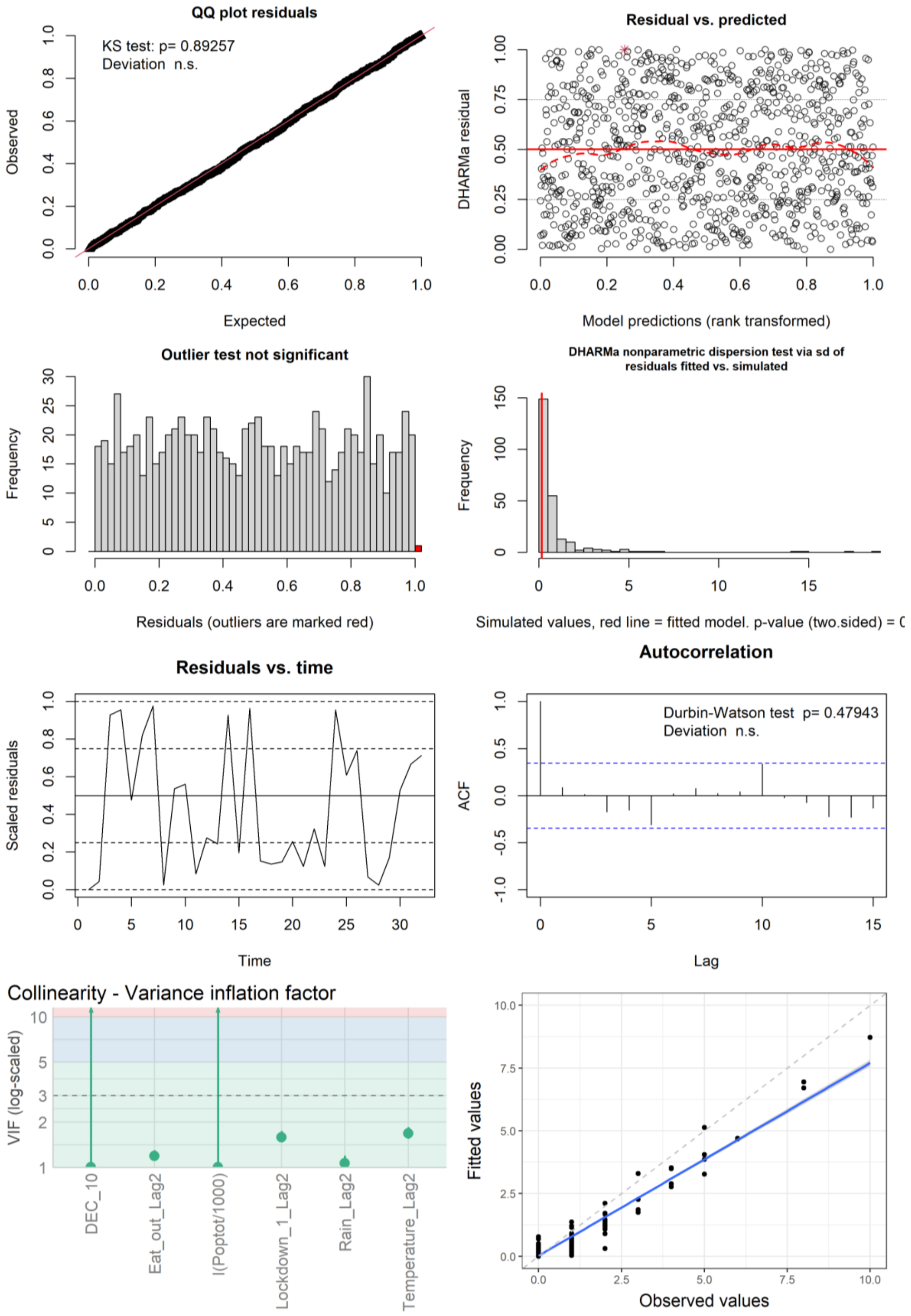

**Figure S29**: Figures used to assess model fit for the lineage B.1.1.119 GLMM with an AR1 term for week grouped by postcode. First row: QQ plot, residuals vs predicted. Second row: outlier check, dispersion check. Third row: residuals vs time, ACF. Bottom row: VIF, observed values vs fitted values from the model.

#### Lineage B.1.1.309 - AR1 term for week (grouped by postcode)

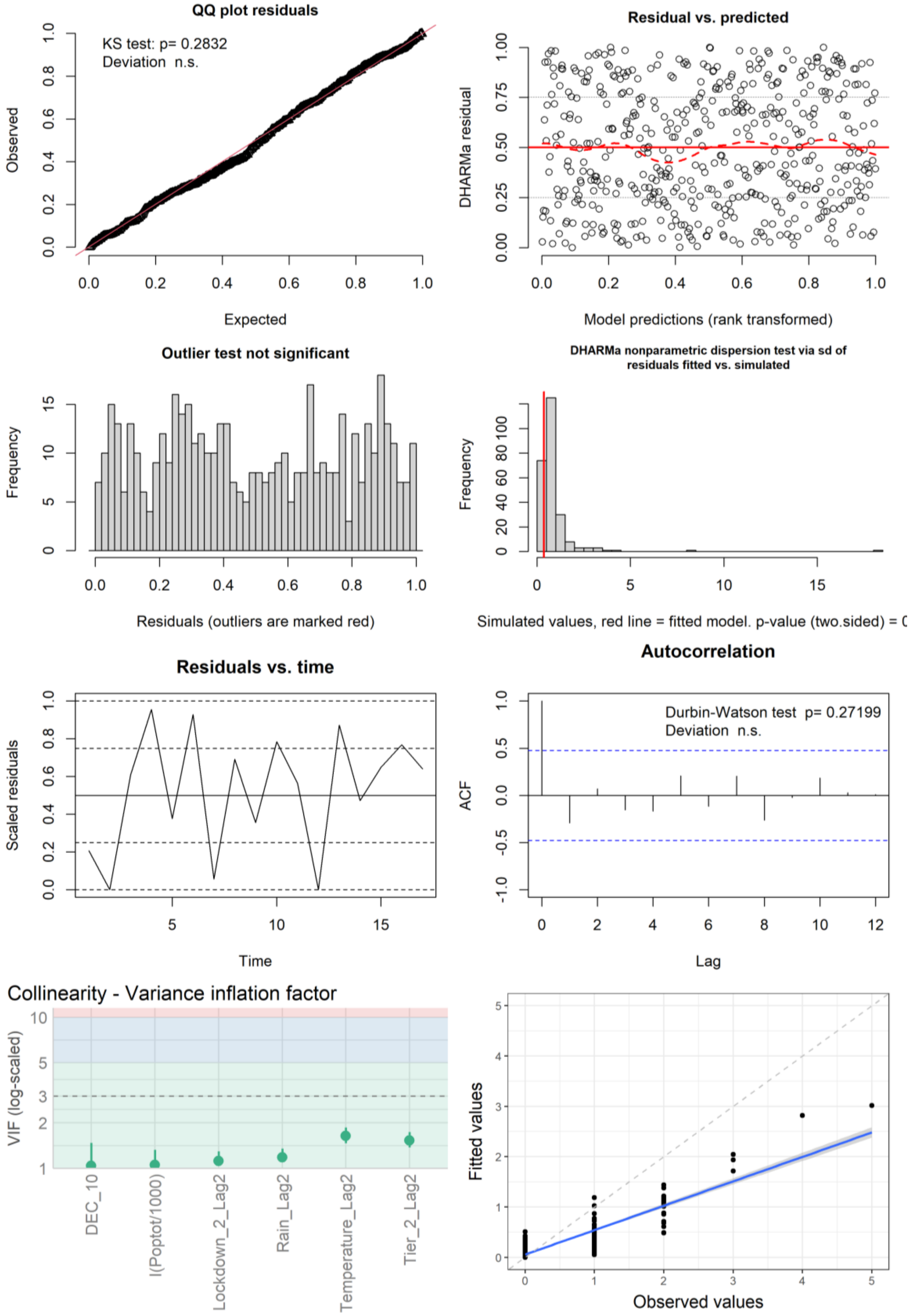

**Figure S30**: Figures used to assess model fit for the lineage B.1.1.309 GLMM with an AR1 term for week grouped by postcode. Eat-out subsidy was dropped from this final model due to collinearity. The random intercept for postcode was dropped due to poorer model fit. First row: QQ plot, residuals vs predicted. Second row: outlier check, dispersion check. Third row: residuals vs time, ACF. Bottom row: VIF, observed values vs fitted values from the model.

#### Lineage B.1.1.315 - AR1 term for week (grouped by postcode)

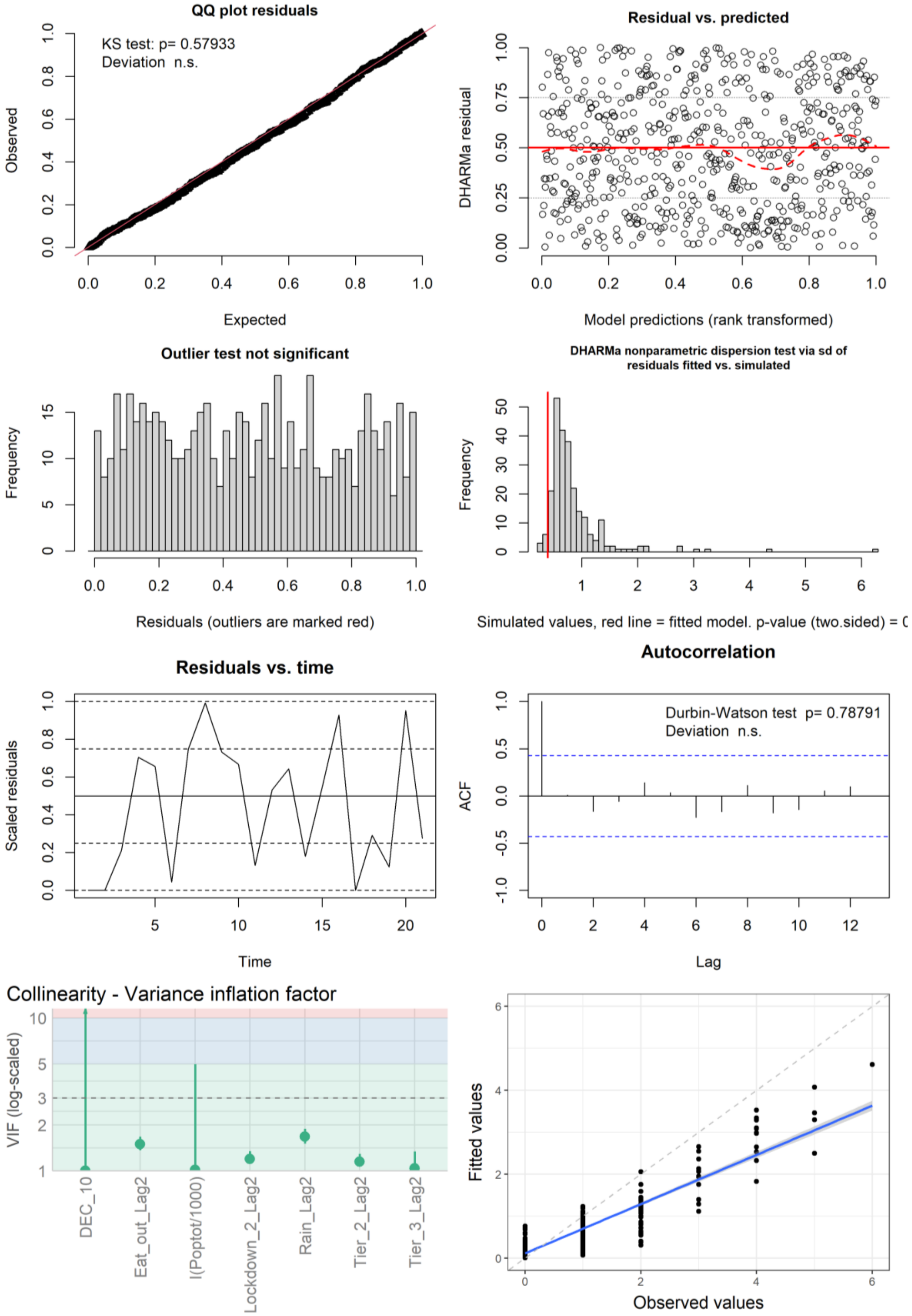

**Figure S31**: Figures used to assess model fit for the lineage B.1.1.315 GLMM with an AR1 term for week grouped by postcode. Temperature was dropped from this final model due to collinearity. First row: QQ plot, residuals vs predicted. Second row: outlier check, dispersion check. Third row: residuals vs time, ACF. Bottom row: VIF, observed values vs fitted values from the model.

#### Lineage B.1.1.37 - AR1 term for week (grouped by postcode)

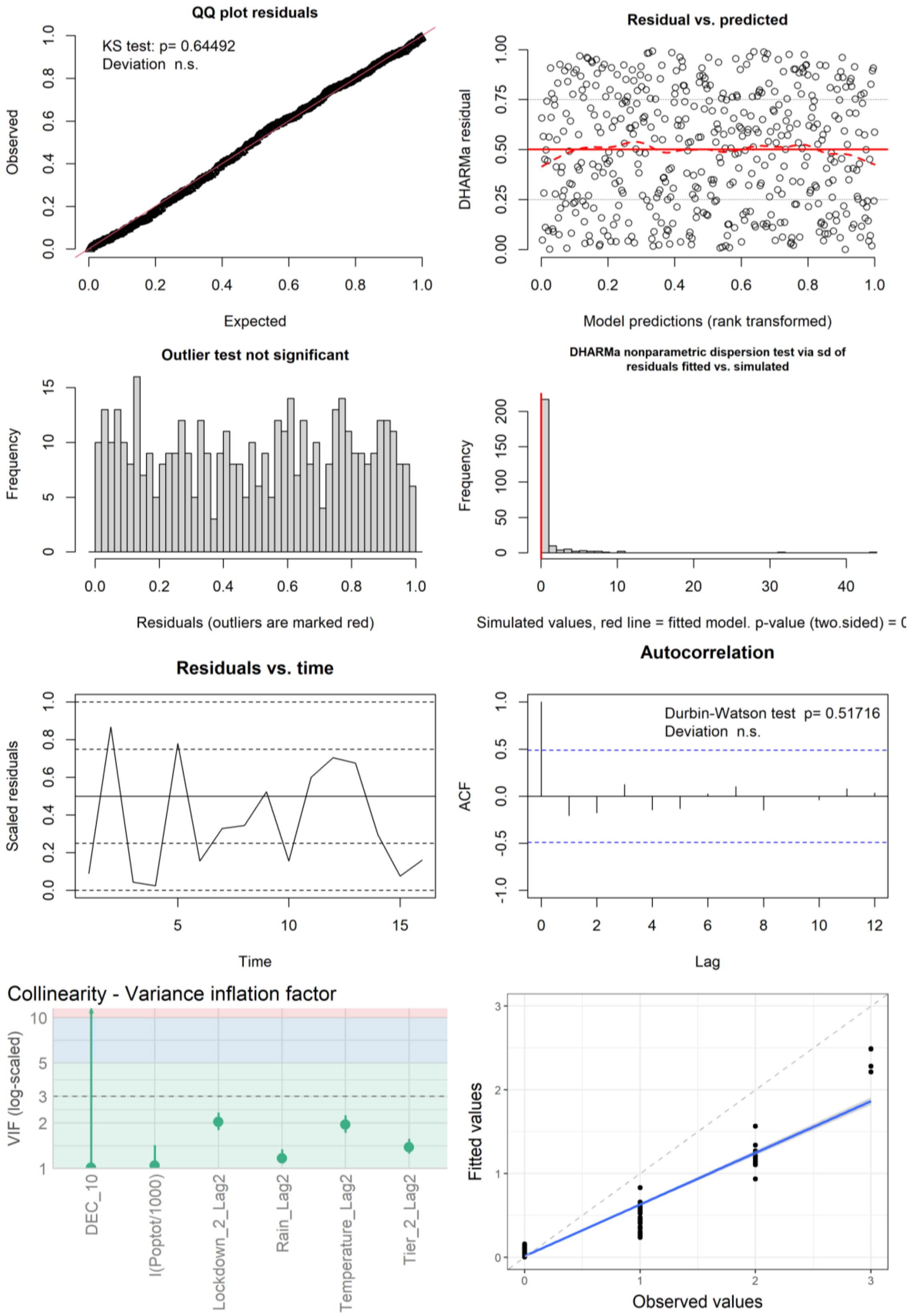

**Figure S32**: Figures used to assess model fit for the lineage B.1.1.37 GLMM with an AR1 term for week grouped by postcode. The eat-out subsidy was dropped from this final model due to collinearity. First row: QQ plot, residuals vs predicted. Second row: outlier check, dispersion check. Third row: residuals vs time, ACF. Bottom row: VIF, observed values vs fitted values from the model.

#### Lineage B.1.1.7 - AR1 term for week (grouped by postcode)

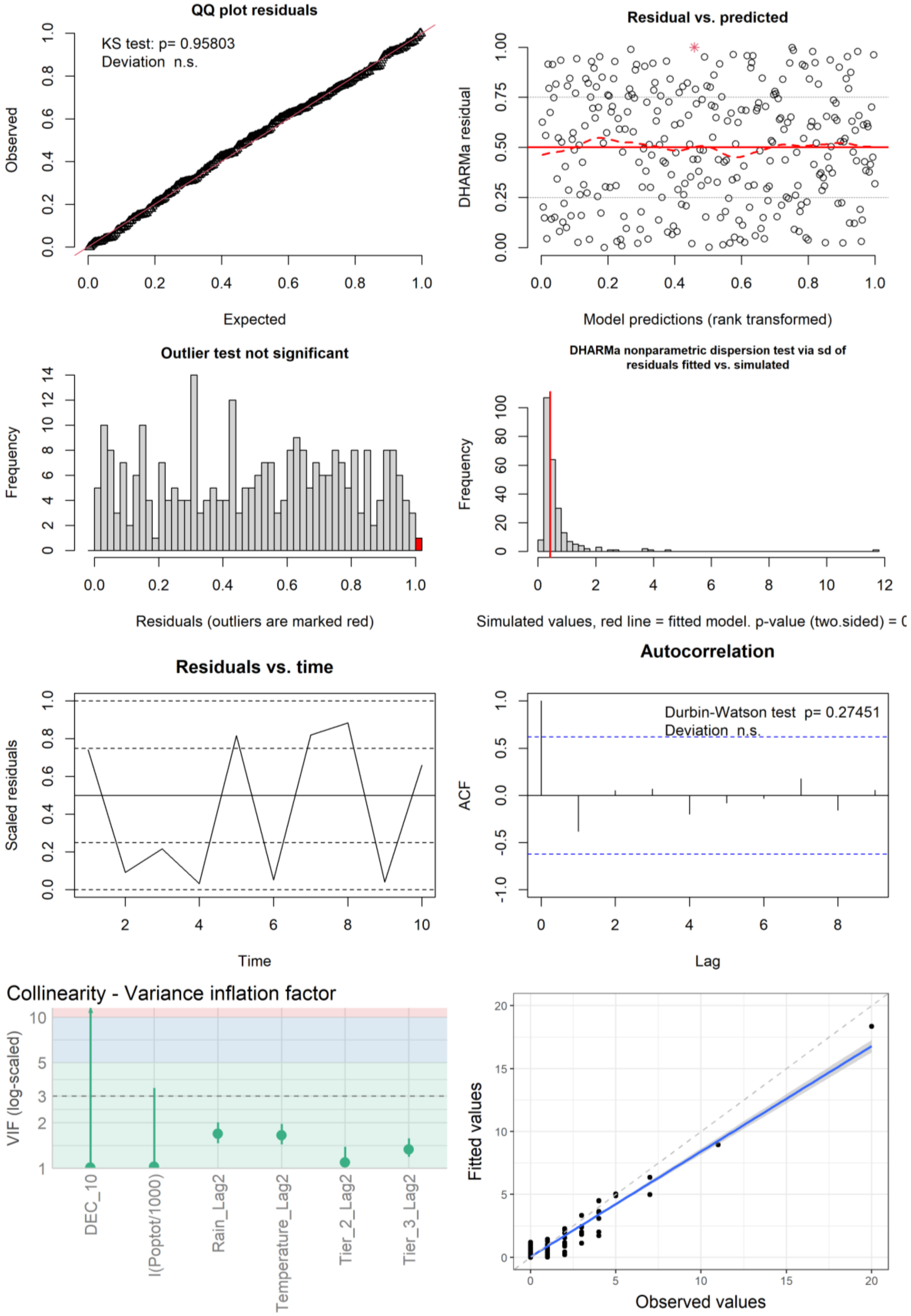

**Figure S33**: Figures used to assess model fit for the lineage B.1.1.7 GLMM with an AR1 term for week grouped by postcode. Lockdown 2 was dropped from this final model due to collinearity. First row: QQ plot, residuals vs predicted. Second row: outlier check, dispersion check. Third row: residuals vs time, ACF. Bottom row: VIF, observed values vs fitted values from the model.

#### Lineage B.1.177 - AR1 term for week (grouped by postcode)

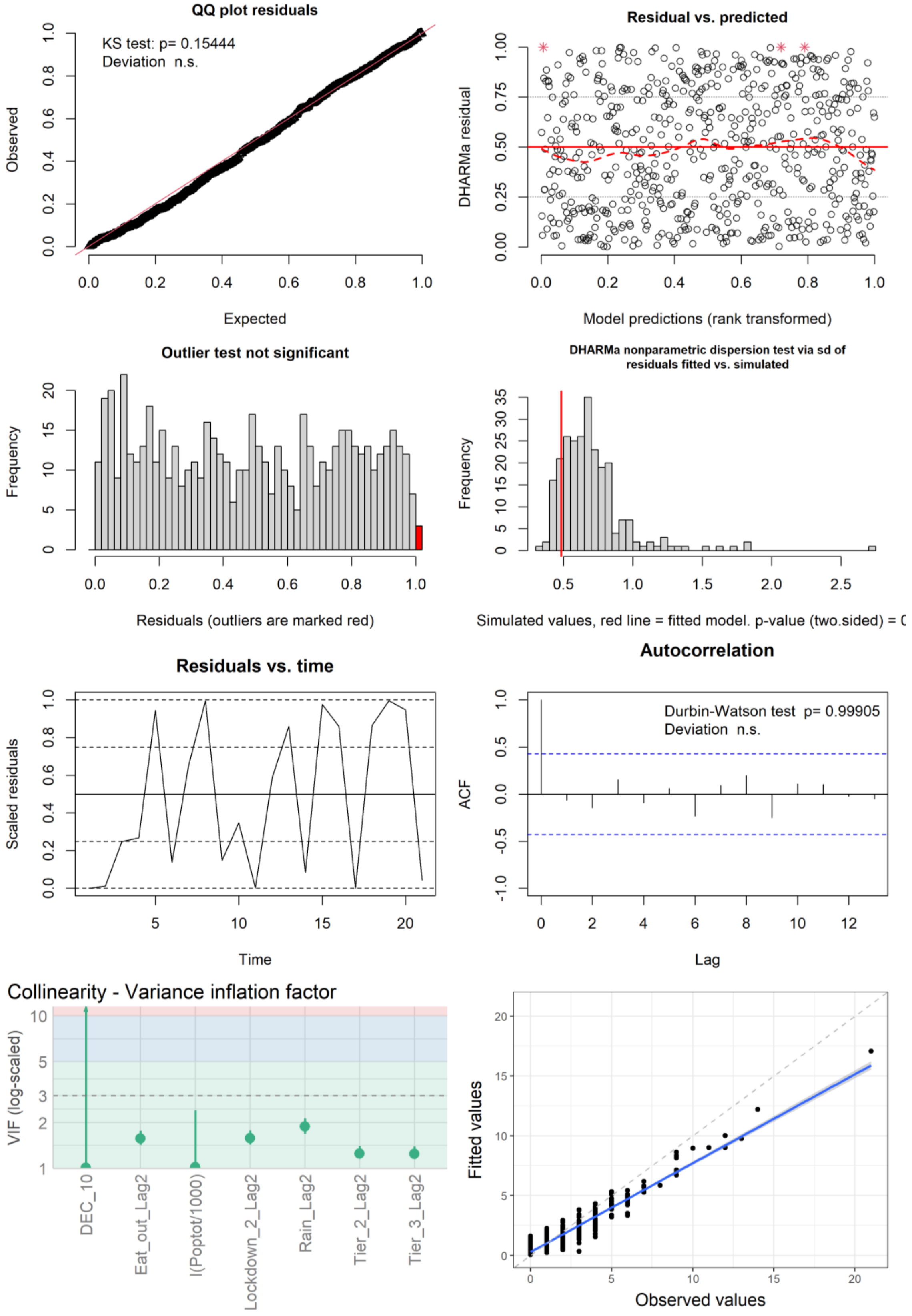

**Figure S34**: Figures used to assess model fit for the lineage B.1.177 GLMM with an AR1 term for week grouped by postcode. Temperature was dropped from this final model due to collinearity. First row: QQ plot, residuals vs predicted. Second row: outlier check, dispersion check. Third row: residuals vs time, ACF. Bottom row: VIF, observed values vs fitted values from the model.

#### Lineage B.1.177.10 - AR1 term for week (grouped by postcode)

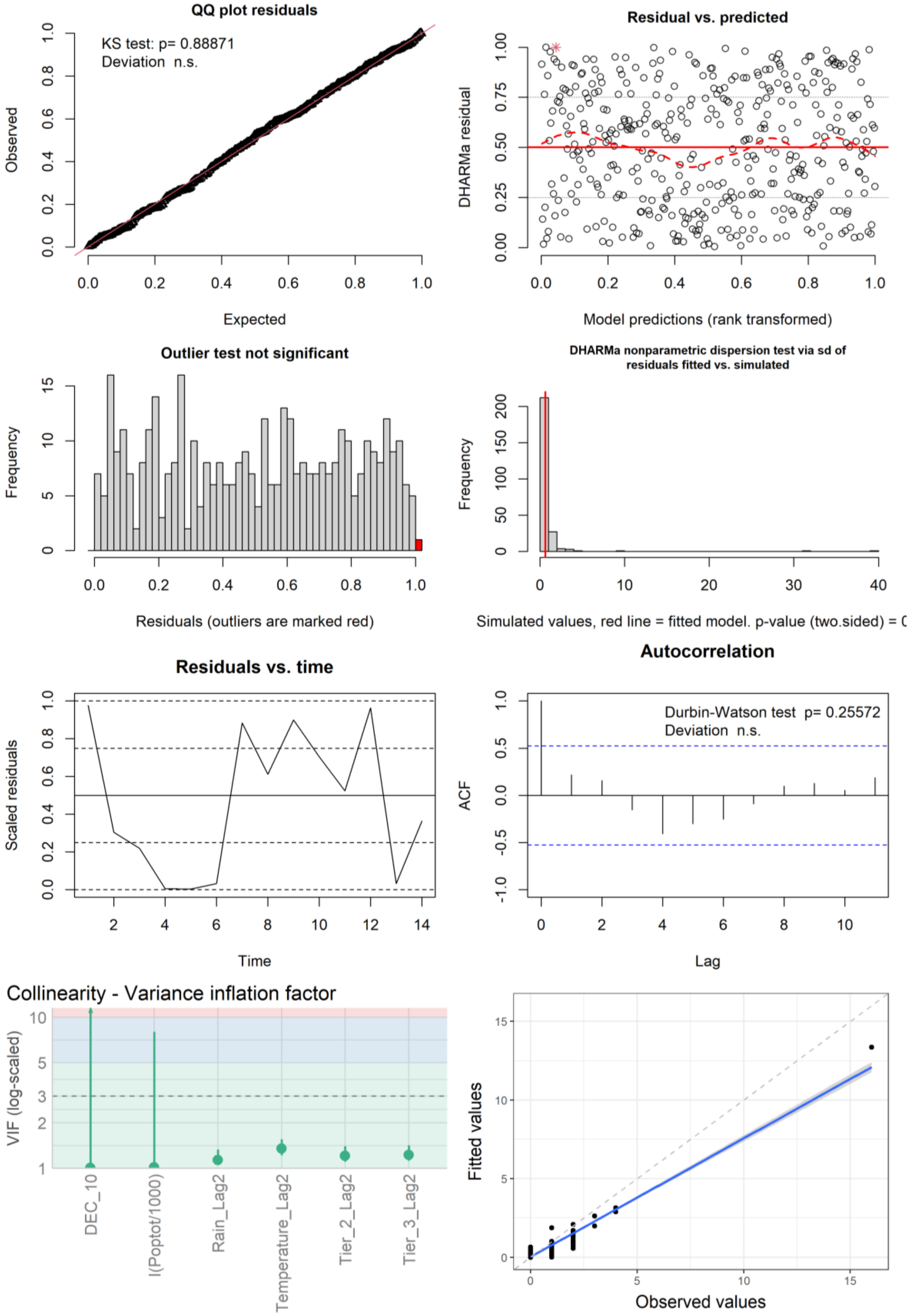

**Figure S35**: Figures used to assess model fit for the lineage B.1.177.10 GLMM with an AR1 term for week grouped by postcode. Lockdown 2 was dropped from this final model due to collinearity. First row: QQ plot, residuals vs predicted. Second row: outlier check, dispersion check. Third row: residuals vs time, ACF. Bottom row: VIF, observed values vs fitted values from the model.

#### Second-wave lineage cases - AR1 term for week (grouped by postcode)

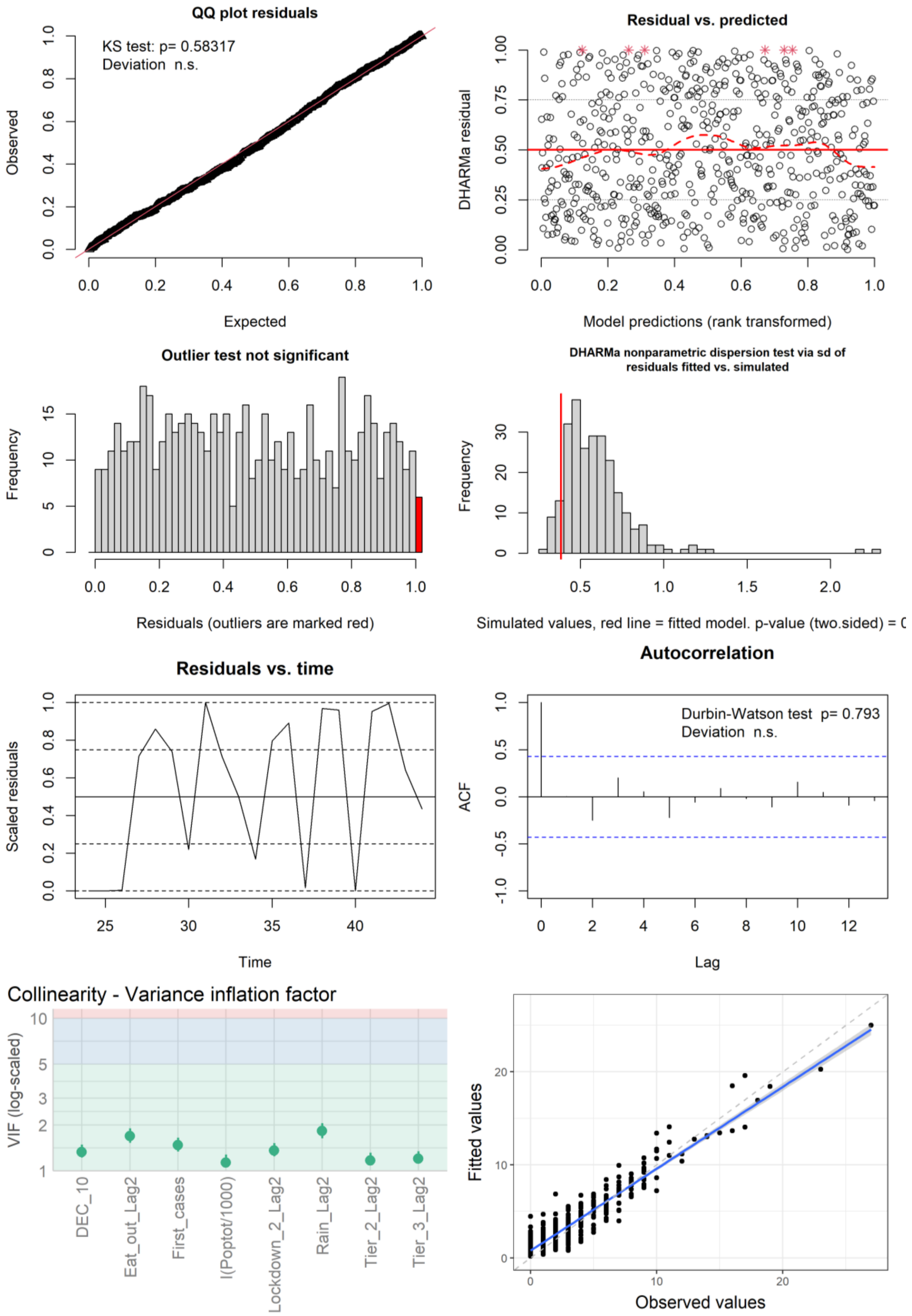

**Figure S36**: Figures used to assess model fit second-wave lineage cases GLMM with an AR1 term for week grouped by postcode. Temperature was dropped from this final model due to collinearity. First row: QQ plot, residuals vs predicted. Second row: outlier check, dispersion check. Third row: residuals vs time, ACF. Bottom row: VIF, observed values vs fitted values from the model.

#### All-cases - Random gradient for week for each postcode

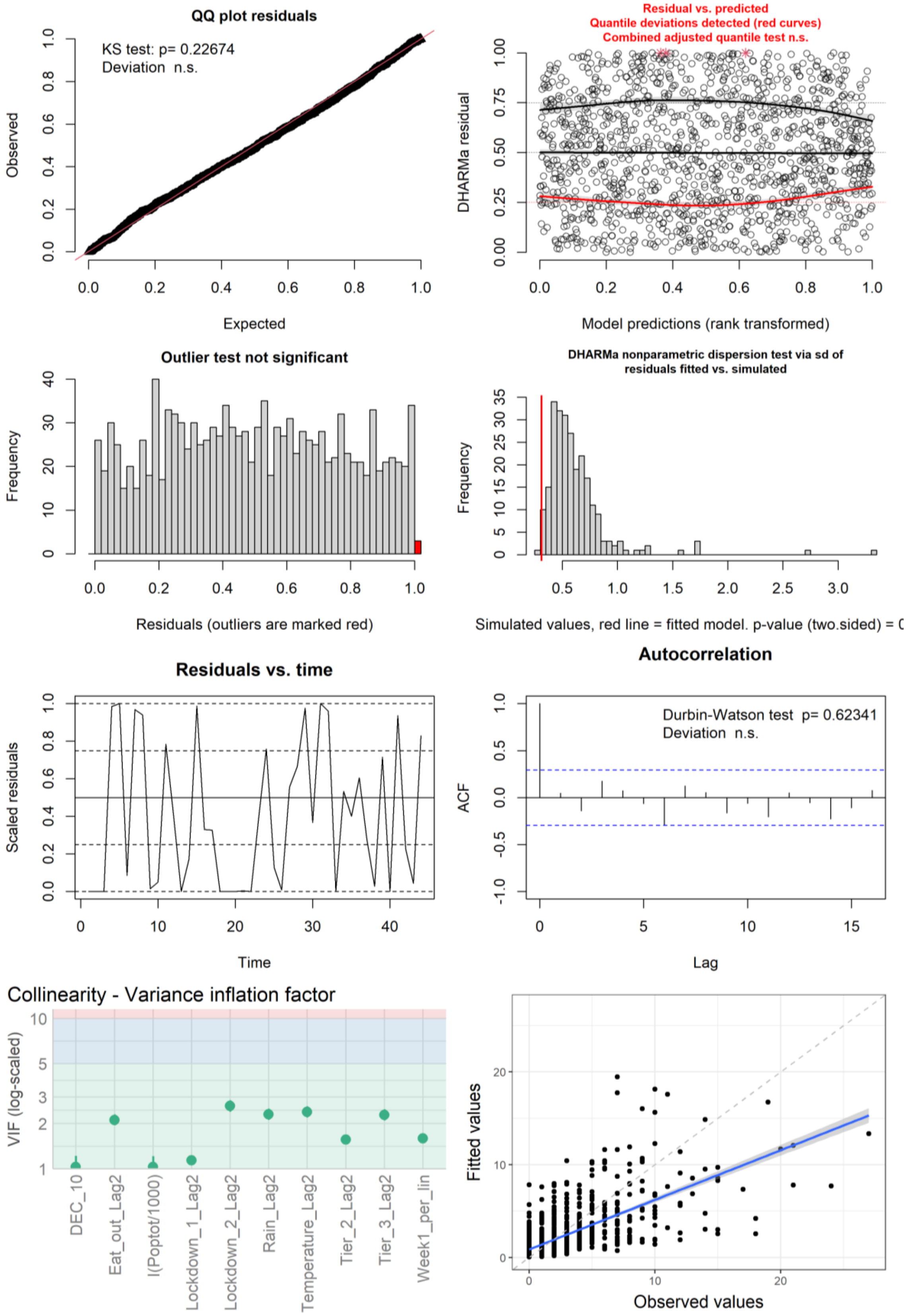

**Figure S37**: Figures used to assess model fit all-cases GLMM with a random gradient for week for each postcode. First row: QQ plot, residuals vs predicted. Second row: outlier check, dispersion check. Third row: residuals vs time, ACF. Bottom row: VIF, observed values vs fitted values from the model.

#### 4. Full List of COG Consortium Members

**Funding acquisition, Leadership and supervision, Metadata curation, Project administration, Samples and logistics, Sequencing and analysis, Software and analysis tools, and Visualisation:**
Dr Samuel C Robson PhD [13, 84]

**Funding acquisition, Leadership and supervision, Metadata curation, Project administration, Samples and logistics, Sequencing and analysis, and Software and analysis tools:**
Dr Thomas R Connor PhD [11, 74] and Prof Nicholas J Loman PhD [43]

**Leadership and supervision, Metadata curation, Project administration, Samples and logistics, Sequencing and analysis, Software and analysis tools, and Visualisation:**
Dr Tanya Golubchik PhD [5]

**Funding acquisition, Leadership and supervision, Metadata curation, Samples and logistics, Sequencing and analysis, and Visualisation:**
Dr Rocio T Martinez Nunez PhD [46]

**Funding acquisition, Leadership and supervision, Project administration, Samples and logistics, Sequencing and analysis, and Software and analysis tools:**
Dr David Bonsall PhD [5]

**Funding acquisition, Leadership and supervision, Project administration, Sequencing and analysis, Software and analysis tools, and Visualisation:**
Prof Andrew Rambaut DPhil [104]

**Funding acquisition, Metadata curation, Project administration, Samples and logistics, Sequencing and analysis, and Software and analysis tools:**
Dr Luke B Snell MSc, MBBS [12]

**Leadership and supervision, Metadata curation, Project administration, Samples and logistics, Software and analysis tools, and Visualisation:**
Rich Livett MSc [116]

**Funding acquisition, Leadership and supervision, Metadata curation, Project administration, and Samples and logistics:**
Dr Catherine Ludden PhD [20, 70]

**Funding acquisition, Leadership and supervision, Metadata curation, Samples and logistics, and Sequencing and analysis:**
Dr Sally Corden PhD [74] and Dr Eleni Nastouli FRCPath [96, 95, 30]

**Funding acquisition, Leadership and supervision, Metadata curation, Sequencing and analysis, and Software and analysis tools:**
Dr Gaia Nebbia PhD, FRCPath [12]

**Funding acquisition, Leadership and supervision, Project administration, Samples and logistics, and Sequencing and analysis:**
Ian Johnston BSc [116]

**Leadership and supervision, Metadata curation, Project administration, Samples and logistics, and Sequencing and analysis:**

Prof Katrina Lythgoe PhD [5], Dr M. Estee Torok FRCP [19, 20] and Prof Ian G Goodfellow PhD [24]
**Leadership and supervision, Metadata curation, Project administration, Samples and logistics, and Visualisation:**
Dr Jacqui A Prieto PhD [97, 82] and Dr Kordo Saeed MD, FRCPath [97, 83]

**Leadership and supervision, Metadata curation, Project administration, Sequencing and analysis, and Software and analysis tools:**
Dr David K Jackson PhD [116]

**Leadership and supervision, Metadata curation, Samples and logistics, Sequencing and analysis, and Visualisation:**
Dr Catherine Houlihan PhD [96, 94]

**Leadership and supervision, Metadata curation, Sequencing and analysis, Software and analysis tools, and Visualisation:**
Dr Dan Frampton PhD [94, 95]

**Metadata curation, Project administration, Samples and logistics, Sequencing and analysis, and Software and analysis tools:**
Dr William L Hamilton PhD [19] and Dr Adam A Witney PhD [41]

**Funding acquisition, Samples and logistics, Sequencing and analysis, and Visualisation:**
Dr Giselda Bucca PhD [101]

**Funding acquisition, Leadership and supervision, Metadata curation, and Project administration:**
Dr Cassie F Pope PhD[40, 41]

**Funding acquisition, Leadership and supervision, Metadata curation, and Samples and logistics:**
Dr Catherine Moore PhD [74]

**Funding acquisition, Leadership and supervision, Metadata curation, and Sequencing and analysis:**
Prof Emma C Thomson PhD, FRCP [53]

**Funding acquisition, Leadership and supervision, Project administration, and Samples and logistics:**
Dr Ewan M Harrison PhD [116, 102]

**Funding acquisition, Leadership and supervision, Sequencing and analysis, and Visualisation:**
Prof Colin P Smith PhD [101]

**Leadership and supervision, Metadata curation, Project administration, and Sequencing and analysis:**
Fiona Rogan BSc [77]

**Leadership and supervision, Metadata curation, Project administration, and Samples and logistics:**
Shaun M Beckwith MSc [6], Abigail Murray Degree [6], Dawn Singleton HNC [6], Dr Kirstine Eastick PhD, FRCPath [37], Dr Liz A Sheridan PhD [98], Paul Randell MSc, PgD [99], Dr Leigh M Jackson PhD [105], Dr Cristina V Ariani PhD [116] and Dr Sónia Gonçalves PhD [116]

**Leadership and supervision, Metadata curation, Samples and logistics, and Sequencing and analysis:**
Dr Derek J Fairley PhD [3, 77], Prof Matthew W Loose PhD [18] and Joanne Watkins MSc [74]

**Leadership and supervision, Metadata curation, Samples and logistics, and Visualisation:**
Dr Samuel Moses MD [25, 106]

**Leadership and supervision, Metadata curation, Sequencing and analysis, and Software and analysis tools:**
Dr Sam Nicholls PhD [43], Dr Matthew Bull PhD [74] and Dr Roberto Amato PhD [116]

**Leadership and supervision, Project administration, Samples and logistics, and Sequencing and analysis:**
Prof Darren L Smith PhD [36, 65, 66]

**Leadership and supervision, Sequencing and analysis, Software and analysis tools, and Visualisation:**
Prof David M Aanensen PhD [14, 116] and Dr Jeffrey C Barrett PhD [116]

**Metadata curation, Project administration, Samples and logistics, and Sequencing and analysis:**
Dr Dinesh Aggarwal MRCP[20, 116, 70], Dr James G Shepherd MBCHB, MRCP [53], Dr Martin D Curran PhD [71] and Dr Surendra Parmar PhD [71]

**Metadata curation, Project administration, Sequencing and analysis, and Software and analysis tools:**
Dr Matthew D Parker PhD [109]

**Metadata curation, Samples and logistics, Sequencing and analysis, and Software and analysis tools:**
Dr Catryn Williams PhD [74]

**Metadata curation, Samples and logistics, Sequencing and analysis, and Visualisation:**
Dr Sharon Glaysher PhD [68]

**Metadata curation, Sequencing and analysis, Software and analysis tools, and Visualisation:**
Dr Anthony P Underwood PhD [14, 116], Dr Matthew Bashton PhD [36, 65], Dr Nicole Pacchiarini PhD [74], Dr Katie F Loveson PhD [84] and Matthew Byott MSc [95, 96]

**Project administration, Sequencing and analysis, Software and analysis tools, and Visualisation:**
Dr Alessandro M Carabelli PhD [20]

**Funding acquisition, Leadership and supervision, and Metadata curation:**
Dr Kate E Templeton PhD [56, 104]

**Funding acquisition, Leadership and supervision, and Project administration:**
Dr Thushan I de Silva PhD [109], Dr Dennis Wang PhD [109], Dr Cordelia F Langford PhD [116] and John Sillitoe BEng [116]

**Funding acquisition, Leadership and supervision, and Samples and logistics:**
Prof Rory N Gunson PhD, FRCPath [55]

**Funding acquisition, Leadership and supervision, and Sequencing and analysis:**
Dr Simon Cottrell PhD [74], Dr Justin O'Grady PhD [75, 103] and Prof Dominic Kwiatkowski PhD [116, 108]

**Leadership and supervision, Metadata curation, and Project administration:**
Dr Patrick J Lillie PhD, FRCP [37]

**Leadership and supervision, Metadata curation, and Samples and logistics:**
Dr Nicholas Cortes MBCHB [33], Dr Nathan Moore MBCHB [33], Dr Claire Thomas DPhil [33], Phillipa J Burns MSc, DipRCPath [37], Dr Tabitha W Mahungu FRCPath [80] and Steven Liggett BSc [86]

**Leadership and supervision, Metadata curation, and Sequencing and analysis:**
Angela H Beckett MSc [13, 81] and Prof Matthew TG Holden PhD [73]

**Leadership and supervision, Project administration, and Samples and logistics:**

Dr Lisa J Levett PhD [34], Dr Husam Osman PhD [70, 35] and Dr Mohammed Hassan-Ibrahim PhD, FRCPath [99]

**Leadership and supervision, Project administration, and Sequencing and analysis:**
Dr David A Simpson PhD [77]

**Leadership and supervision, Samples and logistics, and Sequencing and analysis:**
Dr Meera Chand PhD [72], Prof Ravi K Gupta PhD [102], Prof Alistair C Darby PhD [107] and Prof Steve Paterson PhD [107]

**Leadership and supervision, Sequencing and analysis, and Software and analysis tools:**
Prof Oliver G Pybus DPhil [23], Dr Erik M Volz PhD [39], Prof Daniela de Angelis PhD [52], Prof David L Robertson PhD [53], Dr Andrew J Page PhD [75] and Dr Inigo Martincorena PhD [116]

**Leadership and supervision, Sequencing and analysis, and Visualisation:**
Dr Louise Aigrain PhD [116] and Dr Andrew R Bassett PhD [116]

**Metadata curation, Project administration, and Samples and logistics:**
Dr Nick Wong DPhil, MRCP, FRCPath [50], Dr Yusri Taha MD, PhD [89], Michelle J Erkiert BA [99] and Dr Michael H Spencer Chapman MBBS [116, 102]

**Metadata curation, Project administration, and Sequencing and analysis:**
Dr Rebecca Dewar PhD [56] and Martin P McHugh MSc [56, 111]

**Metadata curation, Project administration, and Software and analysis tools:**
Siddharth Mookerjee MPH [38, 57]

**Metadata curation, Project administration, and Visualisation:**
Stephen Aplin [97], Matthew Harvey [97], Thea Sass [97], Dr Helen Umpleby FRCP [97] and Helen Wheeler [97]

**Metadata curation, Samples and logistics, and Sequencing and analysis:**
Dr James P McKenna PhD [3], Dr Ben Warne MRCP [9], Joshua F Taylor MSc [22], Yasmin Chaudhry BSc [24], Rhys Izuagbe [24], Dr Aminu S Jahun PhD [24], Dr Gregory R Young PhD [36, 65], Dr Claire McMurray PhD [43], Dr Clare M McCann PhD [65, 66], Dr Andrew Nelson PhD [65, 66] and Scott Elliott [68]

**Metadata curation, Samples and logistics, and Visualisation:**
Hannah Lowe MSc [25]

**Metadata curation, Sequencing and analysis, and Software and analysis tools:**
Dr Anna Price PhD [11], Matthew R Crown BSc [65], Dr Sara Rey PhD [74], Dr Sunando Roy PhD [96] and Dr Ben Temperton PhD [105]

**Metadata curation, Sequencing and analysis, and Visualisation:**
Dr Sharif Shaaban PhD [73] and Dr Andrew R Hesketh PhD [101]

**Project administration, Samples and logistics, and Sequencing and analysis:**
Dr Kenneth G Laing PhD[41], Dr Irene M Monahan PhD [41] and Dr Judith Heaney PhD [95, 96, 34]

**Project administration, Samples and logistics, and Visualisation:**
Dr Emanuela Pelosi FRCPath [97], Siona Silviera MSc [97] and Dr Eleri Wilson-Davies MD, FRCPath [97]

**Samples and logistics, Software and analysis tools, and Visualisation:**
Dr Helen Fryer PhD [5]

**Sequencing and analysis, Software and analysis tools, and Visualization:**

Dr Helen Adams PhD [4], Dr Louis du Plessis PhD [23], Dr Rob Johnson PhD [39], Dr William T Harvey PhD [53, 42], Dr Joseph Hughes PhD [53], Dr Richard J Orton PhD [53], Dr Lewis G Spurgin PhD [59], Dr Yann Bourgeois PhD [81], Dr Chris Ruis PhD [102], Áine O'Toole MSc [104], Marina Gourtovaia MSc [116] and Dr Theo Sanderson PhD [116]

**Funding acquisition, and Leadership and supervision:**
Dr Christophe Fraser PhD [5], Dr Jonathan Edgeworth PhD, FRCPath [12], Prof Judith Breuer MD [96, 29], Dr Stephen L Michell PhD [105] and Prof John A Todd PhD [115]

**Funding acquisition, and Project administration:**
Michaela John BSc [10] and Dr David Buck PhD [115]

**Leadership and supervision, and Metadata curation:**
Dr Kavitha Gajee MBBS, FRCPath [37] and Dr Gemma L Kay PhD [75]

**Leadership and supervision, and Project administration:**
Prof Sharon J Peacock PhD [20, 70] and David Heyburn [74]

**Leadership and supervision, and Samples and logistics:**
Katie Kitchman BSc [37], Prof Alan McNally PhD [43, 93], David T Pritchard MSc, CSci [50], Dr Samir Dervisevic FRCPath [58], Dr Peter Muir PhD [70], Dr Esther Robinson PhD [70, 35], Dr Barry B Vipond PhD [70], Newara A Ramadan MSc, CSci, FIBMS [78], Dr Christopher Jeanes MBBS [90], Danni Weldon BSc [116], Jana Catalan MSc [118] and Neil Jones MSc [118]

**Leadership and supervision, and Sequencing and analysis:**
Dr Ana da Silva Filipe PhD [53], Dr Chris Williams MBBS [74], Marc Fuchs BSc [77], Dr Julia Miskelly PhD [77], Dr Aaron R Jeffries PhD [105], Karen Oliver BSc [116] and Dr Naomi R Park PhD [116]

**Metadata curation, and Samples and logistics:**
Amy Ash BSc [1], Cherian Koshy MSc, CSci, FIBMS [1], Magdalena Barrow [7], Dr Sarah L Buchan PhD [7], Dr Anna Mantzouratou PhD [7], Dr Gemma Clark PhD [15], Dr Christopher W Holmes PhD [16], Sharon Campbell MSc [17], Thomas Davis MSc [21], Ngee Keong Tan MSc [22], Dr Julianne R Brown PhD [29], Dr Kathryn A Harris PhD [29, 2], Stephen P Kidd MSc [33], Dr Paul R Grant PhD [34], Dr Li Xu-McCrae PhD [35], Dr Alison Cox PhD [38, 63], Pinglawathee Madona [38, 63], Dr Marcus Pond PhD [38, 63], Dr Paul A Randell MBBCh [38, 63], Karen T Withell FIBMS [48], Cheryl Williams MSc [51], Dr Clive Graham MD [60], Rebecca Denton-Smith BSc [62], Emma Swindells BSc [62], Robyn Turnbull BSc [62], Dr Tim J Sloan PhD [67], Dr Andrew Bosworth PhD [70, 35], Stephanie Hutchings [70], Hannah M Pymont MSc [70], Dr Anna Casey PhD [76], Dr Liz Ratcliffe PhD [76], Dr Christopher R Jones PhD [79, 105], Dr Bridget A Knight PhD [79, 105], Dr Tanzina Haque PhD, FRCPath [80], Dr Jennifer Hart MRCP [80], Dr Dianne Irish-Tavares FRCPath [80], Eric Witele MSc [80], Craig Mower BA [86], Louisa K Watson DipHE [86], Jennifer Collins BSc [89], Gary Eltringham BSc [89], Dorian Crudgington [98], Ben Macklin [98], Prof Miren Iturriza-Gomara PhD [107], Dr Anita O Lucaci PhD [107] and Dr Patrick C McClure PhD [113]

**Metadata curation, and Sequencing and analysis:**
Matthew Carlile BSc [18], Dr Nadine Holmes PhD [18], Dr Christopher Moore PhD [18], Dr Nathaniel Storey PhD [29], Dr Stefan Rooke PhD [73], Dr Gonzalo Yebra PhD [73], Dr Noel Craine DPhil [74], Malorie Perry MSc [74], Dr Nabil-Fareed Alikhan PhD [75], Dr Stephen Bridgett PhD [77], Kate F Cook MScR [84], Christopher Fearn MSc [84], Dr Salman Goudarzi PhD [84], Prof Ronan A Lyons MD [88], Dr Thomas Williams MD [104], Dr Sam T Haldenby PhD [107], Jillian Durham BSc [116] and Dr Steven Leonard PhD [116]

**Metadata curation, and Software and analysis tools:**
Robert M Davies MA (Cantab) [116]

**Project administration, and Samples and logistics:**
Dr Rahul Batra MD [12], Beth Blane BSc [20], Dr Moira J Spyer PhD [30, 95, 96], Perminder Smith MSc [32, 112], Mehmet Yavus [85, 109], Dr Rachel J Williams PhD [96], Dr Adhyana IK Mahanama MD [97], Dr Buddhini Samaraweera MD

[97], Sophia T Girgis MSc [102], Samantha E Hansford CSci [109], Dr Angie Green PhD [115], Dr Charlotte Beaver PhD [116], Katherine L Bellis [116, 102], Matthew J Dorman [116], Sally Kay [116], Liam Prestwood [116] and Dr Shavanthi Rajatileka PhD [116]

**Project administration, and Sequencing and analysis:**
Dr Joshua Quick PhD [43]

**Project administration, and Software and analysis tools:**
Radoslaw Poplawski BSc [43]

**Samples and logistics, and Sequencing and analysis:**
Dr Nicola Reynolds PhD [8], Andrew Mack MPhil [11], Dr Arthur Morriss PhD [11], Thomas Whalley BSc [11], Bindi Patel BSc [12], Dr Iliana Georgana PhD [24], Dr Myra Hosmillo PhD [24], Malte L Pinckert MPhil [24], Dr Joanne Stockton PhD [43], Dr John H Henderson PhD [65], Amy Hollis HND [65], Dr William Stanley PhD [65], Dr Wen C Yew PhD [65], Dr Richard Myers PhD [72], Dr Alicia Thornton PhD [72], Alexander Adams BSc [74], Tara Annett BSc [74], Dr Hibo Asad PhD [74], Alec Birchley MSc [74], Jason Coombes BSc [74], Johnathan M Evans MSc [74], Laia Fina [74], Bree Gatica-Wilcox MPhil [74], Lauren Gilbert [74], Lee Graham BSc [74], Jessica Hey BSc [74], Ember Hilvers MPH [74], Sophie Jones MSc [74], Hannah Jones [74], Sara Kumziene-Summerhayes MSc [74], Dr Caoimhe McKerr PhD [74], Jessica Powell BSc [74], Georgia Pugh [74], Sarah Taylor [74], Alexander J Trotter MRes [75], Charlotte A Williams BSc [96], Leanne M Kermack MSc [102], Benjamin H Foulkes MSc [109], Marta Gallis MSc [109], Hailey R Hornsby MSc [109], Stavroula F Louka MSc [109], Dr Manoj Pohare PhD [109], Paige Wolverson MSc [109], Peijun Zhang MSc [109], George MacIntyre-Cockett BSc [115], Amy Trebes MSc [115], Dr Robin J Moll PhD [116], Lynne Ferguson MSc [117], Dr Emily J Goldstein PhD [117], Dr Alasdair Maclean PhD [117] and Dr Rachael Tomb PhD [117]

**Samples and logistics, and Software and analysis tools:**
Dr Igor Starinskij MSc, MRCP [53]

**Sequencing and analysis, and Software and analysis tools:**
Laura Thomson BSc [5], Joel Southgate MSc [11, 74], Dr Moritz UG Kraemer DPhil [23], Dr Jayna Raghwani PhD [23], Dr Alex E Zarebski PhD [23], Olivia Boyd MSc [39], Lily Geidelberg MSc [39], Dr Chris J Illingworth PhD [52], Dr Chris Jackson PhD [52], Dr David Pascall PhD [52], Dr Sreenu Vattipally PhD [53], Timothy M Freeman MPhil [109], Dr Sharon N Hsu PhD [109], Dr Benjamin B Lindsey MRCP [109], Dr Keith James PhD [116], Kevin Lewis [116], Gerry Tonkin-Hill [116] and Dr Jaime M Tovar-Corona PhD [116]

**Sequencing and analysis, and Visualisation:**
MacGregor Cox MSci [20]

**Software and analysis tools, and Visualisation:**
Dr Khalil Abudahab PhD [14, 116], Mirko Menegazzo [14], Ben EW Taylor MEng [14, 116], Dr Corin A Yeats PhD [14], Afrida Mukaddas BTech [53], Derek W Wright MSc [53], Dr Leonardo de Oliveira Martins PhD [75], Dr Rachel Colquhoun DPhil [104], Verity Hill [104], Dr Ben Jackson PhD [104], Dr JT McCrone PhD [104], Dr Nathan Medd PhD [104], Dr Emily Scher PhD [104] and Jon-Paul Keatley [116]

**Leadership and supervision:**
Dr Tanya Curran PhD [3], Dr Sian Morgan FRCPath [10], Prof Patrick Maxwell PhD [20], Prof Ken Smith PhD [20], Dr Sahar Eldirdiri MBBS, MSc, FRCPath [21], Anita Kenyon MSc [21], Prof Alison H Holmes MD [38, 57], Dr James R Price PhD [38, 57], Dr Tim Wyatt PhD [69], Dr Alison E Mather PhD [75], Dr Timofey Skvortsov PhD [77] and Prof John A Hartley PhD [96]

**Metadata curation:**
Prof Martyn Guest PhD [11], Dr Christine Kitchen PhD [11], Dr Ian Merrick PhD [11], Robert Munn BSc [11], Dr Beatrice Bertolusso Degree [33], Dr Jessica Lynch MBCHB [33], Dr Gabrielle Vernet MBBS [33], Stuart Kirk MSc [34], Dr Elizabeth Wastnedge MD [56], Dr Rachael Stanley PhD [58], Giles Idle [64], Dr Declan T Bradley PhD [69, 77], Dr Jennifer Poyner MD [79] and Matilde Mori BSc [110]

**Project administration:**
Owen Jones BSc [11], Victoria Wright BSc [18], Ellena Brooks MA [20], Carol M Churcher BSc [20], Mireille Fragakis HND [20], Dr Katerina Galai PhD [20, 70], Dr Andrew Jermy PhD [20], Sarah Judges BA [20], Georgina M McManus BSc [20], Kim S Smith [20], Dr Elaine Westwick PhD [20], Dr Stephen W Attwood PhD [23], Dr Frances Bolt PhD [38, 57], Dr Alisha Davies PhD [74], Elen De Lacy MPH [74], Fatima Downing [74], Sue Edwards [74], Lizzie Meadows MA [75], Sarah Jeremiah MSc [97], Dr Nikki Smith PhD [109] and Luke Foulser [116]

**Samples and logistics:**
Dr Themoula Charalampous PhD [12, 46], Amita Patel BSc [12], Dr Louise Berry PhD [15], Dr Tim Boswell PhD [15], Dr Vicki M Fleming PhD [15], Dr Hannah C Howson-Wells PhD [15], Dr Amelia Joseph PhD [15], Manjinder Khakh [15], Dr Michelle M Lister PhD [15], Paul W Bird MSc, MRes [16], Karlie Fallon [16], Thomas Helmer [16], Dr Claire L McMurray PhD [16], Mina Odedra BSc [16], Jessica Shaw BSc [16], Dr Julian W Tang PhD [16], Nicholas J Willford MSc [16], Victoria Blakey BSc [17], Dr Veena Raviprakash MD [17], Nicola Sheriff BSc [17], Lesley-Anne Williams BSc [17], Theresa Feltwell MSc [20], Dr Luke Bedford PhD [26], Dr James S Cargill PhD [27], Warwick Hughes MSc [27], Dr Jonathan Moore MD [28], Susanne Stonehouse BSc [28], Laura Atkinson MSc [29], Jack CD Lee MSc [29], Dr Divya Shah PhD [29], Adela Alcolea-Medina Clinical scientist [32, 112], Natasha Ohemeng-Kumi MSc [32, 112], John Ramble MSc [32, 112], Jasveen Sehmi MSc [32, 112], Dr Rebecca Williams BMBS [33], Wendy Chatterton MSc [34], Monika Pusok MSc [34], William Everson MSc [37], Anibolina Castigador IBMS HCPC [44], Emily Macnaughton FRCPath [44], Dr Kate El Bouzidi MRCP [45], Dr Temi Lampejo FRCPath [45], Dr Malur Sudhanva FRCPath [45], Cassie Breen BSc [47], Dr Graciela Sluga MD, MSc [48], Dr Shazaad SY Ahmad MSc [49, 70], Dr Ryan P George PhD [49], Dr Nicholas W Machin MSc [49, 70], Debbie Binns BSc [50], Victoria James BSc [50], Dr Rachel Blacow MBCHB [55], Dr Lindsay Coupland PhD [58], Dr Louise Smith PhD [59], Dr Edward Barton MD [60], Debra Padgett BSc [60], Garren Scott BSc [60], Dr Aidan Cross MBCHB [61], Dr Mariyam Mirfenderesky FRCPath [61], Jane Greenaway MSc [62], Kevin Cole [64], Phillip Clarke [67], Nichola Duckworth [67], Sarah Walsh [67], Kelly Bicknell [68], Robert Impey MSc [68], Dr Sarah Wyllie PhD [68], Richard Hopes [70], Dr Chloe Bishop PhD [72], Dr Vicki Chalker PhD [72], Dr Ian Harrison PhD [72], Laura Gifford MSc [74], Dr Zoltan Molnar PhD [77], Dr Cressida Auckland FRCPath [79], Dr Cariad Evans PhD [85, 109], Dr Kate Johnson PhD [85, 109], Dr David G Partridge FRCP, FRCPath [85, 109], Dr Mohammad Raza PhD [85, 109], Paul Baker MD [86], Prof Stephen Bonner PhD [86], Sarah Essex [86], Leanne J Murray [86], Andrew I Lawton MSc [87], Dr Shirelle Burton-Fanning MD [89], Dr Brendan AI Payne MD [89], Dr Sheila Waugh MD [89], Andrea N Gomes MSc [91], Maimuna Kimuli MSc [91], Darren R Murray MSc [91], Paula Ashfield MSc [92], Dr Donald Dobie MBCHB [92], Dr Fiona Ashford PhD [93], Dr Angus Best PhD [93], Dr Liam Crawford PhD [93], Dr Nicola Cumley PhD [93], Dr Megan Mayhew PhD [93], Dr Oliver Megram PhD [93], Dr Jeremy Mirza PhD [93], Dr Emma Moles-Garcia PhD [93], Dr Benita Percival PhD [93], Megan Driscoll BSc [96], Leah Ensell BSc [96], Dr Helen L Lowe PhD [96], Laurentiu Maftei BSc [96], Matteo Mondani MSc [96], Nicola J Chaloner BSc [99], Benjamin J Cogger BSc [99], Lisa J Easton MSc [99], Hannah Huckson BSc [99], Jonathan Lewis MSc, PgD, FIBMS [99], Sarah Lowdon BSc [99], Cassandra S Malone MSc [99], Florence Munemo BSc [99], Manasa Mutingwende MSc [99], Roberto Nicodemi BSc [99], Olga Podplomyk FD [99], Thomas Somassa BSc [99], Dr Andrew Beggs PhD [100], Dr Alex Richter PhD [100], Claire Cormie [102], Joana Dias MSc [102], Sally Forrest BSc [102], Dr Ellen E Higginson PhD [102], Mailis Maes MPhil [102], Jamie Young BSc [102], Dr Rose K Davidson PhD [103], Kathryn A Jackson MSc [107], Dr Lance Turtle PhD, MRCP [107], Dr Alexander J Keeley MRCP [109], Prof Jonathan Ball PhD [113], Timothy Byaruhanga MSc [113], Dr Joseph G Chappell PhD [113], Jayasree Dey MSc [113], Jack D Hill MSc [113], Emily J Park MSc [113], Arezou Fanaie MSc [114], Rachel A Hilson MSc [114], Geraldine Yaze MSc [114] and Stephanie Lo [116]

**Sequencing and analysis:**
Safiah Afifi BSc [10], Robert Beer BSc [10], Joshua Maksimovic FD [10], Kathryn McCluggage Masters [10], Karla Spellman FD [10], Catherine Bresner BSc [11], William Fuller BSc [11], Dr Angela Marchbank BSc [11], Trudy Workman HNC [11], Dr Ekaterina Shelest PhD [13, 81], Dr Johnny Debebe PhD [18], Dr Fei Sang PhD [18], Dr Marina Escalera Zamudio PhD [23], Dr Sarah Francois PhD [23], Bernardo Gutierrez MSc [23], Dr Tetyana I Vasylyeva DPhil [23], Dr Flavia Flaviani PhD [31], Dr Manon Ragonnet-Cronin PhD [39], Dr Katherine L Smollett PhD [42], Alice Broos BSc [53], Daniel Mair BSc [53], Jenna Nichols BSc [53], Dr Kyriaki Nomikou PhD [53], Dr Lily Tong PhD [53], Ioulia Tsatsani MSc [53], Prof Sarah O'Brien PhD [54], Prof Steven Rushton PhD [54], Dr Roy Sanderson PhD [54], Dr Jon Perkins MBCHB [55], Seb Cotton MSc [56], Abbie Gallagher BSc [56], Dr Elias Allara MD, PhD [70, 102], Clare Pearson MSc [70, 102], Dr David Bibby PhD [72], Dr Gavin Dabrera PhD [72], Dr Nicholas Ellaby PhD [72], Dr Eileen Gallagher PhD [72],

Dr Jonathan Hubb PhD [72], Dr Angie Lackenby PhD [72], Dr David Lee PhD [72], Nikos Manesis [72], Dr Tamyo Mbisa PhD [72], Dr Steven Platt PhD [72], Katherine A Twohig [72], Dr Mari Morgan PhD [74], Alp Aydin MSci [75], David J Baker BEng [75], Dr Ebenezer Foster-Nyarko PhD [75], Dr Sophie J Prosolek PhD [75], Steven Rudder [75], Chris Baxter BSc [77], Sílvia F Carvalho MSc [77], Dr Deborah Lavin PhD [77], Dr Arun Mariappan PhD [77], Dr Clara Radulescu PhD [77], Dr Aditi Singh PhD [77], Miao Tang MD [77], Helen Morcrette BSc [79], Nadua Bayzid BSc [96], Marius Cotic MSc [96], Dr Carlos E Balcazar PhD [104], Dr Michael D Gallagher PhD [104], Dr Daniel Maloney PhD [104], Thomas D Stanton BSc [104], Dr Kathleen A Williamson PhD [104], Dr Robin Manley PhD [105], Michelle L Michelsen BSc [105], Dr Christine M Sambles PhD [105], Dr David J Studholme PhD [105], Joanna Warwick-Dugdale BSc [105], Richard Eccles MSc [107], Matthew Gemmell MSc [107], Dr Richard Gregory PhD [107], Dr Margaret Hughes PhD [107], Charlotte Nelson MSc [107], Dr Lucille Rainbow PhD [107], Dr Edith E Vamos PhD [107], Hermione J Webster BSc [107], Dr Mark Whitehead PhD [107], Claudia Wierzbicki BSc [107], Dr Adrienn Angyal PhD [109], Dr Luke R Green PhD [109], Dr Max Whiteley PhD [109], Emma Betteridge BSc [116], Dr Iraad F Bronner PhD [116], Ben W Farr BSc [116], Scott Goodwin MSc [116], Dr Stefanie V Lensing PhD [116], Shane A McCarthy [116, 102], Dr Michael A Quail PhD [116], Diana Rajan MSc [116], Dr Nicholas M Redshaw PhD [116], Carol Scott [116], Lesley Shirley MSc [116] and Scott AJ Thurston BSc [116]

**Software and analysis tools:**
Dr Will Rowe PhD[43], Amy Gaskin MSc [74], Dr Thanh Le-Viet PhD [75], James Bonfield BSc [116], Jennifier Liddle [116] and Andrew Whitwham BSc [116]

**1** Barking, Havering and Redbridge University Hospitals NHS Trust, **2** Barts Health NHS Trust, **3** Belfast Health & Social Care Trust, **4** Betsi Cadwaladr University Health Board, **5** Big Data Institute, Nuffield Department of Medicine, University of Oxford, **6** Blackpool Teaching Hospitals NHS Foundation Trust, **7** Bournemouth University, **8** Cambridge Stem Cell Institute, University of Cambridge, **9** Cambridge University Hospitals NHS Foundation Trust, **10** Cardiff and Vale University Health Board, **11** Cardiff University, **12** Centre for Clinical Infection and Diagnostics Research, Department of Infectious Diseases, Guy's and St Thomas' NHS Foundation Trust, **13** Centre for Enzyme Innovation, University of Portsmouth, **14** Centre for Genomic Pathogen Surveillance, University of Oxford, **15** Clinical Microbiology Department, Queens Medical Centre, Nottingham University Hospitals NHS Trust, **16** Clinical Microbiology, University Hospitals of Leicester NHS Trust, **17** County Durham and Darlington NHS Foundation Trust, **18** Deep Seq, School of Life Sciences, Queens Medical Centre, University of Nottingham, **19** Department of Infectious Diseases and Microbiology, Cambridge University Hospitals NHS Foundation Trust, **20** Department of Medicine, University of Cambridge, **21** Department of Microbiology, Kettering General Hospital, **22** Department of Microbiology, South West London Pathology, **23** Department of Zoology, University of Oxford, **24** Division of Virology, Department of Pathology, University of Cambridge, **25** East Kent Hospitals University NHS Foundation Trust, **26** East Suffolk and North Essex NHS Foundation Trust, **27** East Sussex Healthcare NHS Trust**, 28** Gateshead Health NHS Foundation Trust, **29** Great Ormond Street Hospital for Children NHS Foundation Trust, **30** Great Ormond Street Institute of Child Health (GOS ICH), University College London (UCL), **31** Guy's and St. Thomas' Biomedical Research Centre, **32** Guy's and St. Thomas' NHS Foundation Trust, **33** Hampshire Hospitals NHS Foundation Trust, **34** Health Services Laboratories, **35** Heartlands Hospital, Birmingham, **36** Hub for Biotechnology in the Built Environment, Northumbria University, **37** Hull University Teaching Hospitals NHS Trust, **38** Imperial College Healthcare NHS Trust, **39** Imperial College London, **40** Infection Care Group, St George's University Hospitals NHS Foundation Trust, **41** Institute for Infection and Immunity, St George's University of London, **42** Institute of Biodiversity, Animal Health & Comparative Medicine, **43** Institute of Microbiology and Infection, University of Birmingham, **44** Isle of Wight NHS Trust, **45** King's College Hospital NHS Foundation Trust, **46** King's College London, **47** Liverpool Clinical Laboratories, **48** Maidstone and Tunbridge Wells NHS Trust, **49** Manchester University NHS Foundation Trust, **50** Microbiology Department, Buckinghamshire Healthcare NHS Trust, **51** Microbiology, Royal Oldham Hospital, **52** MRC Biostatistics Unit, University of Cambridge, **53** MRC-University of Glasgow Centre for Virus Research, **54** Newcastle University, **55** NHS Greater Glasgow and Clyde, **56** NHS Lothian, **57** NIHR Health Protection Research Unit in HCAI and AMR, Imperial College London, **58** Norfolk and Norwich University Hospitals NHS Foundation Trust, **59** Norfolk County Council, **60** North Cumbria Integrated Care NHS Foundation Trust, **61** North Middlesex University Hospital NHS Trust, **62** North Tees and Hartlepool NHS Foundation Trust, **63** North West London Pathology, **64** Northumbria Healthcare NHS Foundation Trust, **65** Northumbria University, **66** NU-OMICS,

Northumbria University, **67** Path Links, Northern Lincolnshire and Goole NHS Foundation Trust, **68** Portsmouth Hospitals University NHS Trust, **69** Public Health Agency, Northern Ireland, **70** Public Health England, **71** Public Health England, Cambridge, **72** Public Health England, Colindale, **73** Public Health Scotland, **74** Public Health Wales, **75** Quadram Institute Bioscience, **76** Queen Elizabeth Hospital, Birmingham, **77** Queen's University Belfast, **78** Royal Brompton and Harefield Hospitals, **79** Royal Devon and Exeter NHS Foundation Trust, **80** Royal Free London NHS Foundation Trust, **81** School of Biological Sciences, University of Portsmouth, **82** School of Health Sciences, University of Southampton, **83** School of Medicine, University of Southampton, **84** School of Pharmacy & Biomedical Sciences, University of Portsmouth, **85** Sheffield Teaching Hospitals NHS Foundation Trust, **86** South Tees Hospitals NHS Foundation Trust, **87** Southwest Pathology Services, **88** Swansea University, **89** The Newcastle upon Tyne Hospitals NHS Foundation Trust, **90** The Queen Elizabeth Hospital King's Lynn NHS Foundation Trust, **91** The Royal Marsden NHS Foundation Trust, **92** The Royal Wolverhampton NHS Trust, **93** Turnkey Laboratory, University of Birmingham, **94** University College London Division of Infection and Immunity**, 95** University College London Hospital Advanced Pathogen Diagnostics Unit**, 96** University College London Hospitals NHS Foundation Trust, **97** University Hospital Southampton NHS Foundation Trust, **98** University Hospitals Dorset NHS Foundation Trust, **99** University Hospitals Sussex NHS Foundation Trust, **100** University of Birmingham, **101** University of Brighton, **102** University of Cambridge, **103** University of East Anglia, **104** University of Edinburgh, **105** University of Exeter, **106** University of Kent, **107** University of Liverpool, **108** University of Oxford, **109** University of Sheffield, **110** University of Southampton, **111** University of St Andrews, **112** Viapath, Guy's and St Thomas' NHS Foundation Trust, and King's College Hospital NHS Foundation Trust, **113** Virology, School of Life Sciences, Queens Medical Centre, University of Nottingham, **114** Watford General Hospital, **115** Wellcome Centre for Human Genetics, Nuffield Department of Medicine, University of Oxford, **116** Wellcome Sanger Institute, **117** West of Scotland Specialist Virology Centre, NHS Greater Glasgow and Clyde, **118** Whittington Health NHS Trust
